# Efficacy, Immunogenicity, and Safety of an Inactivated Bivalent EV-A71-CVA16 Vaccine in Children: A Randomized Phase 3 Trial

**DOI:** 10.64898/2026.11.23.26368796

**Authors:** Yeqing Tong, Hui Liang, Jihai Tang, Cangning Wang, Zhao Wang, Jianxiong Xiao, Gang Zeng, Ran Wu, Lei Wang, Chao Cheng, Yunrui Sun, Mei Li, Jing He, Zhuhang Huang, Yuyi Liao, Kun Xuan, Tao Li, Xianwei Luo, Lifang Huang, Junrong Li, Weiming Wang, Fan Gao, Qian Wang, Xing Wu, Weixiao Han, Jinxia Chen, Xing Han, Juan Yang, Jing Qian, Jiabing Wu, Dongjuan Zhang, Jikai Zhang, Xiao Ma, Xiaoqiang Liu, Jing Li

## Abstract

**Background:** Enterovirus A71 (EV-A71) and coxsackievirus A16 (CVA16) cause hand, foot, and mouth disease (HFMD). We evaluated the efficacy, immunogenicity, and safety of a bivalent EV-A71–CVA16 vaccine.

**Methods:** In this multicenter, double-blind, phase 3 trial, 8,000 healthy children aged 6–71 months were randomly assigned (1:1) to two doses of the bivalent or a licensed EV-A71 vaccine, 1 month apart. The primary efficacy endpoint was RT-PCR-confirmed CVA16-associated HFMD with onset at least 14 days after dose 2. EV-A71 immunogenicity noninferiority was assessed among baseline-seronegative participants without previous EV-A71 vaccination.

**Results:** Of 7,975 participants receiving at least one dose, 7,419 underwent the primary efficacy analysis. CVA16-associated HFMD occurred in 4 bivalent-group versus 44 EV-A71-group participants (vaccine efficacy, 90.91%; 95% CI, 75.03–97.63). At 30 days after dose 2, the EV-A71 geometric mean titer (GMT) ratio was 2.04 (95% CI, 1.71–2.44) among baseline-seronegative participants without previous EV-A71 vaccination. In the humoral immunogenicity population at this time point, CVA16 seroconversion rates were 96.94% versus 8.68%, and GMTs were 918.96 versus 3.88 U/mL. Adverse reactions within 30 days after either dose occurred in 46.73% versus 36.07% of reactogenicity-subgroup participants. In the overall safety population, serious adverse events occurred in 6.22% versus 7.55% from dose 1 through 6 months after dose 2; one bivalent-group event was considered possibly vaccine-related. No deaths occurred.

**Conclusions:** The bivalent EV-A71–CVA16 vaccine met the prespecified efficacy criterion against CVA16-associated HFMD and the EV-A71 immunogenicity noninferiority criterion in children aged 6–71 months (ClinicalTrials.gov: NCT06734832).

## Background

Hand, foot, and mouth disease (HFMD) is a common acute enteroviral infection worldwide, mainly affecting children under 5 years^1,2^. Most cases are self-limited, few progress to aseptic meningitis, autonomic nervous system dysfunction, or neurogenic pulmonary edema^3,4^. Severe neurologic disease may leave persistent cognitive or motor impairment^5^.

EV-A71 and CVA16 have historically represented the predominant etiological agents of HFMD^6^. Large-scale EV-A71 outbreaks have been documented across Asia^7,8^, Australia^9^, and Europe^10^, but mainland China has borne a disproportionate burden: during 2008–2012, 7.2 million probable HFMD cases were reported, including 82,486 severe cases with cardiopulmonary or neurological complications and 2,457 deaths^11^. Coinfection reportedly causes severe central nervous system complications and increases genetic recombination risk^12^. Recurrent outbreaks and sustained transmission have also been documented in Vietnam^13^, Malaysia^14^, and Thailand^15^.

EV-A71 vaccines, licensed in China since 2015, reached a cumulative coverage of 25% by 2021, with regional variation^16,17^ ^18^. Multiregional trials support broader use^19,20^, but data scarcity outside China highlights limited access and a persistent disease burden^21,22^. In China, EV-A71 outbreaks decreased from 40% in the pre-vaccine period to 3% post-vaccination, whereas CVA16 outbreaks increased from 49% to 60%^17^. Without licensed CVA16 vaccines or cross-protection from current monovalent EV-A71 vaccines^23,24^, continued cocirculation leaves a protection gap supporting bivalent vaccine development^13,15^.

We therefore conducted a randomized phase 3 trial in children aged 6–71 months to evaluate the efficacy of an investigational bivalent EV-A71–CVA16 vaccine against RT-PCR-confirmed CVA16-associated HFMD and assess its immunogenicity and safety.

## Methods

### 1. Study Design and Oversight

This multicenter, randomized, double-blind, active-controlled phase 3 trial was sponsored by Beijing Sinovac and led by the Yunnan Provincial CDC in collaboration with the CDCs of Guangdong, Hubei, Fujian, and Anhui. The National Institutes for Food and Drug Control (NIFDC) and participating institutions designed the protocol. Conducted at 16 county- and city-level CDCs, the trial followed the Declaration of Helsinki and Good Clinical Practice. Each center obtained ethics approval; each participant’s guardian provided written informed consent. The trial was registered at ClinicalTrials.gov (NCT06734832). The protocol and statistical analysis plan accompany the full text.

### 2. Participants

Healthy children aged 6–23 and 24–71 months were enrolled from December 12, 2024, to March 15, 2025. Eligible children had never received an EV-A71 vaccine or had completed a two-dose EV-A71 vaccination regimen at least 6 months before enrollment. Study populations and planned subgroup allocations are summarized in **Tables S1–S3**. Efficacy analyses included all randomized participants receiving two doses and entering the effective surveillance period. In a humoral immunogenicity subset of the reactogenicity subgroup, serum was collected before dose 1 and at 30 days, 6 months, and 12 months after dose 2. In a cellular immunogenicity subgroup of EV-A71 vaccine-naive children aged 24–71 months, blood was collected before and 14 days after each dose. For safety, a reactogenicity subgroup was established, stratified by age group and EV-A71 vaccination history. Children with a known history of HFMD or herpangina or other protocol-specified exclusion criterion were excluded (**Table S4**). Participants were randomized 1:1 to bivalent or EV-A71 vaccination using a stratified block-randomization schedule generated by an independent statistician. Randomization was centrally administered through an interactive web response system (IWRS); the allocation sequence was concealed from enrolling staff and participants, and vaccine was dispensed by IWRS-assigned number.

### 3. Vaccines

The investigational Vero cell-based bivalent vaccine contained inactivated EV-A71 and CVA16 antigens at 1,920 U and 1,200 SU, respectively, per 0.5-mL dose^25^. The control was the licensed inactivated EV-A71 vaccine (Inlive®, Sinovac), with 480U EV-A71 antigen. Both vaccines were supplied in identical packaging. Participants received two 0.5-mL doses 1 month apart^26^ (**Table S5)**.

### 4. Surveillance and Case Definitions

Case surveillance to evaluate vaccine efficacy (VE) began after dose 1 and continued through two consecutive HFMD epidemic seasons; the prespecified efficacy period began 14 days after dose 2 (**Table S6**). Cases were identified through active and passive surveillance. Staff contacted parents or guardians weekly during epidemic seasons and biweekly during non-epidemic periods by telephone, text message, or in-person visit (**Table S7**). Parents or guardians received education on early HFMD and herpangina manifestations and were instructed to promptly report suspected enterovirus-associated illness.

For suspected cases, throat and/or anal swabs were collected within 3 days of symptom onset and tested by RT-PCR at local laboratories using a licensed five-target assay for pan-enterovirus, EV-A71, CVA16, CVA10, and CVA6 RNA. If EV-A71 or CVA16 was detected, a second specimen was collected within 24 hours of the result; a third was collected if the first two results were discordant. Clinical manifestations, medical visits, and treatment were recorded. The NIFDC retested locally CVA16-positive samples, providing definitive results for discordant cases. The primary endpoint, virologically confirmed CVA16-associated HFMD, required compatible clinical manifestations and CVA16 RNA positivity at ≥2 sampling visits. Cases with only one positive sampling visit were included in supplementary analyses. The Clinical Event Adjudication Committee adjudicated all suspected cases using clinical and laboratory findings.

### 5. Efficacy end points

The primary efficacy end point was VE against RT-PCR-confirmed CVA16-associated HFMD. Secondary endpoints included CVA16-associated herpangina, severe disease, hospitalization, and all CVA16-associated diseases. VE against infections caused by EV-A71, CVA10, CVA6, and other enteroviruses was also assessed (**Table S8)**.

### 6. Immunogenicity

In the humoral immunogenicity subgroup, blood was collected before dose 1 and at 30 days, 6 months, and 12 months after dose 2; this report includes results through 6 months after dose 2. EV-A71 and CVA16 neutralizing antibodies were measured at the NIFDC by micro–cytopathic effect neutralization assay. Seropositivity was defined as a titer ≥1:8 for each virus. Antibody levels in seropositive samples were calibrated against national standards and expressed in IU/mL (EV-A71) and U/mL (CVA16).

Seroconversion was defined as postvaccination seropositivity (initially seronegative) or ≥4-fold antibody increase (initially seropositive). CVA6 and CVA10 neutralizing antibodies were measured before dose 1 and 30 days after dose 2 in 200 participants (first 100 by randomization number in each prior-vaccination stratum). In the cellular immunogenicity subgroup, blood was collected before and 14 days after each dose. ELISpot quantified EV-A71– and CVA16-responsive IFN-γ– and IL-4–secreting cells per 3 × 10⁶ peripheral-blood mononuclear cells (PBMCs).

### 7. Safety Evaluation

Events considered definitely, probably, or possibly related to vaccination were classified as adverse reactions (ARs). The prespecified safety end points were the incidence of adverse events (AEs) and ARs within 30 days after each dose and the incidence of serious adverse events (SAEs) through 6 months after dose 2. AE severity was graded according to *the 2019 guideline of the National Medical Products Administration* (**Table S9**).

### 8. Statistical Analysis

We estimated that 4000 participants per group were required, Assuming a 12-month CVA16-associated HFMD incidence of 1.2%, VE of 85%, and approximately 15% dropout or exclusion, we estimated 4,000 participants per group. With 47 observed cases, power was 90% for the 95% confidence interval (CI) lower bound for VE to be ≥50%, at an overall two-sided α of 0.05. No interim analysis or stopping guideline was prespecified.

VE was calculated as *[1−(incidence density in the experimental group ÷ incidence density in the control group)] × 100*^24^. Estimates used Poisson regression based on person-years of follow-up, with treatment group, age group, and previous EV-A71 vaccination as fixed effects. Secondary efficacy analyses were not adjusted for multiplicity. For primary immunogenicity analyses, noninferiority margins were −10% for the EV-A71 neutralizing-antibody seroconversion-rate difference and 2/3 for the geometric mean titer (GMT) ratio.

Primary efficacy analysis used the modified Full Analysis Set for Efficacy (E-mFAS), with supplementary analysis in the Per-Protocol Set for Efficacy (E-PPS). Primary humoral immunogenicity was analyzed in the modified Full Analysis Set for Humoral Immunogenicity (HI-mFAS); safety in the AE Safety Set (ASS) and SAE Safety Set (SSS). Antibody persistence at 6 and 12 months used the Humoral Immunity Persistence Sets (IPS-6 and IPS-12); cellular immunogenicity used the Cellular Immunogenicity Set (CIS). Participants without follow-up were assumed event-free; missing antibody values were multiply imputed (10 imputations), with tipping-point and jump-to-reference sensitivity analyses. Seroconversion rates, log-transformed titers, cellular responses, and AE frequencies were analyzed using Cochran–Mantel–Haenszel methods, analysis of covariance, rank-sum tests, and Fisher’s exact test, respectively. Analyses used SAS 9.4 and StatXact for stratified exact calculations.

## Results

### 1. Study Population

Of 8,937 children screened, 8,000 were randomly assigned 1:1 to the bivalent vaccine (n = 4,002) or the EV-A71 vaccine (n = 3,998) (**Figure 1**). Among them, 7,975 participants received at least one dose and were included in the SSS, while 7,422 (92.8%) participants completed the two-dose vaccination series and entered the post-vaccination surveillance period. For the efficacy analysis, 7,421 participants were included in the EV-A71 E-mFAS and 7,419 in the CVA16 E-mFAS. A total of 169 (2.11%) participants withdrew prematurely. Baseline demographic and clinical characteristics were well balanced between the two groups (**Table 1**).

**Figure 1.**
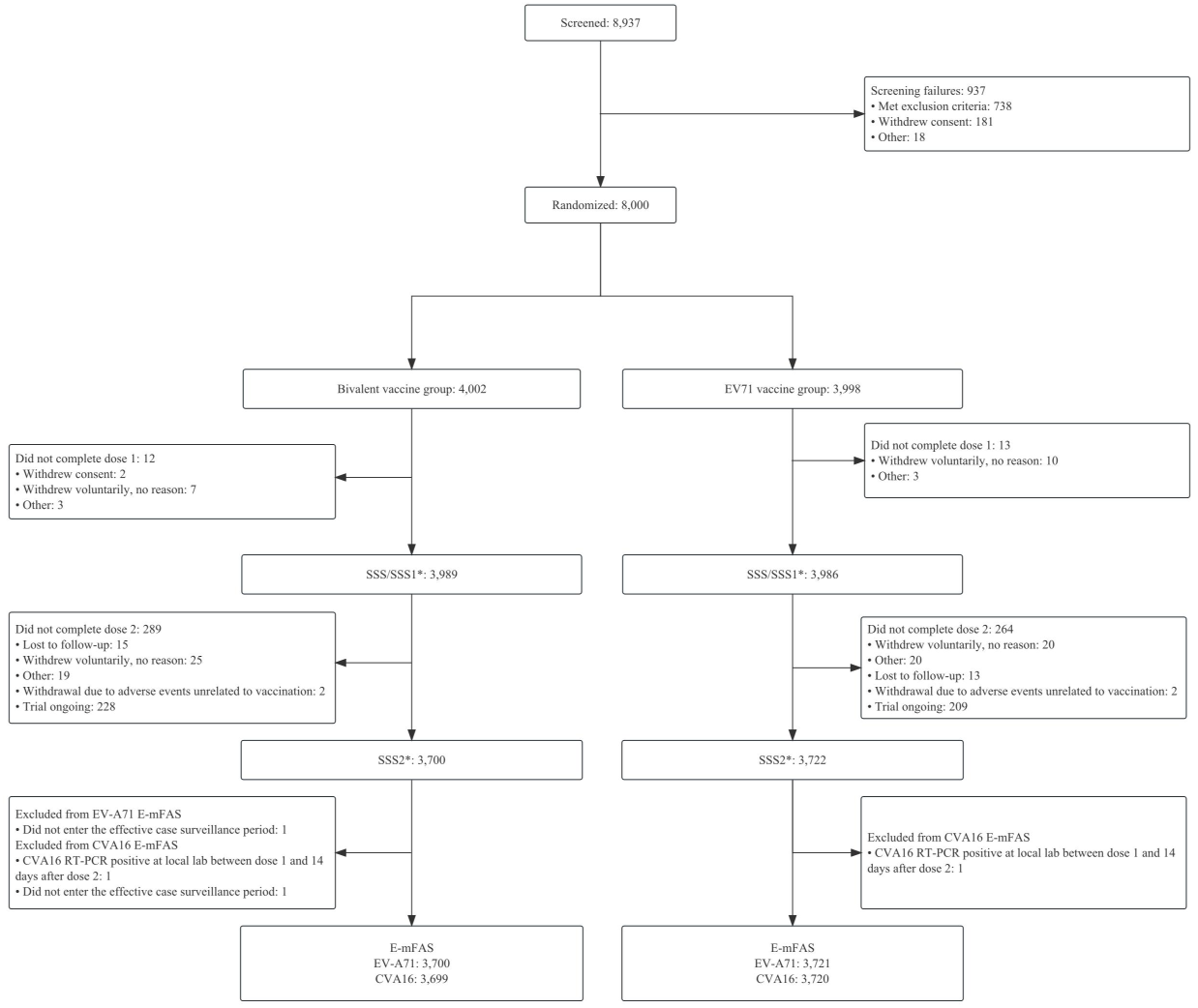
Screening, Randomization, and Inclusion in the EV-A71-CVA16 Bivalent Vaccine Clinical Trial (E-mFAS) * Participant R22186 in the non-reactogenicity subgroup was assigned to the vaccine group but mistakenly received two doses of the control vaccine. SSS, SSS1 and SSS2 analyses use the actual treatment (control group). Efficacy analyses use the assigned treatment (vaccine group)

**Table 1.** Baseline Characteristics of the Study Participants*.

| **Characteristic** | **Bivalent vaccine<sup>1</sup>group** | **EV-A71 vaccine group** |
|---|---|---|
| **E-mFAS** |  |  |
| **No. of participants** | 3,699 | 3,720 |
| **Age group (IWRS<sup>2</sup>)** |  |  |
| 6-23 months | 1,851 | 1,864 |
| 24-71 months | 1,848 | 1,856 |
| **Sex - Male, n (%)** | 1,880 (50.82) | 1,893 (50.89) |
| **Height, cm** | 87.72 ± 14.37 | 87.44 ± 14.13 |
| **Weight, kg** | 13.23 ± 4.09 | 13.18 ± 3.96 |
| **Previous EV-A71 vaccination (IWRS) - Yes, n (%)** | 190 (5.14) | 183 (4.92) |
| **HI-mFAS** |  |  |
| **No. of participants** | 549 | 556 |
| **Age group (IWRS<sup>2</sup>)** |  |  |
| 6-23 months | 271 (49.36) | 279 (50.18) |
| 24-71 months | 278 (50.64) | 277 (49.82) |
| **Sex - Male, n (%)** | 282 (51.37) | 274 (49.28) |
| **Height, cm** | 88.97 ± 14.95 | 88.52 ± 14.87 |
| **Weight, kg** | 13.62 ± 4.12 | 13.56 ± 4.38 |
| **Previous EV-A71 vaccination (IWRS) - Yes, n (%)** | 95 (17.30) | 91 (16.37) |
| **ASS** |  |  |
| **No. of participants** | 993 | 998 |
| **Age group (IWRS<sup>2</sup>)** |  |  |
| 6-23 months | 495 (49.85) | 499 (50.00) |
| 24-71 months | 498 (50.15) | 499 (50.00) |
| **Sex - Male, n (%)** | 488 (49.14) | 492 (49.30) |
| **Height, cm** | 88.80 ± 15.41 | 88.36 ± 14.83 |
| **Weight, kg** | 13.58 ± 4.26 | 13.54 ± 4.20 |
| **Previous EV-A71 vaccination (IWRS) - Yes, n (%)** | 100 (10.07) | 99 (9.92) |

\* Plus–minus values are means ± SD. E-mFAS, modified Full Analysis Set for Efficacy; HI-mFAS, modified Full Analysis Set for Humoral Immunogenicity; ASS, Adverse Event Safety Set.
<sup>1</sup> Bivalent vaccine, EV-A71-CVA16 vaccine (EV-A71, Enterovirus 71; CVA16, Coxsackievirus A16).
<sup>2</sup> IWRS, interactive web response system.

The EV-A71 and CVA16 E-PPS included 7,147 and 7,146 participants, respectively (**Figure S1**). Additional participant flowcharts for efficacy, humoral immunogenicity, and reactogenicity analyses are provided in **Figure S2-S8**.

### 2. Vaccine Efficacy

In the E-mFAS, from 14 days after dose 2, 4 versus 44 RT-PCR-confirmed CVA16-associated HFMD cases occurred in the bivalent and EV-A71 vaccine groups over 3,063.3 versus 3,058.1 person-years, respectively. Incidence rates were 0.13 versus 1.44 per 100 person-years (VE, 90.91%; 95% CI, 75.03–97.63; ***P***<0.0001). VE estimates were similar across age subgroups. Estimates stratified by previous EV-A71 vaccination status are shown in **Table 2**; the estimate for previously vaccinated participants was imprecise because of few cases. Per-protocol results were consistent with the primary analysis (**Table S10**).

**Table 2.** Efficacy against RT-PCR-Confirmed CVA16-Associated HFMD^1^ (E-mFAS^2^)

| **Subgroup** | **Bivalent vaccine group<sup>3</sup> (N=3699)** |  | **EV-A71 vaccine group (N=3720)** |  | **Vaccine efficacy, % (95% CI)** | ***P* value** |
|---|---|---|---|---|---|---|
|  | **Cases/person-years** | **Incidence per 100 person-years (95% CI)<sup>4</sup>** | **Cases/person-years** | **Incidence per 100 person-years (95% CI)** |  |  |
| Overall (6-71 months) | 4/3063.3 | 0.13 (0.05, 0.35) | 44/3058.1 | 1.44 (1.07, 1.93) | 90.91 (75.03, 97.63) | <0.0001 |
| **Age group** |  |  |  |  |  |  |
| 6-23 months | 3/1522.7 | 0.20 (0.06, 0.61) | 19/1525.6 | 1.25 (0.79, 1.95) | 84.18 (46.54, 95.32) | 0.0030 |
| 24-71 months | 1/1540.6 | 0.06 (0.01, 0.46) | 25/1532.5 | 1.63 (1.10, 2.41) | 96.01 (75.61, 99.90) | <0.0001 |
| **Previous EV-A71 vaccination** |  |  |  |  |  |  |
| Yes | 0/172.1 | 0.00 (0.00, 2.14) | 1/164.3 | 0.61 (0.02, 3.39) | 100.00 (-3623.64, 100.00) | 0.9769 |
| No | 4/2891.2 | 0.14 (0.05, 0.37) | 43/2893.7 | 1.49 (1.10, 2.00) | 90.69 (68.14, 97.28) | 0.0002 |
| 24-71 months, no previous EV-A71 vaccination | 1/1368.4 | 0.07 (0.01, 0.52) | 24/1368.2 | 1.75 (1.18, 2.62) | 95.83 (69.21, 99.44) | 0.0018 |

<sup>1</sup> HFMD, hand, foot, and mouth disease.
<sup>2</sup> E-mFAS, modified Full Analysis Set for Efficacy.
<sup>3</sup> Bivalent vaccine: EV-A71-CVA16 vaccine (EV-A71, Enterovirus 71; CVA16, Coxsackievirus A16).
<sup>4</sup> 95% CI, 95% confidence interval. Cases/person-years entries denote cases over person-years of observation. Incidence rates and 95% CIs were estimated using an unstratified Poisson model. Vaccine efficacy and 95% CIs were estimated using Poisson regression with age group, previous EV-A71 vaccination, and study group as fixed effects and log person-years as the offset; an exact stratified method was used when a stratum contained zero cases. Only one endpoint event occurred in this subgroup (EV-A71 vaccine group); the estimate is imprecise and should be interpreted cautiously.

For secondary endpoints, the bivalent and EV-A71 vaccine groups had 6 versus 49 CVA16-associated disease cases (VE, 87.76%; 95% CI, 71.39–95.71), 0 versus 5 CVA16-associated herpangina cases (VE, 100.00%; 95% CI, −9.15–100.00), and 0 versus 6 CVA16-associated hospitalizations (VE, 100.00%; 95% CI, 14.91–100.00). No severe CVA16-associated disease occurred in either group, precluding VE estimation. All 6 hospitalized children were aged 6–23 months (**Table S11**). VEs against CVA10- and CVA6-associated disease were 29.05% and 18.81% (95% CIs, −48.20–66.03 and 3.89–31.40), respectively (**Table S12**).

### 3. Immunogenicity

Baseline EV-A71 and CVA16 neutralizing-antibody levels were comparable between groups. At 30 days after dose 2, EV-A71 seroconversion rates were 99.27% versus 97.94%, and GMTs were 1996.54 versus 987.58 IU/mL in the bivalent and EV-A71 vaccine groups, respectively (GMT ratio, 2.02; 95% CI, 1.63–2.50) (**Figure 2, Table S13 and S14**).

**Figure 2.**
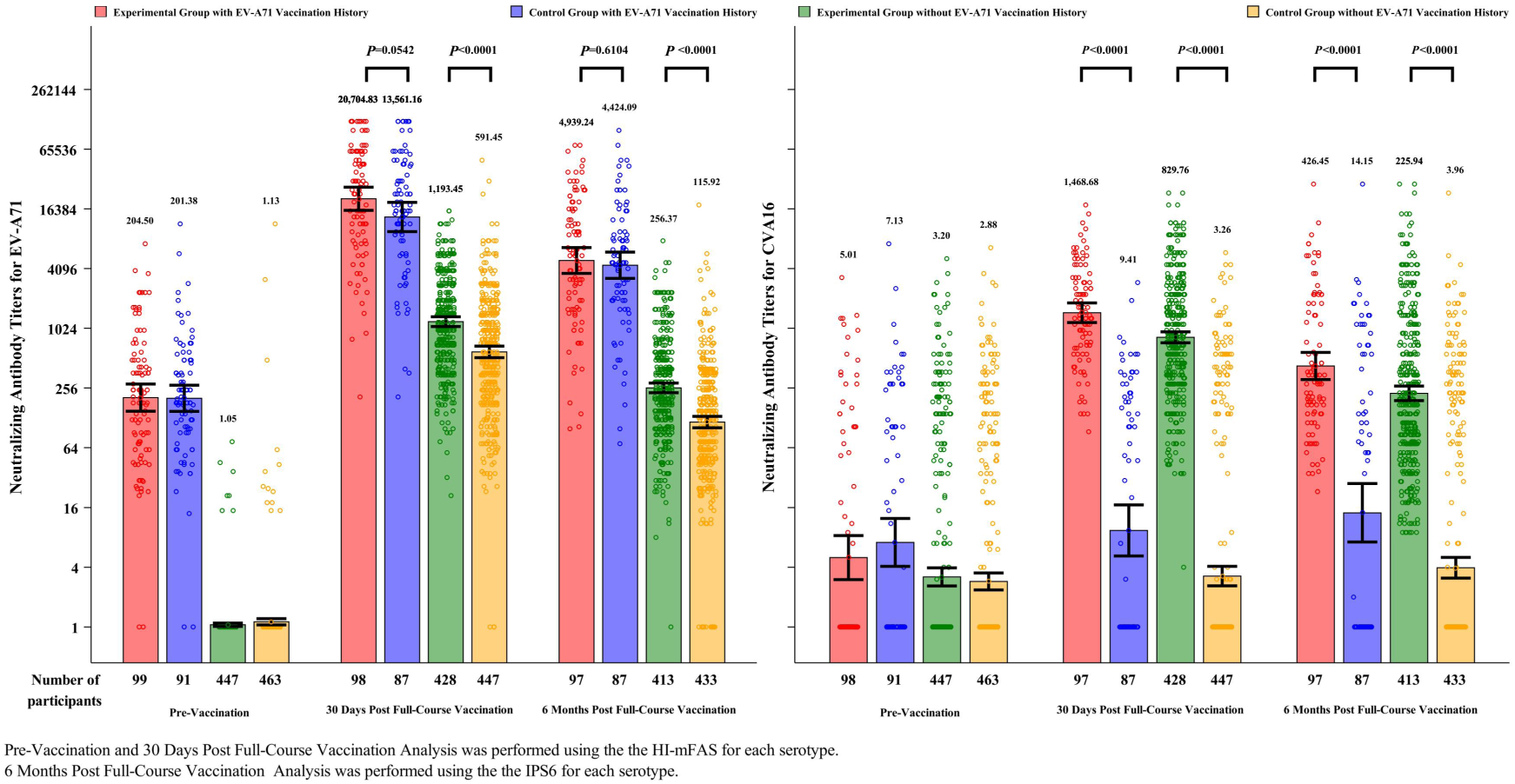
Neutralizing Antibody Levels among Participants Aged 6-71 Months (HI-mFAS/IPS6)

Among baseline-seronegative participants without previous EV-A71 vaccination, seroconversion rates 30 days after dose 2 were 100.00% vs 99.33% in the bivalent vs EV-A71 group (**Table S15**). The difference was 0.67 percentage points (95% CI, −0.08 to 1.42), exceeding the −10-percentage-point noninferiority margin; GMT ratio was 2.04 (95% CI, 1.71–2.44) (**Table S16**).

At 30 days after dose 2, CVA16 seropositivity rates were 100.00% versus 26.46%, seroconversion rates 96.94% versus 8.68%, and GMTs 918.96 versus 3.88 U/mL in the bivalent and EV-A71 vaccine groups, respectively (**Figure 2, Table S13**). CVA10 and CVA6 seroconversion rates were also higher in the bivalent group, but GMTs remained low, suggesting limited cross-reactive antibody responses (**Figure S9, Tables S13–S14**). At 6 months after dose 2, EV-A71 seropositivity remained high in both groups, with GMTs of 450.01 versus 213.20 IU/mL. CVA16 seropositivity remained 100.00% in the bivalent group, compared with 27.88% in the EV-A71 group, with GMTs of 254.95 versus 4.90 U/mL (**Figure 2, Table S13**). PPS analyses at 30 days after dose 2 were consistent with the HI-mFAS findings (**Figures S10–S11**).

Cellular immunogenicity was analyzed in 38 participants. Median frequencies of interferon-γ–secreting cells responding to EV-A71 and CVA16 were higher after vaccination than at baseline in both groups, with the highest observed medians 14 days after dose 2. Median frequencies in the bivalent and EV-A71 vaccine groups were 3,000 versus 2,435 spot-forming cells per 3 × 10⁶ PBMCs for EV-A71 (**Table S17**) and 2,740 versus 2,620 for CVA16 (**Table S18**). Median interleukin-4–secreting cell frequencies remained low across time points (**Tables S19-S20**). No statistically significant between-group differences in cell frequencies or their ratios to baseline were detected at any assessed time point (**Tables S17-S20**).

### 4. Adverse Reactions

From dose 1 through 6 months after dose 2, 779 SAEs occurred in 549 participants: 349 events in 248 participants (6.22%) in the bivalent group and 430 in 301 (7.55%) in the EV-A71 group. One SAE, a febrile seizure in the bivalent group, was considered possibly vaccine-related. No deaths occurred (**Tables S21-S22**). In the reactogenicity subgroup, 1,361/1,991 participants (68.36%) experienced ≥1 AE within 30 days after either dose: 704/993 (70.90%) in the bivalent group and 657/998 (65.83%) in the EV-A71 group (**Table S23**). Fever was the most common AE (40.58% vs. 28.76%) (**Table S24**). ARs occurred in 824 reactogenicity-subgroup participants (41.39%): 464 (46.73%) in the bivalent group and 360 (36.07%) in the EV-A71 vaccine group (**Table S23**). The higher AR rate in the bivalent group mainly reflected fever (29.71% vs. 14.53%). Grade 3 ARs occurred in 4.33% versus 2.40%; no Grade 4-5 ARs were reported (**Table 3**). Overall safety population AR rates were 18.15% and 13.47% in the bivalent and EV-A71 groups, respectively (**Table S25**).

**Table 3.** Adverse Reactions in the Reactogenicity Subgroup (ASS)

| **Adverse reaction** | **Any grade** |  | **Grade 3** |  |
|---|---|---|---|---|
|  | **Bivalent vaccine n/N (%)** | **EV-A71 vaccine n/N (%)** | **Bivalent vaccine n/N (%)** | **EV-A71 vaccine n/N (%)** |
| **Any adverse reaction** | 464/993 (46.73) | 360/998 (36.07) | 43/993 (4.33) | 24/998 (2.40) |
| **General disorders and administration site conditions** | 366/993 (36.86) | 224/998 (22.44) | 32/993 (3.22) | 18/998 (1.80) |
| Fever | 295/993 (29.71) | 145/998 (14.53) | 29/993 (2.92) | 17/998 (1.70) |
| Vaccination site pain | 74/993 (7.45) | 53/998 (5.31) | 0/993 (0.00) | 0/998 (0.00) |
| Postvaccination irritability | 32/993 (3.22) | 24/998 (2.40) | 3/993 (0.30) | 1/998 (0.10) |
| Decreased activity | 23/993 (2.32) | 14/998 (1.40) | 0/993 (0.00) | 0/998 (0.00) |
| Vaccination site swelling | 20/993 (2.01) | 20/998 (2.00) | 0/993 (0.00) | 0/998 (0.00) |
| Vaccination site erythema | 14/993 (1.41) | 22/998 (2.20) | 0/993 (0.00) | 0/998 (0.00) |
| **Respiratory, thoracic and mediastinal disorders** | 114/993 (11.48) | 120/998 (12.02) | 0/993 (0.00) | 0/998 (0.00) |
| Cough | 82/993 (8.26) | 85/998 (8.52) | 0/993 (0.00) | 0/998 (0.00) |
| Rhinorrhoea | 72/993 (7.25) | 66/998 (6.61) | 0/993 (0.00) | 0/998 (0.00) |
| **Gastrointestinal disorders** | 111/993 (11.18) | 113/998 (11.32) | 9/993 (0.91) | 7/998 (0.70) |
| Diarrhea | 72/993 (7.25) | 71/998 (7.11) | 3/993 (0.30) | 3/998 (0.30) |
| Vomiting | 58/993 (5.84) | 66/998 (6.61) | 7/993 (0.70) | 5/998 (0.50) |
| **Metabolism and nutrition disorders** | 88/993 (8.86) | 70/998 (7.01) | 1/993 (0.10) | 0/998 (0.00) |
| Decreased appetite | 88/993 (8.86) | 70/998 (7.01) | 1/993 (0.10) | 0/998 (0.00) |
| **Skin and subcutaneous tissue disorders** | 13/993 (1.31) | 16/998 (1.60) | 0/993 (0.00) | 0/998 (0.00) |
| **Immune system disorders** | 7/993 (0.70) | 2/998 (0.20) | 1/993 (0.10) | 0/998 (0.00) |
| Hypersensitivity | 7/993 (0.70) | 2/998 (0.20) | 1/993 (0.10) | 0/998 (0.00) |
| **Nervous system disorders** | 1/993 (0.10) | 0/998 (0.00) | 1/993 (0.10) | 0/998 (0.00) |
| Febrile convulsion | 1/993 (0.10) | 0/998 (0.00) | 1/993 (0.10) | 0/998 (0.00) |

Data are participants with adverse reactions, n/N (%). The main-text table includes the overall adverse-reaction summary, categories or events with an overall incidence of at least 1%, and any category or event with a Grade 3 reaction. Participants could report more than one reaction.

## 6. Discussion

In this trial, the bivalent EV-A71–CVA16 vaccine met the prespecified efficacy criterion against RT-PCR-confirmed CVA16-associated HFMD, with protection also observed against the broader CVA16-associated disease endpoint. EV-A71 immunogenicity met the prespecified noninferiority criterion in the designated population.

After dose 2, the vaccine elicited strong neutralizing-antibody responses against both viruses. Among participants without previous EV-A71 vaccination, EV-A71 GMTs were higher with the bivalent than the EV-A71 vaccine. Responses in previously vaccinated participants suggested potential to reinforce EV-A71 immunity while extending responses to CVA16, although the small subgroup was not powered for clinical efficacy. Antibody levels declined during follow-up but remained higher in the bivalent group. CVA16 seropositivity was maintained, and GMTs remained above pre-vaccination levels in baseline-seropositive participants.

Although CVA6 point estimates suggested a modest reduction, secondary efficacy analyses were unadjusted for multiplicity and CVA10 estimates were imprecise. Notably, the absolute neutralizing antibody titers against CVA6 and CVA10 remained substantially lower than those against CVA16, suggesting that cross-reactive antibodies were unlikely to confer meaningful protection. As CVA6 and CVA10 have become prominent causes of HFMD^27,28^, continued etiologic surveillance and evaluation of higher-valency vaccines are warranted. Most ARs were Grade 1–2, self-limited, and less frequent after dose 2. Phase 3 trials of EV-A71 vaccines have also reported transient systemic reactions, including fever^24,29^. No Grade 4-5 ARs occurred, cumulative SAEs were numerically more frequent in the EV-A71 group, one SAE in the bivalent group was considered possibly vaccine-related. These data identify no additional safety signal but are insufficient to exclude rare events or risks that may emerge with wider use.

The multicenter randomized design, active control, blinded adjudication, and laboratory-confirmed primary endpoint supported the reliability of the main findings. Concordant analyses and integrated efficacy and immunogenicity assessments provided complementary evidence of vaccine performance. However, the absence of severe CVA16 cases precluded estimation of efficacy against severe disease, while underpowered subgroup analyses and the small cellular-immunity sample limited precision. This report includes one epidemic season of efficacy data and six months of immunogenicity data, leaving long-term protection uncertain. Given that the trial was conducted in China, effectiveness in other settings may differ with local serotype distribution, vaccination coverage, and implementation.

These findings support the bivalent vaccine’s potential to extend protection against CVA16-associated HFMD in children aged 6–71 months, particularly where EV-A71 vaccines are already used. Longer follow-up, larger safety datasets, and effectiveness studies in settings with different enterovirus distributions are needed to assess protection durability.

## Data Sharing Statement

Individual participant data that underlie the results reported in this article, after de-identification, and the study protocol and statistical analysis plan are available from the corresponding author upon reasonable request.

## Data Availability

Individual participant data that underlie the results reported in this article, after de-identification, and the study protocol and statistical analysis plan are available upon reasonable request.

## Acknowledgments

We are grateful to the children and their families for their participation. We thank the staff at the Yunnan, Guangdong, Hubei, Fujian, and Anhui Provincial Centers for Disease Control and Prevention for their dedication to this trial.

## Trial Registration

Clinical Trial Notification No. 2023LP01345; 2023LP01346; 2023LP01347.ClinicalTrials.gov (NCT06734832).

## Ethical Approval

This research protocol was approved by relevant institutional review boards or ethics committees and that all human participants gave written informed consent.

