## Supplemental file for "Efficacy, Immunogenicity, and Safety of an Inactivated Bivalent EV-A71-CVA16 Vaccine in Children: A Randomized Phase 3 Trial"

### Supplementary Appendix

Table S1. Bivalent Inactivated Enterovirus Vaccine Phase III Clinical Study Vaccine Efficacy Study Plan ........ 3
Table S2. Bivalent Inactivated Enterovirus Vaccine Phase III Clinical Study Reactogenicity Subgroup Study Plan ........ 5

**Table S1. Bivalent Inactivated Enterovirus Vaccine Phase III Clinical Study Vaccine Efficacy Study Plan**

| History of EV-A71 Vaccination | Month(s) of age group | Bivalent vaccine Group | EV-A71 vaccine Group | Total | Case monitoring period |
|---|---|---|---|---|---|
| **None** | 6-23 | 2000 | 2000 | 4000 | From First Dose Vaccination, continued until the end of two consecutive epidemic seasons. |
| **None** | 24-71 | 1800 | 1800 | 3600 | |
| **Yes** | 24-71 | 200 | 200 | 400 | |
| **Total** | | 4000 | 4000 | **8000** | |

**Table S2. Bivalent Enterovirus Inactivated Vaccine Phase III Clinical Study Immunogenicity Subgroup Study Plan**

| **Subgroup Name** | EV-A71 Vaccination history | Months of age group | Bivalent vaccine Group | EV-A71 vaccine Group | Total | **Blood collection time** | **Laboratory Tests** |
|---|---|---|---|---|---|---|---|
| Body fluid Immunogenicity subgroup | None | 6-23<br>24-71 | 290<br>190 | 290<br>190 | 580<br>380 | Before vaccination, 30 days post two doses of vaccination, post two doses of vaccination 6 months, 12 months | EV-A71, CVA16Neutralizing antibodies* |
| | Yes | 24-71 | 100 | 100 | 200 | | |
| cell Immunogenicity subgroup | None. | 24-71 | 20 | 20 | 40 | Before each dose of Vaccination and on day 14 post Vaccination | Detect the expression levels of Monocytes INF-γ and IL-4. |
| Total | | | **600** | **600** | **1200** | | |

* Additionally, for the humoral immunogenicity subgroup, the first 100 participants with a prior EV-A71 vaccination history and the first 100 participants without prior EV-A71 vaccination history will be selected based on random numbers. Sera collected prior to the first vaccination dose and at Day 30 post completion of the two-dose vaccination schedule from these participants will be tested for CVA10 and CVA6 neutralizing antibodies. Serum specimens obtained from the remaining 960 participants in the humoral immunogenicity subgroup at pre-first vaccination and Day 30 post-second vaccination are tested for neutralizing antibodies against multiple enterovirus subtypes such as EV-A71 and CVA16.

**Table S3. Bivalent Inactivated Enterovirus Vaccine Phase III Clinical Study Reactogenicity Subgroup Study Plan**

| EV-A71 Vaccination history | Month(s) of age group | Bivalent vaccine Group | EV-A71 vaccine Group | Total | Safety observation period | Safety observation method |
|---|---|---|---|---|---|---|
| **None** | 6-23 | 500 | 500 | 1,000 | First Dose Vaccination to 6 months post the last Vaccination | Collect AEs within 30 minutes post each dose of Vaccination, solicited/unsolicited AEs from First Dose Vaccination to 30 days post the last Vaccination (using diary cards/contact cards), and SAEs from First Dose Vaccination to 6 months post the last Vaccination. |
| | 24-71 | 400 | 400 | 800 | | |
| **Yes** | 24-71 | 100 | 100 | 200 | | |
| **Total** | | 1,000 | 1,000 | 2,000 | | |

Note: The non-Reactogenicity subgroup needs to collect AEs within 30 minutes post each dose of Vaccination, and SAEs from First Dose Vaccination to 6 months post the last Vaccination. In addition, AEs reported voluntarily by the participant's guardian or obtained through any other means from the start of First Dose Vaccination to 30 days post the two doses of Vaccination will be noted, accurately recorded, and included in the analysis.

**Table S4. Exclusion Criteria**

| No. | Exclusion Criteria | Assessment Method |
|---|---|---|
| 1 | Known history of hand, foot and mouth disease / herpangina | Inquiry |
| 2 | Presence of uncontrolled chronic diseases or history of serious illnesses, including but not limited to cardiovascular diseases, hematologic diseases, hepatorenal diseases, digestive system diseases, respiratory system diseases, malignant tumors, or major organ transplantation | Inquiry |
| 3 | Presence of autoimmune diseases or immunodeficiency diseases (including but not limited to systemic lupus erythematosus, ankylosing spondylitis, autoimmune thyroid disease, asplenia, functional asplenia, HIV infection) | Inquiry |
| 4 | Presence of coagulation disorders (e.g., clotting factor deficiencies, platelet abnormalities) | Inquiry |
| 5 | History of severe neurological diseases (epilepsy, convulsions or seizures [excluding history of febrile seizures]) or psychiatric disorders, or family history of psychiatric disorders | Inquiry |
| 6 | Acute illness or acute exacerbation of chronic illness within the past 3 days, or known or suspected active infection | Inquiry |
| 7 | Previous vaccination with any CVA16-containing vaccine | Inquiry |
| 8 | Receipt of immunosuppressants or other immunomodulatory therapy for ≥14 days within the past 6 months (prednisone ≥2 mg/kg/day or equivalent; excluding topical or inhaled corticosteroids), cytotoxic therapy, or planned receipt of such therapies during the trial period | Inquiry |
| 9 | Receipt of immunoglobulins or other blood products within the past 6 months, or planned receipt of such products during the trial period | Inquiry |
| 10 | Receipt of other investigational drugs or vaccines within the past 30 days, or planned receipt of such drugs or vaccines during the trial | Inquiry |

| No. | Exclusion Criteria | Assessment Method |
|---|---|---|
| | period | |
| 11 | Receipt of live-attenuated vaccines or nucleic acid vaccines within the past 14 days, or subunit or inactivated vaccines within the past 7 days | Inquiry |
| 12 | Known allergy to any component of the investigational vaccine (inactivated EV-A71 virus, inactivated CVA16 virus, aluminum hydroxide, sodium chloride, disodium hydrogen phosphate, sodium dihydrogen phosphate, water for injection) | Inquiry |
| 13 | Fever on the day of scheduled vaccination, with axillary temperature >37.0°C before vaccination | On-site assessment on the day of vaccination |
| 14 | Unqualified physical examination on the day of scheduled vaccination | On-site assessment on the day of vaccination |
| 15 | Skin lesions, inflammation, ulcers, rashes, or scars at the intended injection site that may interfere with vaccination or local reaction observation | On-site assessment on the day of vaccination |
| 16 | Any other conditions that, in the investigator's judgment, make the participant unsuitable for participation in the clinical trial | Comprehensive investigator judgment |

**Note:** Exclusion criteria No. 1–12 and No. 16 are assessed by inquiry; criteria No. 13–15 are assessed on-site on the day of vaccination through body temperature measurement, physical examination, and injection site inspection.

**Table S5. Basic Information of the Investigational Vaccine**

| Vaccine name | Manufacturer | Dosage | Usage* | Batch Number |
|---|---|---|---|---|
| Bivalent Inactivated Enterovirus Vaccine (Vero cells) high dose | Beijing Sinovac | EV-A71 1920U-CVA16 1200SU/0.5ml | Intramuscular injection into the deltoid muscle of the upper arm | PA20240803V |
| EV-A71 inactivated Vaccine (Vero cells)-Inlive | Beijing Sinovac | 480U/0.5ml specification | Intramuscular injection into the deltoid muscle of the upper arm | HA202311021 |

*:Among participants in the humoral immunogenicity subgroup, 50 eligible participants aged less than 12 months with no prior EV-A71 vaccination history received the full vaccination course via intramuscular injection into the anterolateral thigh muscle. All other participants in the study completed the full vaccination course via intramuscular injection into the deltoid muscle of the upper arm.

**Table S6. Schedule of Visits for Phase III Clinical Study of Bivalent Enterovirus Vaccine (Vero Cell), Inactivated**

| **Planned Visit:** | | **V0[a]** | **V1** | **V2** | **V3[l]** | **V4** | **V5** | **V6[l]** | **V7** | **V8** | **V9** | **V10** | **V11[m]** |
|---|---|---|---|---|---|---|---|---|---|---|---|---|---|
| Follow-up time | D-21 ~ D0 | D-7~D0 | D0 | V1+8d | V1+14d | V1+30d | V4+8d | V4+14d | V4+30d | V4+90d | V4+180d | V4+360d | study End |
| Window period | | | | | +3d | +10d | | +3d | +10d | | +30d | +30d | |
| Preliminary Notice, Participant Recruitment | **X** | | | | | | | | | | | | |
| Informed Consent [b] | | **X** | | | | | | | | | | | |
| Demographics | | **X** | | | | | | | | | | | |
| Medical History and Related Concomitant Medication [c] | | X | X | | | | | | | | | | |
| Physical Examination | | | X | | | | | | | | | | |
| Vital Signs Investigation | | | X | | | X | | | | | | | |
| Inclusion/Exclusion Criteria confirmed[d] | | | X | | | | | | | | | | |
| Randomization | | | X | | | | | | | | | | |
| Vaccination Discontinuation Screening Standards[d] | | | | | | X | | | | | | | |
| Vaccination [e] | | | X | | | X | | | | | | | |
| Body Fluids Immunization Subgroup Blood Collection [f] | | | X | | | | | | X | | X | X | |
| Cell Immunization Subgroup Blood Collection [f] | | | X | | X | X | | X | | | | | |

| | | | | | | | | | | | |
|---|---|---|---|---|---|---|---|---|---|---|---|
| Reactogenicity subgroup: AEs, concomitant medications, and all concomitant vaccines were recorded on diary cards/contact cards [g] | X | X | X | X | X | X | X | | | | |
| All participants: SAEs and concomitant medications associated with SAEs were recorded [h] | X | X | X | X | X | X | X | X | X | | |
| Non-Reactogenicity Subgroup: AEs spontaneously reported by the participant's guardian or obtained through other approaches, as well as concomitant medications and concomitant vaccines, were recorded [i] | X | X | X | X | X | X | X | | | | |
| The usage of all hand, foot, and mouth disease vaccines other than the investigational vaccine was collected [j] | X | X | X | X | X | X | X | X | X | X | X |
| Case monitoring [k] | X | | | | | | X | | | | |

a. Pre-enrollment screening is performed 7 days before vaccination or on the day of vaccination.

b. The Informed Consent Form must be signed prior to the performance of any study-related procedures.

c. Participants' medical history and relevant concomitant medications are inquired before enrollment and on the day of the first vaccination. On the day of the first vaccination, physical examination and vital sign assessment are completed prior to vaccination. On the day before the second vaccination, vital sign assessment is performed to confirm that participants are in good condition for vaccination.

d. Inclusion and exclusion criteria are strictly verified before enrollment. Participants can only be enrolled and receive the first vaccination if all inclusion criteria are met and no exclusion criteria are satisfied. Before the second vaccination, participants are assessed for vaccination discontinuation criteria; the second vaccination can only be administered if no discontinuation criteria are met.

e. All participants undergo a 30-minute on-site observation post each vaccination. Any adverse events occurring during the on-site observation period are recorded in source documents.

f. For the humoral immunogenicity subgroup, venous blood samples are collected using coagulation tubes before the first vaccination, at Day 30 post the completion of two-dose vaccination, at Month 6 post two-dose vaccination, and at Month 12 post two-dose vaccination. For the cellular immunogenicity subgroup, venous blood samples are collected using anticoagulant tubes before each vaccination and at Day 14 post each vaccination. The blood collection volume at each time point is approximately 3.0 mL.

g. For the reactogenicity subgroup, safety observation data are recorded via diary cards from Day 0 to Day 7 post each vaccination, with daily body temperature monitoring. Solicited adverse events (including abnormal vital signs) are recorded in the designated area of the diary card on a daily basis, and any unsolicited adverse events are recorded in a separate area. From Day 8 to Day 30 post each vaccination, adverse events are tracked and recorded via contact cards. Concomitant medications and concomitant vaccines (including HFMD vaccines other than the investigational vaccine and non-HFMD vaccines) used from Day 0 to Day 30 post each vaccination are recorded in diary cards and contact cards. Diary cards are distributed on the day of each vaccination, reviewed and collected on Day 8 post each vaccination, with contact cards issued simultaneously. Contact cards are reviewed and collected on Day 30 post each vaccination.

h. For all participants, serious adverse events (SAEs) and relevant concomitant medications are collected from the first vaccination to 6 months post the second vaccination. For the reactogenicity subgroup, only SAEs are collected from Day 31 post the last vaccination to 6 months post the last vaccination, with dedicated SAE follow-up visits scheduled at 3 months and 6 months post the last vaccination. For the non-reactogenicity subgroup, dedicated SAE follow-up visits are conducted on the day of each vaccination, as well as 3 months and 6 months post the last vaccination.

i. For the non-reactogenicity subgroup, adverse events spontaneously reported by participants' guardians or identified through other active inquiry approaches, as well as corresponding concomitant medications and concomitant vaccines, are recorded from the first vaccination to 30 days post the completion of two-dose vaccination.

j. For all participants, the use of all hand, foot, and mouth disease vaccines other than the investigational vaccine throughout the entire study period is collected and recorded.

k. All participants enter the case surveillance period post completing the first vaccination. The effective surveillance period starts 14 days post the second vaccination and lasts until the end of two consecutive HFMD epidemic seasons. The epidemic season is defined as the annual peak period of HFMD incidence. In this study, each participating study site defines the local provincial HFMD epidemic season based on regional epidemiological characteristics; two incidence peaks within a single year are regarded as one epidemic season. The case surveillance window covers the period from 14 days post the second vaccination onwards. This study adopts combined active and passive case surveillance strategies. During epidemic seasons, surveillance visits are performed approximately once a week; during non-epidemic seasons, surveillance visits are conducted approximately once every two weeks.

l. Only participants in the cellular immunogenicity subgroup are required to complete V3 and V6 study visits.

m. The overall study duration covers the period until the completion of two consecutive HFMD epidemic seasons.

**Table S7. Definitions of epidemic season by various research centers**

| Seasonal Epidemic Period of Hand, Foot and Mouth Disease by Province | |
|---|---|
| Yunnan | May ~ July |
| Anhui | April ~ July |
| Guangdong | April ~ July |
| Hubei | April ~ July |
| Fujian | May ~ July |

**Table S8. Clinical and Laboratory Case Definitions for Enterovirus-Associated Disease**

| Case category | Definition |
|---|---|
| **Suspected enterovirus-associated disease** | Any of the following: (1) rash involving the hands, feet, mouth, pharynx, or buttocks, or fever accompanied by oral ulcers; (2) a clinical diagnosis of hand, foot, and mouth disease (HFMD), herpangina, meningitis, brain-stem encephalitis, encephalitis, Guillain–Barré syndrome, or acute viral myocarditis; or (3) gastrointestinal symptoms, such as abdominal pain or diarrhea, or respiratory symptoms, such as cough or sore throat, together with an outbreak of HFMD in the participant's childcare facility, school, or other close-contact setting, or direct or indirect contact with a child with HFMD before symptom onset. |
| **Separate episodes of suspected enterovirus-associated disease** | Consecutive episodes were considered separate if the onset of symptoms of the subsequent episode occurred ≥5 days after resolution of symptoms of the preceding episode. |
| **HFMD** | Rash involving at least one of the following sites: hands, feet, mouth, or buttocks, with or without fever, cough, rhinorrhea, sore throat, decreased appetite, nausea, vomiting, abdominal pain, or diarrhea. Typical rash consisted of maculopapular lesions, papules, or vesicles, generally with an inflammatory halo and little vesicular fluid and resolving without crusting or scarring. Atypical rash could be small, thick, firm, and sparse and could include petechiae or ecchymoses. |
| **CVA16-associated HFMD** | HFMD meeting the clinical definition above and a positive CVA16 RT-PCR result from a throat swab and/or anal swab. |
| **Herpangina** | Hyperemic red spots or gray-white vesicles on the soft palate, uvula, faucial pillars, or tonsillar surface, which could rupture and form ulcers, with or without fever, pharyngeal erythema or pain, |

| | |
|---|---|
| | drooling, refusal to eat, decreased appetite, irritability or crying, or vomiting. |
| **CVA16-associated herpangina** | Herpangina meeting the clinical definition above and a positive CVA16 RT-PCR result from a throat swab and/or anal swab. |
| **Severe disease** | The presence of one or more of the following: persistent high fever (>39°C) with poor response to conventional antipyretic therapy; neurologic involvement, including brain-stem encephalitis, encephalomyelitis, meningoencephalitis, or acute flaccid paralysis; respiratory abnormalities compatible with neurogenic pulmonary edema; circulatory dysfunction; manifestations suggestive of myocarditis; peripheral white-cell count $\geq 15\times10^9$ /L after exclusion of other infectious causes; stress hyperglycemia with blood glucose >8.3 mmol/L; elevated blood lactate, generally ≥2.0 mmol/L in the presence of circulatory dysfunction; or shock with multiple-organ failure or death. |
| **CVA16-associated severe disease** | Severe disease meeting the clinical definition above and a positive CVA16 RT-PCR result from a throat swab and/or anal swab. |
| **Laboratory confirmation of CVA16 infection** | Local laboratories performed initial RT-PCR testing using a five-target enterovirus assay for EV-G, CVA16, CVA10, CVA6, and EV-A71. A sampling visit was considered positive if either the throat swab or anal swab was positive. For participants with at least one CVA16-positive sample, all swab specimens collected during that illness episode were retested by the National Institutes for Food and Drug Control (NIFDC). If local and NIFDC results were discordant, the NIFDC result prevailed. |

| | |
|---|---|
| **Laboratory-confirmed CVA16 case for the primary efficacy analysis** | A clinically compatible CVA16-associated disease with CVA16-positive RT-PCR results from at least two sampling visits. Cases with only one CVA16-positive sampling visit were included in supplementary analyses but not in the primary efficacy analysis. |
| **Clinical end-point adjudication** | A CVA16-associated case was required to meet both the prespecified clinical and laboratory criteria. The Clinical Event Adjudication Committee (CEAC) reviewed the clinical manifestations and laboratory results of all suspected cases and made the final determination as to whether each case qualified as a clinical end-point event. |

**Table S9. Adverse Event Grading Table**

| Symptoms | Grade 1 | Grade 2 | Grade 3 | Grade 4 |
|---|---|---|---|---|
| Injection Site | | | | |
| Tenderness | Resistance or withdrawal upon contact or touch | Crying or fussing post contact or touch, but consolable | Persistent crying and agitation that is inconsolable | Requiring emergency intervention or hospitalization |
| Pain | Does not affect or slightly affects limb movement. | Interference with limb movement | Interference with daily activities | Loss of basic self-care ability or requiring hospitalization |
| Induration*# | Diameter < 2.5 cm | Diameter ≥ 2.5 cm and lesion area < 50% of the vaccinated limb (anatomical limb containing the injection site, such as the upper arm or thigh) | Lesion area ≥ 50% of the vaccinated limb; or presence of ulceration, secondary infection, phlebitis, or wound drainage | Abscess, exfoliative dermatitis, or dermal/deep tissue necrosis |
| Swelling | Diameter < 2.5 cm | Diameter ≥ 2.5 cm and lesion area < 50% of the vaccinated limb (anatomical limb containing the injection site, such as upper arm or thigh) | Lesion area ≥ 50% of the vaccinated limb; or presence of ulceration, secondary infection, phlebitis, or wound drainage | Abscess, exfoliative dermatitis, or dermal/deep tissue necrosis |
| Erythema | Diameter < 2.5 cm | Diameter ≥ 2.5 cm and lesion area < 50% of the vaccinated limb (anatomical limb containing the injection site, such as upper arm or thigh) | Lesion area ≥ 50% of the vaccinated limb; or presence of ulceration, secondary infection, phlebitis, or wound drainage | Abscess, exfoliative dermatitis, or dermal/deep tissue necrosis |
| Rash*# | Diameter < 2.5 cm | Diameter ≥ 2.5 cm and lesion area < 50% of the vaccinated limb (anatomical limb containing the injection site, such as upper arm or thigh) | Lesion area ≥ 50% of the vaccinated limb; or presence of ulceration, secondary infection, phlebitis, or wound drainage | Abscess, exfoliative dermatitis, or dermal/deep tissue necrosis |

| Symptoms | Grade 1 | Grade 2 | Grade 3 | Grade 4 |
|---|---|---|---|---|
| Itching | Injection site Itching resolving spontaneously or within 48 hours post symptomatic treatment | Injection site Itching unrelieved within 48 hours post symptomatic treatment | Interference with daily activities | Not applicable (NA) |
| Non-Injection Site | | | | |
| Acute allergic reaction** | Localized urticaria (blister) without treatment required | Localized urticaria requiring treatment, or mild angioedema without treatment required | Extensive urticaria or angioedema requiring treatment, or mild bronchospasm | Anaphylactic shock, or life-threatening bronchospasm or laryngeal edema |
| Loss of appetite | Decreased appetite without reduction in food intake | Decreased appetite with reduced oral intake but no significant weight loss | Decreased appetite accompanied by significant weight loss | Requiring nutritional intervention measures, such as nasogastric tube feeding or parenteral nutrition |
| Vomiting | 1–2 episodes per 24 hours without activity impairment | 3–5 episodes per 24 hours or limited daily activity | More than 6 episodes per 24 hours or requiring intravenous fluid replacement | Requiring hospitalization due to hypotensive shock or mandatory nutritional support |
| Irritability | Mild irritability, no behavioral disturbance | Moderate irritability with obvious behavioral disturbance | Severe, inconsolable irritability | Not applicable (NA) |
| Decreased activity | Mild reduction in activity level with normal responsiveness | Moderate somnolence, arousable with stimulation | Severe poor responsiveness, difficult to arouse | Not applicable (NA) |
| Diarrhea | Mild or transient diarrhea, 3–4 episodes daily with abnormal stool consistency, or mild diarrhea lasting less than 1 week | Moderate or persistent diarrhea, 5–7 episodes daily with abnormal stool consistency, or diarrhea lasting more than 1 week | More than 7 episodes daily with abnormal stool consistency, or hemorrhagic diarrhea, orthostatic hypotension, electrolyte imbalance requiring intravenous infusion | Hypotensive shock requiring inpatient hospitalization |

| Symptoms | Grade 1 | Grade 2 | Grade 3 | Grade 4 |
|---|---|---|---|---|
| | | | exceeding 2 L | |
| Nausea | Transient (<24 hours) or intermittent nausea with nearly normal oral food intake | Persistent nausea resulting in reduced oral intake for 24–48 hours | Persistent nausea resulting in negligible oral intake for more than 48 hours or requiring intravenous fluid resuscitation | Life-threatening conditions, such as hypotensive shock |
| Fatigue | No impairment of daily activities | Mild impairment of routine daily activities | Severe impairment of daily activities and inability to perform routine tasks | Requiring emergency intervention or inpatient hospitalization |
| Vital Signs | | | | |
| Fever [axillary temperature ℃] | 37.5 to <38.0 | 38.0 to <39.5 | ≥39.5 | ≥39.5 ℃ and persistent for more than 5 consecutive days |

*Note: In addition to grading and evaluating Induration and Rash by directly measuring their diameters, the progression of changes in the measurement results should also be recorded.

#Induration and Swelling, Rash and Erythema should use the maximum measured diameter or area; evaluation and grading should be based on functional grade and actual measurement results, selecting the indicator with the higher grade.

**Refers to Type I Hypersensitivity. Given the immediate-onset characteristics of Type I Hypersensitivity [24], solicitation is conducted only within 48 hours post each dose of Vaccination.

**Table S10. Vaccine Efficacy against Hand-foot-and-mouth disease caused by CVA16 Infection confirmed by NIFDC RT-PCR in the overall population from Day 14 post full-course Immunization - based on Poisson regression model (E-PPS)**

| | Bivalent vaccine group (N=3560) | | | | EV-A71 vaccine group (N=3586) | | | | | |
|---|---|---|---|---|---|---|---|---|---|---|
| | **n[1]** | **Number of cases** | **Person-years of exposure** | **Annual incidence rate per person-year (%, 95% CI)[2]** | **n[1]** | **Number of cases** | **Person-years of exposure** | **Annual incidence rate per person-year (%, 95% CI)[2]** | **Protection rate (%, 95% CI)[3]** | ***P* value [3]** |
| 6-71 Month(s) of age | 3,560 | 4 | 2,962.6 | 0.14 (0.05, 0.36) | 3,586 | 43 | 2,961.9 | 1.45 (1.08, 1.96) | 90.68 (74.36, 97.57) | <0.0001 |
| Age Group | | | | | | | | | | |
| 6-23 Month(s) of age | 1,780 | 3 | 1,472.4 | 0.20 (0.07, 0.63) | 1,795 | 18 | 1,477.2 | 1.22 (0.77, 1.93) | 83.28 (43.23, 95.07) | 0.0041 |
| 24-71 Month(s) of age | 1,780 | 1 | 1,490.2 | 0.07 (0.01, 0.48) | 1,791 | 25 | 1,484.7 | 1.68 (1.14, 2.49) | 96.00 (75.56, 99.90) | <0.0001 |
| History of EV-A71 Vaccination | | | | | | | | | | |
| Yes | 191 | 0 | 165.9 | 0.00 (0.00, 2.22) | 183 | 1 | 157.3 | 0.64 (0.02, 3.54) | 100.00 (-3,598.72, 100.00) | 0.9735 |
| None | 3,369 | 4 | 2,796.7 | 0.14 (0.05, 0.38) | 3403 | 42 | 2,804.6 | 1.50 (1.11, 2.03) | 90.45 (65.61, 97.35) | 0.0003 |
| 24-71 Month(s) with no EV-A71 vaccination history | 1,589 | 1 | 1,324.3 | 0.08 (0.01, 0.54) | 1608 | 24 | 1,327.4 | 1.81 (1.21, 2.70) | 95.82 (69.13, 99.44) | 0.0019 |

[1] n: Number of evaluable cases.

[2] A Poisson regression model that does not account for stratification factors was used to calculate the incidence rates per person-year for each group and their 95% confidence intervals.[3] A Poisson regression model that accounts for stratification factors was used to calculate the vaccine protection rate and its 95% confidence interval. In these models, the number of cases served as the dependent variable, with age groups (6–23 months, 24–71 months), EV-A71 vaccination history (yes/no), and group assignment as fixed effects, and the logarithm of participant exposure-person-years as the offset, using the log link function. If the number of cases at any stratification level in either the treatment or EV-A71 vaccine Group is 0, the 95% confidence interval for the protection rate is calculated using the exact method that accounts for stratification factors in StatXact software. (For age group analysis, EV-A71 vaccination history and group are treated as fixed effects; for EV-A71 vaccination history analysis, age group and group are treated as fixed effects.)

(1) Subject R22186 in the non-reactivity subgroup, who was randomly assigned to the treatment group but mistakenly received two doses of the control vaccine, will be analyzed as part of the treatment group in the efficacy evaluation analysis set.

(2)Data analysis cutoff date: 2025-12-31.

(3)Bivalent vaccine: EV-A71-CVA16 vaccine (EV-A71, Enterovirus 71; CVA16, Coxsackievirus A16).

(4)95% CI, 95% confidence interval.

#### Table S11. Vaccine Efficacy against CVA16 Infection Confirmed by NIFDC RT-PCR in the Overall Population 14 Days post Full Immunization – Based on Poisson Regression Model (E-mFAS)

| | Bivalent vaccine group (N=3,699) | | | | EV-A71 vaccine group (N=3,720) | | | | | |
|---|---|---|---|---|---|---|---|---|---|---|
| | n[1] | Number of cases | Person-years of exposure | Person-year incidence rate (%, 95% CI)[2] | n[1] | Number of cases | Person-years of exposure | Person-year incidence rate (%, 95% CI)[2] | Protection rate (%, 95% CI)[3] | *P* value [3] |
| Herpangina caused by CVA16 infection | | | | | | | | | | |
| 6-71 Month(s) of age | 3,696 | 0 | 3,063.3 | 0.00 (0.00, 0.12) | 3,717 | 5 | 3,058.1 | 0.16 (0.05, 0.38) | 100.00 (-9.15, 100.00) | 0.0625 |
| Age Group | | | | | | | | | | |
| 6-23 Month(s) of age | 1,850 | 0 | 1,522.7 | 0.00 (0.00, 0.24) | 1,862 | 1 | 1,525.6 | 0.07 (0.00, 0.37) | 100.00 (-3,807.29, 100.00) | 1.0000 |
| 24-71 Month(s) of age | 1,846 | 0 | 1,540.6 | 0.00 (0.00, 0.24) | 1,855 | 4 | 1,532.5 | 0.26 (0.07, 0.67) | 100.00 (-51.46, 100.00) | 0.1249 |
| History of EV-A71 Vaccination | | | | | | | | | | |
| Yes | 199 | 0 | 172.1 | 0.00 (0.00, 2.14) | 192 | 0 | 164.3 | 0.00 (0.00, 2.24) | NE | NE |
| None | 3,497 | 0 | 2,891.2 | 0.00 (0.00, 0.13) | 3,525 | 5 | 2,893.7 | 0.17 (0.06, 0.40) | 100.00 (-9.15, 100.00) | 0.0625 |
| 24-71 Month(s) of age | 1,647 | 0 | 1,368.4 | 0.00 (0.00, 0.27) | 1,663 | 4 | 1,368.2 | 0.29 (0.08, 0.75) | 100.00 (-51.46, 100.00) | 0.1249 |
| Diseases Related to CVA16 Infection | | | | | | | | | | |
| 6-71 Month(s) of age | 3,696 | 6 | 3,063.3 | 0.20 (0.09, 0.44) | 3,717 | 49 | 3,058.1 | 1.60 (1.21, 2.12) | 87.76 (71.39, 95.71) | <0.0001 |
| Age Group | | | | | | | | | | |
| 6-23 Month(s) of age | 1,850 | 3 | 1,522.7 | 0.20 (0.06, 0.61) | 1,862 | 20 | 1,525.6 | 1.31 (0.85, 2.03) | 84.97 (49.43, 95.53) | 0.0022 |
| 24-71 Month(s) of age | 1,846 | 3 | 1,540.6 | 0.19 (0.06, 0.60) | 1,855 | 29 | 1,532.5 | 1.89 (1.32, 2.72) | 89.67 (66.68, 97.99) | <0.0001 |
| History of EV-A71 Vaccination | | | | | | | | | | |
| Yes | 199 | 0 | 172.1 | 0.00 (0.00, 2.14) | 192 | 1 | 164.3 | 0.61 (0.02, 3.39) | 100.00 (-3,623.64, 100.00) | 0.9769 |
| None | 3,497 | 6 | 2,891.2 | 0.21 (0.09, 0.46) | 3,525 | 48 | 2,893.7 | 1.66 (1.25, 2.20) | 87.49 (82.59, 91.01) | <0.0001 |

| | Bivalent vaccine group (N=3,699) | | | | EV-A71 vaccine group (N=3,720) | | | | | |
|---|---|---|---|---|---|---|---|---|---|---|
| | n[1] | Number of cases | Person-years of exposure | Person-year incidence rate (%, 95% CI)[2] | n[1] | Number of cases | Person-years of exposure | Person-year incidence rate (%, 95% CI)[2] | Protection rate (%, 95% CI)[3] | *P* value [3] |
| 24-71 Month(s) of age | 1,647 | 3 | 1,368.4 | 0.22 (0.07, 0.68) | 1,663 | 28 | 1,368.2 | 2.05 (1.41, 2.96) | 89.29 (64.77, 96.74) | 0.0002 |
| Severe cases caused by CVA16 infection | | | | | | | | | | |
| 6-71 Month(s) of age | 3,696 | 0 | 3,063.3 | 0.00 (0.00, 0.12) | 3,717 | 0 | 3,058.1 | 0.00 (0.00, 0.12) | NE | NE |
| Age Group | | | | | | | | | | |
| 6-23 Month(s) of age | 1,850 | 0 | 1,522.7 | 0.00 (0.00, 0.24) | 1,862 | 0 | 1,525.6 | 0.00 (0.00, 0.24) | NE | NE |
| 24-71 Month(s) of age | 1,846 | 0 | 1,540.6 | 0.00 (0.00, 0.24) | 1,855 | 0 | 1,532.5 | 0.00 (0.00, 0.24) | NE | NE |
| History of EV-A71 Vaccination | | | | | | | | | | |
| Yes | 199 | 0 | 172.1 | 0.00 (0.00, 2.14) | 192 | 0 | 164.3 | 0.00 (0.00, 2.24) | NE | NE |
| None | 3,497 | 0 | 2,891.2 | 0.00 (0.00, 0.13) | 3,525 | 0 | 2,893.7 | 0.00 (0.00, 0.13) | NE | NE |
| 24-71 Month(s) of age | 1,647 | 0 | 1,368.4 | 0.00 (0.00, 0.27) | 1,663 | 0 | 1,368.2 | 0.00 (0.00, 0.27) | NE | NE |
| Hospitalization cases caused by CVA16 infection | | | | | | | | | | |
| 6-71 Month(s) of age | 3,696 | 0 | 3,063.3 | 0.00 (0.00, 0.12) | 3,717 | 6 | 3,058.1 | 0.20 (0.07, 0.43) | 100.00 (14.91, 100.00) | 0.0314 |
| Age Group | | | | | | | | | | |
| 6-23 Month(s) of age | 1,850 | 0 | 1,522.7 | 0.00 (0.00, 0.24) | 1,862 | 6 | 1,525.6 | 0.39 (0.14, 0.86) | 100.00 (14.91, 100.00) | 0.0314 |
| 24-71 Month(s) of age | 1,846 | 0 | 1,540.6 | 0.00 (0.00, 0.24) | 1,855 | 0 | 1,532.5 | 0.00 (0.00, 0.24) | NE | NE |
| History of EV-A71 Vaccination | | | | | | | | | | |
| Yes | 199 | 0 | 172.1 | 0.00 (0.00, 2.14) | 192 | 0 | 164.3 | 0.00 (0.00, 2.24) | NE | NE |
| None | 3,497 | 0 | 2,891.2 | 0.00 (0.00, 0.13) | 3,525 | 6 | 2,893.7 | 0.21 (0.08, 0.45) | 100.00 (14.91, 100.00) | 0.0314 |

| | Bivalent vaccine group (N=3,699) | | | | EV-A71 vaccine group (N=3,720) | | | | | |
|---|---|---|---|---|---|---|---|---|---|---|
| | **n[1]** | **Number of cases** | **Person-years of exposure** | **Person-year incidence rate (%, 95% CI)[2]** | **n[1]** | **Number of cases** | **Person-years of exposure** | **Person-year incidence rate (%, 95% CI)[2]** | **Protection rate (%, 95% CI)[3]** | ***P* value [3]** |
| 24-71 Month(s) of age | 1,647 | 0 | 1,368.4 | 0.00 (0.00, 0.27) | 1,663 | 0 | 1,368.2 | 0.00 (0.00, 0.27) | NE | NE |

[1] n: Number of evaluable cases. Six participants (3 in the Bivalent vaccine Group and 3 in the EV-A71 vaccine Group) used prohibited medications prior to the valid case monitoring period and were excluded from evaluable cases.

[2] The Poisson regression model without stratification factors was used to calculate the person-year incidence rate and its 95% confidence interval of each group.

[3] The Poisson regression model with stratification factors was used to calculate the vaccine protection rate and its 95% confidence interval. In the model, the number of onset cases served as the dependent variable. Age group (6-23 months, 24-71 months), EV-A71 vaccination history (Yes, No) and group were set as fixed effects. The logarithm of participant exposure person-years was taken as the offset, and the log link function was applied. When the number of cases was zero at any stratification level in the Bivalent vaccine Group or EV-A71 vaccine Group, the exact method considering stratification factors in StatXact software was adopted to calculate the 95% confidence interval of the protection rate. For age group analysis, EV-A71 vaccination history and group were defined as fixed effects. For EV-A71 vaccination history analysis, age group and group were defined as fixed effects.

(1) Participant with random number R22186 in the non-reactogenicity subgroup was scheduled to receive treatment vaccine but erroneously administered two doses of control vaccine, and was analyzed as part of the Bivalent vaccine Group in the vaccine efficacy analysis set.

(2) Data analysis cutoff date: 2025-12-31.

(3)Bivalent vaccine: EV-A71-CVA16 vaccine (EV-A71, Enterovirus 71; CVA16, Coxsackievirus A16).

(4)95% CI, 95% confidence interval.

**Table S12. Vaccine Efficacy Based on Incidence Rate (E-mFAS) for CVA10, CVA6, and Other Enterovirus Infection Cases Confirmed by Local Laboratory RT-PCR in the Overall Population 14 Days Post Full Immunization**

| | Bivalent vaccine group(N=3,700) | | | EV-A71 vaccine group(N=3,720) | | | | |
|---|---|---|---|---|---|---|---|---|
| | n[1] | Number of cases | Incidence rate (%, 95% CI) | n[1] | Number of cases | Incidence rate (%, 95% CI) | Protection rate (%, 95% CI)[2] | *P* value [2] |
| CVA10 Infection cases | | | | | | | | |
| 6-71 Month(s) of age | 3,697 | 12 | 0.32 (0.17, 0.57) | 3,717 | 17 | 0.46 (0.27, 0.73) | 29.05 (-48.20, 66.03) | 0.3588 |
| Age Group | | | | | | | | |
| 6-23 Month(s) of age | 1,850 | 6 | 0.32 (0.12, 0.70) | 1,863 | 8 | 0.43 (0.19, 0.84) | 24.47 (-117.25, 73.74) | 0.6014 |
| 24-71 Month(s) of age | 1,847 | 6 | 0.32 (0.12, 0.71) | 1,854 | 9 | 0.49 (0.22, 0.92) | 33.10 (-87.09, 76.08) | 0.4405 |
| History of EV-A71 Vaccination | | | | | | | | |
| Yes | 200 | 0 | 0.00 (0.00, 1.83) | 192 | 2 | 1.04 (0.13, 3.71) | 100.00 (-410.69, 100.00) | 0.1484 |
| None. | 3,497 | 12 | 0.34 (0.18, 0.60) | 3,525 | 15 | 0.43 (0.24, 0.70) | 19.36 (-72.03, 62.20) | 0.5771 |
| 24-71 Month(s) of age | 1,647 | 6 | 0.36 (0.13, 0.79) | 1,662 | 7 | 0.42 (0.17, 0.87) | 13.51 (-156.82, 70.87) | 0.7937 |
| CVA6 Infection cases | | | | | | | | |
| 6-71 Month(s) of age | 3,697 | 226 | 6.11 (5.36, 6.93) | 3,716 | 280 | 7.53 (6.71, 8.43) | 18.81 (3.89, 31.40) | 0.0152 |
| Age Group | | | | | | | | |
| 6-23 Month(s) of age | 1,850 | 143 | 7.73 (6.55, 9.04) | 1,861 | 191 | 10.26 (8.92, 11.73) | 24.69 (7.37, 38.77) | 0.0070 |
| 24-71 Month(s) of age | 1,847 | 83 | 4.49 (3.59, 5.54) | 1,855 | 89 | 4.80 (3.87, 5.87) | 6.19 (-25.60, 29.94) | 0.6678 |
| History of EV-A71 Vaccination | | | | | | | | |
| Yes | 200 | 5 | 2.50 (0.82, 5.74) | 192 | 8 | 4.17 (1.82, 8.04) | 40.00 (-80.19, 80.02) | 0.3575 |
| None. | 3,497 | 221 | 6.32 (5.54, 7.18) | 3,524 | 272 | 7.72 (6.86, 8.65) | 18.17 (2.94, 31.01) | 0.0210 |
| 24-71 Month(s) of age | 1,647 | 78 | 4.74 (3.76, 5.88) | 1,663 | 81 | 4.87 (3.89, 6.02) | 2.77 (-31.69, 28.21) | 0.8561 |
| Other Enterovirus infection cases | | | | | | | | |
| 6-71 Month(s) of age | 3,689 | 82 | 2.22 (1.77, 2.75) | 3,710 | 90 | 2.43 (1.96, 2.97) | 8.29 (-23.24, 31.75) | 0.5659 |
| Age Group | | | | | | | | |
| 6-23 Month(s) of age | 1,845 | 48 | 2.60 (1.92, 3.43) | 1,858 | 50 | 2.69 (2.00, 3.53) | 3.32 (-42.90, 34.60) | 0.8654 |
| 24-71 Month(s) of age | 1,844 | 34 | 1.84 (1.28, 2.57) | 1,852 | 40 | 2.16 (1.55, 2.93) | 14.48 (-34.39, 45.58) | 0.4971 |

| | Bivalent vaccine group(N=3,700) | | | EV-A71 vaccine group(N=3,720) | | | | |
|---|---|---|---|---|---|---|---|---|
| | n[1] | Number of cases | Incidence rate (%, 95% CI) | n[1] | Number of cases | Incidence rate (%, 95% CI) | Protection rate (%, 95% CI)[2] | *P* value [2] |
| History of EV-A71 Vaccination | | | | | | | | |
| Yes | 200 | 1 | 0.50 (0.01, 2.75) | 190 | 5 | 2.63 (0.86, 6.03) | 81.00 (-61.14, 97.76) | 0.0878 |
| None. | 3,489 | 81 | 2.32 (1.85, 2.88) | 3,520 | 85 | 2.41 (1.93, 2.98) | 3.88 (-29.83, 28.84) | 0.7963 |
| 24-71 Month(s) of age | 1,644 | 33 | 2.01 (1.39, 2.81) | 1,662 | 35 | 2.11 (1.47, 2.92) | 4.68 (-52.61, 40.46) | 0.8417 |

[1] n: Number of evaluable cases. 6 participants (3 in Bivalent vaccine Group, 3 in EV-A71 vaccine Group). Use of prohibited medications before the effective case monitoring period will result in exclusion from the evaluable case count.

[2] CMH- $\chi$ 2 method incorporating stratification factors including Age Group (6-23 months, 24-71 months) and EV-A71 Vaccination history (Yes, No) was adopted to calculate protection rate and P value. For Age Group analysis, EV-A71 Vaccination history served as the stratification factor; for EV-A71 Vaccination history analysis, Age Group served as the stratification factor.

(1) Participant with randomization number R22186 in Non-Reactogenicity subgroup was assigned to Bivalent vaccine Group, erroneously received two doses of EV-A71 vaccine Group Vaccine, and was analyzed as Bivalent vaccine Group in the Vaccine Efficacy Evaluation Analysis Set.

(2) Data analysis cutoff Date: 2025-12-31.

(3)Bivalent vaccine: EV-A71-CVA16 vaccine (EV-A71, Enterovirus 71; CVA16, Coxsackievirus A16).

(4)95% CI, 95% confidence interval.

**Table S13. Pre-vaccination and 30 days post-full vaccination Neutralizing antibodies levels by type_EV-A71, CVA16, CVA6 and CVA10 types_6-71 Month(s) of age (HI-mFAS/IPS-6)**

| | Bivalent vaccine group with history of EV-A71 vaccination | Bivalent vaccine group with no history of EV-A71 vaccination | Total bivalent vaccine group | EV-A71 vaccine group with History of EV-A71 vaccination | EV-A71 vaccine group with No history of EV-A71 vaccination | Total EV-A71 vaccine group |
|---|---|---|---|---|---|---|
| **EV-A71** | | | | | | |
| **Pre-free** | | | | | | |
| n (Missing) | 99 (0) | 447 (3) | 546 (3) | 91 (0) | 463 (2) | 554 (2) |
| Positive rate n(%) | 97 (97.98) | 7 (1.57) | 104 (19.05) | 89 (97.80) | 13 (2.81) | 102 (18.41) |
| 95% CI(%) | (92.89, 99.75) | (0.63, 3.20) | (15.84, 22.60) | (92.29, 99.73) | (1.50, 4.75) | (15.27, 21.89) |
| GMT | 204.50 | 1.05 | 2.74 | 201.38 | 1.13 | 2.64 |
| 95% CI | (149.16, 280.37) | (1.01, 1.10) | (2.28, 3.29) | (148.55, 272.99) | (1.05, 1.21) | (2.21, 3.16) |
| **30 days post full-course Vaccination** | | | | | | |
| n(Missing) | 99(0) | 450(0) | 549(0) | 91(0) | 465(0) | 556(0) |
| Positive rate (%) | 100.00 | 100.00 | 100.00 | 100.00 | 99.35 | 99.46 |
| 95% CI (%) | (96.34,100.00) | (99.18,100.00) | (99.33,100.00) | (96.03,100.00) | (98.13, 99.87) | (98.43,99.89) |
| GMT | 20,704.83 | 1,193.45 | 1,996.54 | 13,561.16 | 591.45 | 987.58 |
| 95% CI | (15,823.89, 27,091.33) | (1,065.36,1,336.94) | (1,737.05, 2,294.80) | (9,656.13, 19,045.43) | (517.18, 676.39) | (841.87,1,158.49) |
| n(Missing) | 99(0) | 447(3) | 546(3) | 91(0) | 463(2) | 554(2) |
| Seroconversion rate (%) | 95.96 | 100.00 | 99.27 | 94.07 | 98.70 | 97.94 |
| 95% CI (%) | (89.98,98.89) | (99.18,100.00) | (98.13,99.80) | (88.61,99.52) | (97.20,99.52) | (96.69, 99.19) |
| GMFR | 101.25 | 1,129.18 | 729.21 | 67.34 | 521.76 | 372.75 |
| 95% CI | (72.61,141.17) | (1,004.63, 1,269.18) | (635.77,836.38) | (46.82, 96.85) | (452.60, 601.49) | (321.15,432.62) |
| **6 months post full-course Vaccination** | | | | | | |
| n(Missing) | 97 (0) | 413 (0) | 510 (0) | 87 (0) | 433 (0) | 520 (0) |
| Positive rate (%) | 97 (100.00) | 412 (99.76) | 509 (99.80) | 87 (100.00) | 420 (97.00) | 507 (97.50) |
| 95% CI (%) | (96.27, 100.00) | (98.66, 99.99) | (98.91, 100.00) | (95.85, 100.00) | (94.92, 98.39) | (95.76, 98.66) |
| GMT | 4,939.24 | 256.37 | 450.01 | 4,424.09 | 115.92 | 213.20 |

| | Bivalent vaccine group with history of EV-A71 vaccination | Bivalent vaccine group with no history of EV-A71 vaccination | Total bivalent vaccine group | EV-A71 vaccine group with History of EV-A71 vaccination | EV-A71 vaccine group with No history of EV-A71 vaccination | Total EV-A71 vaccine group |
|---|---|---|---|---|---|---|
| 95% CI | (3,660.07, 6,665.46) | (229.10, 286.88) | (388.43, 521.36) | (3,260.42, 6,003.07) | (101.51, 132.37) | (180.10, 252.38) |
| **CVA16** | | | | | | |
| **Pre-free** | | | | | | |
| n (Missing) | 98 (0) | 447 (3) | 545 (3) | 91 (0) | 463 (2) | 554 (2) |
| Positive rate n(%) | 31 (31.63) | 104 (23.27) | 135 (24.77) | 35 (38.46) | 99 (21.38) | 134 (24.19) |
| 95% CI (%) | (22.61, 41.80) | (19.43, 27.46) | (21.20, 28.62) | (28.45, 49.25) | (17.73, 25.40) | (20.68, 27.98) |
| GMT | 5.01 | 3.20 | 3.47 | 7.13 | 2.88 | 3.34 |
| 95% CI | (3.00, 8.35) | (2.59, 3.95) | (2.85, 4.22) | (4.09, 12.43) | (2.36, 3.50) | (2.76, 4.04) |
| **30 days post full-course Vaccination** | | | | | | |
| n(Missing) | 98(0) | 450(0) | 548(0) | 91(0) | 465(0) | 556(0) |
| Positive rate (%) | 100.00 | 100.00 | 100.00 | 44.73 | 22.88 | 26.46 |
| 95% CI (%) | (96.31,100.00) | (99.18,100.00) | (99.33,100.00) | (34.01,55.44) | (18.91, 26.85) | (22.64,30.28) |
| GMT | 1,468.68 | 829.76 | 918.96 | 9.41 | 3.26 | 3.88 |
| 95% CI | (1,169.84, 1,843.87) | (730.50, 942.50) | (820.29, 1,029.51) | (5.20,17.03) | (2.60, 4.09) | (3.12, 4.82) |
| n(Missing) | 98(0) | 447(3) | 545(3) | 91(0) | 463(2) | 554(2) |
| Seroconversion rate (%) | 96.94 | 96.94 | 96.94 | 11.10 | 8.21 | 8.68 |
| 95% CI (%) | (91.31, 99.36) | (95.26,98.61) | (95.43,98.44) | (4.26, 17.94) | (5.52,10.90) | (6.15, 11.21) |
| GMFR | 293.22 | 258.83 | 264.70 | 1.32 | 1.14 | 1.17 |
| 95% CI | (195.62, 439.54) | (217.10, 308.58) | (225.25,311.05) | (0.95,1.85) | (1.03,1.26) | (1.05,1.30) |
| **6 months post full-course Vaccination** | | | | | | |
| n(Missing) | 97 (0) | 413 (0) | 510 (0) | 87 (0) | 433 (0) | 520 (0) |
| Positive rate (%) | 97 (100.00) | 413 (100.00) | 510 (100.00) | 39 (44.83) | 106 (24.48) | 145 (27.88) |
| 95% CI (%) | (96.27, 100.00) | (99.11, 100.00) | (99.28, 100.00) | (34.15, 55.87) | (20.50, 28.81) | (24.07, 31.95) |
| GMT | 426.45 | 225.94 | 254.95 | 14.15 | 3.96 | 4.90 |
| 95% CI | (311.23, 584.33) | (190.76, 267.61) | (219.28, 296.43) | (7.19, 27.83) | (3.11, 5.03) | (3.88, 6.18) |
| **CVA10** | | | | | | |

| | Bivalent vaccine group with history of EV-A71 vaccination | Bivalent vaccine group with no history of EV-A71 vaccination | Total bivalent vaccine group | EV-A71 vaccine group with History of EV-A71 vaccination | EV-A71 vaccine group with No history of EV-A71 vaccination | Total EV-A71 vaccine group |
|---|---|---|---|---|---|---|
| **Pre-free** | | | | | | |
| n (Missing) | 47 (0) | 46 (0) | 93 (0) | 46 (0) | 47 (0) | 93 (0) |
| Positive rate n(%) | 23 (48.94) | 13 (28.26) | 36 (38.71) | 25 (54.35) | 14 (29.79) | 39 (41.94) |
| 95% CI (%) | (34.08, 63.94) | (15.99, 43.46) | (28.78, 49.38) | (39.01, 69.10) | (17.34, 44.89) | (31.78, 52.62) |
| GMT | 9.98 | 2.22 | 4.74 | 11.70 | 2.05 | 4.86 |
| 95% CI | (4.41, 22.56) | (1.38, 3.58) | (2.90, 7.76) | (4.83, 28.34) | (1.32, 3.20) | (2.90, 8.12) |
| **30 days post full-course Vaccination** | | | | | | |
| n (Missing) | 47(0) | 46(0) | 93(0) | 46(0) | 47(0) | 93(0) |
| Positive rate (%) | 59.57 | 40.87 | 50.32 | 54.35 | 18.30 | 36.13 |
| 95% CI (%) | (44.27,73.63) | (26.54,55.20) | (40.12,60.53) | (39.01, 69.10) | (7.02,29.58) | (26.30,45.96) |
| Seroconversion rate (%) | 25.53 | 23.48 | 24.52 | 8.70 | 11.91 | 10.32 |
| 95% CI (%) | (13.94,40.35) | (11.09, 35.87) | (15.72,33.31) | (2.42, 20.79) | (2.38,21.45) | (4.03,16.61) |
| GMT | 16.21 | 3.55 | 7.64 | 10.90 | 2.04 | 4.67 |
| 95% CI | (6.94, 37.86) | (1.99,6.32) | (4.51,12.94) | (4.56,26.11) | (1.13,3.68) | (2.72,8.03) |
| GMFR | 1.62 | 1.60 | 1.61 | 0.93 | 0.99 | 0.96 |
| 95% CI | (1.14, 2.31) | (1.09,2.34) | (1.25, 2.08) | (0.63,1.39) | (0.68, 1.46) | (0.73,1.26) |
| **CVA6** | | | | | | |
| **Pre-free** | | | | | | |
| n (Missing) | 47 (0) | 46 (0) | 93 (0) | 46 (0) | 47 (0) | 93 (0) |
| Positive rate n(%) | 21 (44.68) | 7 (15.22) | 28 (30.11) | 21 (45.65) | 4 (8.51) | 25 (26.88) |
| 95% CI (%) | (30.17, 59.88) | (6.34, 28.87) | (21.03, 40.50) | (30.90, 60.99) | (2.37, 20.38) | (18.21, 37.08) |
| GMT | 17.52 | 2.51 | 6.71 | 27.95 | 1.57 | 6.53 |
| 95% CI | (6.32, 48.61) | (1.21, 5.23) | (3.50, 12.85) | (8.88, 87.98) | (0.94, 2.64) | (3.30, 12.91) |
| **30 days post full-course Vaccination** | | | | | | |
| n (Missing) | 47(0) | 46(0) | 93(0) | 46(0) | 47(0) | 93(0) |
| Positive rate (%) | 46.81 | 18.91 | 33.01 | 45.65 | 6.38 | 26.45 |

| | Bivalent vaccine group with history of EV-A71 vaccination | Bivalent vaccine group with no history of EV-A71 vaccination | Total bivalent vaccine group | EV-A71 vaccine group with History of EV-A71 vaccination | EV-A71 vaccine group with No history of EV-A71 vaccination | Total EV-A71 vaccine group |
|---|---|---|---|---|---|---|
| 95% CI (%) | (32.11,61.92) | (7.39,30.43) | (23.39, 42.63) | (30.90, 60.99) | (0.00,17.02) | (17.42,35.49) |
| Seroconversion rate (%) | 8.51 | 12.39 | 10.43 | 2.17 | 4.26 | 2.15 |
| 95% CI (%) | (2.37,20.38) | (2.63,22.15) | (4.13, 16.73) | (0.06, 11.53) | (0.00,10.64) | (0.00,6.45) |
| GMT | 23.69 | 3.25 | 8.86 | 26.36 | 1.62 | 6.43 |
| 95% CI | (8.13,68.96) | (1.45,7.29) | (4.46, 17.63) | (8.60,80.85) | (0.91,2.88) | (3.28,12.61) |
| GMFR | 1.35 | 1.29 | 1.32 | 0.94 | 1.03 | 0.99 |
| 95% CI | (0.93,1.96) | (0.84,1.98) | (1.00,1.75) | (0.76,1.17) | (0.72,1.47) | (0.80, 1.21) |

(1) Data analysis cutoff Date: 2025-12-31.

(2)Bivalent vaccine: EV-A71-CVA16 vaccine (EV-A71, Enterovirus 71; CVA16, Coxsackievirus A16).

(3)95% CI, 95% confidence interval.

#### Table S14. Pre-vaccination and 30 days post-full vaccination neutralizing antibody levels by type_EV-A71, CVA16, CVA6, and CVA10 types_6-71 Month(s) of age (HI-mFAS/IPS-6)

| | **Total bivalent vaccine group *VS* EV-A71 vaccine group** | | **Bivalent vaccine group with history of EV-A71 vaccination *VS* No EV-A71 vaccination history bivalent vaccine group** | | **Bivalent vaccine group with history of EV-A71 vaccination *VS* had a history of EV-A71 vaccination EV-A71 vaccine group** | | **Bivalent vaccine group with no history of EV-A71 vaccination *VS* EV-A71 vaccine group without EV-A71 vaccination History** | | **EV-A71 vaccine group with history of EV-A71 vaccination *VS* EV-A71 vaccine group without EV-A71 vaccination history** | |
|---|---|---|---|---|---|---|---|---|---|---|
| | **Rate Difference/GMT Ratio (95% CI)** | ***P* value** | **Rate Difference/GMT Ratio (95% CI)** | ***P* value** | **Rate Difference/GMT Ratio (95% CI)** | ***P* value** | **Rate Difference/GMT Ratio (95% CI)** | ***P* value** | **Rate Difference/GMT Ratio (95% CI)** | ***P* value** |
| **EV-A71** | | | | | | | | | | |
| **Pre-free** | | | | | | | | | | |
| Positive rate n(%) | - | 0.7869 | - | <0.0001 | - | 1.0000 | - | 0.2015 | - | <0.0001 |
| GMT | - | 0.7882 | - | <0.0001 | - | 0.9447 | - | 0.1038 | - | <0.0001 |
| **30 days post full-course Vaccination** | | | | | | | | | | |
| Positive rate (%) | 0.54 (-0.07, 1.16) | 0.0822 | 0.00(-3.74,0.85) | 1.0000 | 0.00(-3.75,4.07) | 1.0000 | 0.64(-0.08,1.37) | 0.0884 | 1.61(-0.20,3.42) | 0.2240 |
| Seroconversion rate (%) | 1.35(-0.09,2.79) | 0.1177 | -4.04(-7.92,-0.16) | 0.0069 | 1.89(-4.79,8.58) | 0.8484 | 1.29 (0.26, 2.32) | 0.0155 | -2.71(-8.72,3.30) | 0.6686 |
| GMT | 2.02 (1.63, 2.50) | <0.0001 | 17.35 (13.33, 22.58) | <0.0001 | 1.53 (0.99, 2.35) | 0.0542 | 2.02 (1.69, 2.41) | <0.0001 | 22.93 (16.25, 32.34) | <0.0001 |
| GMFR | 1.96 (1.60, 2.40) | <0.0001 | 0.09 (0.07, 0.12) | <0.0001 | 1.50 (0.92, 2.46) | 0.1045 | 2.16 (1.80, 2.61) | <0.0001 | 0.13 (0.09, 0.19) | <0.0001 |
| **6 months post full-course Vaccination** | | | | | | | | | | |
| Positive rate (%) | 2.29 (0.90, 3.69) | 0.0015 | 0.24 (-3.58, 1.36) | 1.0000 | 0.00 (-3.83, 4.25) | 1.0000 | 2.76 (1.08, 4.44) | 0.0017 | 2.82 (0.38, 5.27) | 0.1142 |
| GMT | 2.11 (1.69, 2.64) | <0.0001 | 19.27 (14.67, 25.31) | <0.0001 | 1.12 (0.73, 1.71) | 0.6104 | 2.21 (1.86, 2.63) | <0.0001 | 38.16 (27.57, 52.84) | <0.0001 |
| **CVA16** | | | | | | | | | | |
| **Pre-free** | | | | | | | | | | |
| Positive rate n(%) | - | 0.8222 | - | 0.0823 | - | 0.3251 | - | 0.4950 | - | 0.0005 |
| GMT | - | 0.7867 | - | 0.0831 | - | 0.3545 | - | 0.4702 | - | 0.0005 |
| **30 days post full-course** | | | | | | | | | | |

| | | | | | | | | | | |
|---|---|---|---|---|---|---|---|---|---|---|
| **Vaccination** | | | | | | | | | | |
| Positive rate (%) | 73.45 (69.62, 77.28) | <0.0001 | 0.00(-3.78,0.85) | 1.0000 | 55.27 (44.56, 65.99) | <0.0001 | 77.15 (73.18, 81.11) | <0.0001 | 7.57(-5.19,20.34) | 0.4821 |
| Seroconversion rate (%) | 88.22 (85.33, 91.11) | <0.0001 | -0.94(-5.09,3.21) | 0.7492 | 85.84 (78.20, 93.48) | <0.0001 | 88.73 (85.64, 91.83) | <0.0001 | -1.43(-9.72,6.86) | 0.5587 |
| GMT | 237.15 (185.24, 303.60) | <0.0001 | 1.77 (1.33, 2.36) | <0.0001 | 156.03 (83.94, 290.01) | <0.0001 | 254.74 (195.81, 331.42) | <0.0001 | 2.89 (1.62, 5.17) | 0.0004 |
| GMFR | 227.06 (187.39, 275.14) | <0.0001 | 1.13 (0.75, 1.72) | 0.5572 | 221.98 (130.61, 377.29) | <0.0001 | 227.54 (186.14, 278.16) | <0.0001 | 1.16 (0.87, 1.54) | 0.2958 |
| **6 months post full-course Vaccination** | | | | | | | | | | |
| Positive rate (%) | 71.91 (68.04, 75.79) | <0.0001 | 0.00 (-3.82, 0.92) | 1.0000 | 55.17 (44.72, 65.62) | <0.0001 | 75.51 (71.46, 79.56) | <0.0001 | 4.15 (-8.56, 16.86) | 0.5216 |
| GMT | 52.08 (39.44, 68.78) | <0.0001 | 1.89 (1.29, 2.76) | 0.0011 | 30.14 (14.72, 61.71) | <0.0001 | 57.12 (42.47, 76.83) | <0.0001 | 3.58 (1.93, 6.61) | <0.0001 |
| **CVA10** | | | | | | | | | | |
| **Pre-free** | | | | | | | | | | |
| Positive rate n(%) | - | 0.6538 | - | 0.0407 | - | 0.6016 | - | 0.8712 | - | 0.0164 |
| GMT | - | 0.9480 | - | 0.0020 | - | 0.7899 | - | 0.8102 | - | 0.0006 |
| **30 days post full-course Vaccination** | | | | | | | | | | |
| Positive rate (%) | 13.64(-0.12,27.40) | 0.1032 | -5.13(-31.83,21.57) | 0.7123 | 5.23 (-14.87, 25.33) | 0.6127 | 22.28 (3.52, 41.04) | 0.0375 | 48.10 (29.45, 66.75) | 0.0009 |
| Seroconversion rate (%) | 14.15 (3.35, 24.94) | 0.0227 | -9.76(-35.67,16.15) | 0.4466 | 16.84 (1.95, 31.73) | 0.0324 | 11.41 (-4.19, 27.02) | 0.3026 | 2.45 (-11.94, 16.83) | 0.7589 |
| GMT | 1.63 (0.77, 3.47) | 0.2011 | 4.57 (1.66, 12.57) | 0.0032 | 1.49 (0.45, 4.94) | 0.5139 | 1.74 (0.76, 3.96) | 0.1882 | 5.35 (1.91, 14.98) | 0.0014 |
| GMFR | 1.67 (1.16, 2.42) | 0.0064 | 1.02 (0.61, 1.70) | 0.9487 | 1.74 (1.03, 2.95) | 0.0381 | 1.61 (0.94, 2.75) | 0.0819 | 0.94 (0.54, 1.62) | 0.8174 |
| **CVA6** | | | | | | | | | | |
| **Pre-free** | | | | | | | | | | |
| Positive rate n(%) | - | 0.6260 | - | 0.0020 | - | 0.9250 | - | 0.3167 | - | <0.0001 |
| GMT | - | 0.9553 | - | 0.0026 | - | 0.5413 | - | 0.2947 | - | <0.0001 |
| **30 days post full-course** | | | | | | | | | | |

| | | | | | | | | | | |
|---|---|---|---|---|---|---|---|---|---|---|
| **Vaccination** | | | | | | | | | | |
| Positive rate (%) | 5.90(-6.62,18.42) | 0.7224 | 11.51 (-15.31, 38.34) | 0.4158 | 1.16(-19.11,21.42) | 0.9114 | 11.05 (-1.94, 26.13) | 0.0802 | 37.23 (21.92, 54.39)# | <0.0001 |
| Seroconversion rate (%) | 7.53 (1.08, 15.05) | 0.0400 | -9.14(-28.94,10.66) | 0.3048 | 6.34 (-2.69, 15.36) | 0.1779 | 8.74 (-1.99, 19.66) | 0.0684 | -2.08(-8.51,6.52)# | 0.9011 |
| GMT | 1.38 (0.53, 3.60) | 0.5135 | 7.30 (1.95, 27.36) | 0.0032 | 0.90 (0.20, 4.14) | 0.8895 | 2.01 (0.75, 5.37) | 0.1654 | 16.30 (4.79, 55.47) | <0.0001 |
| GMFR | 1.34 (0.95, 1.89) | 0.0934 | 1.05 (0.60, 1.83) | 0.8730 | 1.43 (0.94, 2.19) | 0.0967 | 1.26 (0.73, 2.17) | 0.4123 | 0.92 (0.61, 1.39) | 0.6831 |

(1) Data analysis cutoff Date: 2025-12-31.

(2)Bivalent vaccine: EV-A71-CVA16 vaccine (EV-A71, Enterovirus 71; CVA16, Coxsackievirus A16).

(3)95% CI, 95% confidence interval.

**Table S15. Neutralizing antibodies levels by type at 30 days post full-course vaccination in the pre-vaccination negative population_EV71, CA16, CA6 and CA10 types_6-71 Month(s) of age (HI-mFAS/IPS-6)**

| | Bivalent vaccine Group with History of EV71 Vaccination | Bivalent vaccine Group with No History of EV71 Vaccination | Total Bivalent vaccine Group | EV-A71 vaccine Group with History of EV71 Vaccination | EV-A71 vaccine Group with No History of EV71 Vaccination | EV-A71 vaccine Group Total |
|---|---|---|---|---|---|---|
| EV71 | | | | | | |
| 30 days post full-course Vaccination | | | | | | |
| n(Missing) | 2(0) | 440(0) | 442(0) | 2(0) | 450(0) | 452(0) |
| Seroconversion rate (%) | 100.00 | 100.00 | 100.00 | 100.00 | 99.33 | 99.34 |
| 95% CI (%) | (15.81,100.00) | (99.17,100.00) | (99.17,100.00) | (15.81,100.00) | (98.06, 99.86) | (98.07,99.86) |
| GMT | 25325.42 | 1175.07 | 1191.51 | 998.96 | 576.4 | 577.8 |
| 95% CI | (585.43,1.10E+06) | (1047.54, 1318.13) | (1060.34,1338.92) | (0.00,4.03E+11) | (504.26,658.86) | (505.59, 660.33) |
| 6 months post full-course Vaccination | | | | | | |
| n(Missing) | 1 (0) | 403 (0) | 404 (0) | 2 (0) | 419 (0) | 421 (0) |
| GMT | 6336.00 | 253.59 | 255.61 | 203.91 | 111.94 | 112.26 |
| 95% CI | NA | (226.43, 284.00) | (228.06, 286.50) | (0.02, 1.98E+06) | (98.15, 127.67) | (98.48, 127.97) |
| CA16 | | | | | | |
| 30 days post full-course Vaccination | | | | | | |
| n(Missing) | 67(0) | 343(0) | 410(0) | 56(0) | 364(0) | 420(0) |
| Seroconversion | 100.00 | 100.00 | 100.00 | 10.89 | 4.01 | 4.93 |

| | Bivalent vaccine Group with History of EV71 Vaccination | Bivalent vaccine Group with No History of EV71 Vaccination | Total Bivalent vaccine Group | EV-A71 vaccine Group with History of EV71 Vaccination | EV-A71 vaccine Group with No History of EV71 Vaccination | EV-A71 vaccine Group Total |
|---|---|---|---|---|---|---|
| rate (%) | | | | | | |
| 95% CI (%) | (94.64,100.00) | (98.93,100.00) | (99.10,100.00) | (1.93,19.85) | (1.73,6.29) | (2.60,7.25) |
| GMT | 912.17 | 562.26 | 608.52 | 1.38 | 1.08 | 1.12 |
| 95% CI | (722.99,1150.85) | (495.92,637.46) | (543.33, 681.52) | (0.91, 2.08) | (0.99,1.18) | (1.02,1.23) |
| 6 months post full-course Vaccination | | | | | | |
| n(Missing) | 67 (0) | 317 (0) | 384 (0) | 54 (0) | 340 (0) | 394 (0) |
| GMT | 228.55 | 128.81 | 142.37 | 1.98 | 1.46 | 1.52 |
| 95% CI | (167.63, 311.60) | (109.89, 150.99) | (123.40, 164.25) | (1.15, 3.40) | (1.25, 1.71) | (1.31, 1.77) |
| CA10 | | | | | | |
| 30 days post full-course Vaccination | | | | | | |
| n(Missing) | 24(0) | 33(0) | 57(0) | 21(0) | 33(0) | 54(0) |
| Seroconversion rate (%) | 33.33 | 29.70 | 31.23 | 9.52 | 13.94 | 12.22 |
| 95% CI (%) | (15.63,55.32) | (13.89,45.50) | (19.10, 43.35) | (1.17,30.38) | (1.69, 26.18) | (3.27,21.18) |
| GMT | 2.12 | 1.99 | 2.04 | 1.11 | 1.35 | 1.25 |
| 95% CI | (1.28,3.52) | (1.24, 3.19) | (1.46,2.86) | (0.96, 1.29) | (0.91,2.02) | (0.97,1.61) |
| CA6 | | | | | | |
| 30 days post full-course Vaccination | | | | | | |
| n(Missing) | 26(0) | 39(0) | 65(0) | 25(0) | 43(0) | 68(0) |

| | Bivalent vaccine Group with History of EV71 Vaccination | Bivalent vaccine Group with No History of EV71 Vaccination | Total Bivalent vaccine Group | EV-A71 vaccine Group with History of EV71 Vaccination | EV-A71 vaccine Group with No History of EV71 Vaccination | EV-A71 vaccine Group Total |
|---|---|---|---|---|---|---|
| Seroconversion rate (%) | 3.85 | 7.69 | 6.15 | 4.00 | 4.65 | 2.94 |
| 95% CI (%) | (0.10, 19.64) | (0.00,15.38) | (0.00,12.31) | (0.10,20.35) | (0.00, 11.63) | (0.00,8.82) |
| GMT | 1.33 | 1.33 | 1.33 | 1.09 | 1.15 | 1.13 |
| 95% CI | (0.74,2.38) | (0.85, 2.07) | (0.94,1.87) | (0.91, 1.31) | (0.81, 1.64) | (0.90, 1.42) |

(1)Data analysis cutoff Date: 2025-12-31.

(2)Bivalent vaccine: EV-A71-CVA16 vaccine (EV-A71, Enterovirus 71; CVA16, Coxsackievirus A16).

(3)95% CI, 95% confidence interval.

**Table S16. Neutralizing antibodies levels by type at 30 days post full-course vaccination in pre-vaccination negative population_EV71, CA16, CA6 and CA10 types_6-71 Month(s) of age (HI-mFAS/IPS-6)**

| | Total Bivalent vaccine Group Total for VS EV-A71 vaccine Group | | Bivalent vaccine Group with History of EV71 Vaccination VS No EV71 Vaccination History Bivalent vaccine Group | | Bivalent vaccine Group with History of EV71 Vaccination VS had a history of EV71 Vaccination EV-A71 vaccine Group | | Bivalent vaccine Group with No History of EV71 Vaccination VS EV-A71 vaccine Group without EV71 Vaccination History | | EV-A71 vaccine Group with History of EV71 Vaccination VS EV-A71 vaccine Group without EV71 Vaccination History | |
|---|---|---|---|---|---|---|---|---|---|---|
| | Rate Difference/GMT Ratio (95% CI) | *P* value | Rate Difference/GMT Ratio (95% CI) | *P* value | Rate Difference/GMT Ratio (95% CI) | *P* value | Rate Difference/GMT Ratio (95% CI) | *P* value | Rate Difference/GMT Ratio (95% CI) | *P* value |
| EV71 | | | | | | | | | | |
| 30 days post full-course Vaccination | | | | | | | | | | |
| Seroconversion rate (%) | 0.67(-0.08,1.42) | 0.0841 | 0.00(-65.81,0.87) | 1.0000 | 0.00(-71.92,71.92) | 1.0000 | 0.67(-0.08,1.42) | 0.0841 | 1.68(-0.20,3.56) | 0.8539 |
| GMT | 2.06 (1.73, 2.46) | <0.0001 | 21.55 (2.16, 215.18) | 0.0103 | 25.35 (0.20, 3228.00) | 0.1908 | 2.04 (1.71, 2.44) | <0.0001 | 1.73 (0.25, 11.79) | 0.5740 |
| 6 months post full-course Vaccination | | | | | | | | | | |
| GMT | 2.28 (1.91, 2.71) | <0.0001 | 24.99 (2.56, 243.41) | 0.0057 | 31.07 (0.00, 250364771.10) | 0.2224 | 2.27 (1.90, 2.70) | <0.0001 | 1.82 (0.27, 12.26) | 0.5366 |
| CA16 | | | | | | | | | | |
| 30 days post full-course Vaccination | | | | | | | | | | |
| Seroconversion rate (%) | 95.06 (92.74, 97.38) | <0.0001 | 0.00(-5.44,1.11) | 1.0000 | 89.11 (80.15, 98.07) | <0.0001 | 96.03 (93.77, 98.28) | <0.0001 | 8.35 (-1.05, 17.75) | 0.0663 |
| GMT | 544.55 (470.43, | <0.000 | 1.62 (1.21, 2.17) | 0.001 | 662.49 (413.96, | <0.000 | 519.57 (447.92, | <0.000 | 1.27 (0.89, 1.81) | 0.172 |

| | Total Bivalent vaccine Group Total for VS EV-A71 vaccine Group | | Bivalent vaccine Group with History of EV71 Vaccination VS No EV71 Vaccination History Bivalent vaccine Group | | Bivalent vaccine Group with History of EV71 Vaccination VS had a history of EV71 Vaccination EV-A71 vaccine Group | | Bivalent vaccine Group with No History of EV71 Vaccination VS EV-A71 vaccine Group without EV71 Vaccination History | | EV-A71 vaccine Group with History of EV71 Vaccination VS EV-A71 vaccine Group without EV71 Vaccination History | |
|---|---|---|---|---|---|---|---|---|---|---|
| | **Rate Difference/GMT Ratio (95% CI)** | ***P* value** | **Rate Difference/GMT Ratio (95% CI)** | ***P* value** | **Rate Difference/GMT Ratio (95% CI)** | ***P* value** | **Rate Difference/GMT Ratio (95% CI)** | ***P* value** | **Rate Difference/GMT Ratio (95% CI)** | ***P* value** |
| | 630.35) | 1 | | 1 | 1060.21) | 1 | 602.68) | 1 | | 2 |
| 6 months post full-course Vaccination | | | | | | | | | | |
| GMT | 93.39 (75.82, 115.04) | <0.0001 | 1.77 (1.22, 2.58) | 0.0027 | 115.47 (64.05, 208.17) | <0.0001 | 88.08 (70.60, 109.89) | <0.0001 | 1.35 (0.87, 2.10) | 0.1783 |
| CA10 | | | | | | | | | | |
| 30 days post full-course Vaccination | | | | | | | | | | |
| Seroconversion rate (%) | 19.13 (3.76, 34.50) | 0.0316 | -29.17(-67.65,9.32) | 0.1522 | 23.81 (1.15, 46.47) | 0.0581 | 16.50(-4.75,37.75) | 0.2208 | 9.52 (-3.03, 22.08) | 0.2981 |
| GMT | 1.63 (1.07, 2.48) | 0.0228 | 1.07 (0.54, 2.11) | 0.8496 | 1.91 (1.11, 3.30) | 0.0211 | 1.47 (0.80, 2.71) | 0.2171 | 0.82 (0.50, 1.34) | 0.4269 |
| CA6 | | | | | | | | | | |
| 30 days post full-course Vaccination | | | | | | | | | | |
| Seroconversion rate (%) | 1.74(-5.75,9.37) | 0.5100 | -3.85(-14.10,7.69) | 0.5157 | -0.15(-10.81,10.51) | 0.9776 | 3.04 (-6.74, 13.30) | 0.4278 | -0.65(-9.30,12.00) | 0.9828 |
| GMT | 1.18 (0.78, 1.76) | 0.4314 | 1.00 (0.50, 1.99) | 0.9995 | 1.22 (0.66, 2.23) | 0.5197 | 1.15 (0.66, 2.00) | 0.6096 | 0.95 (0.61, 1.49) | 0.8163 |

(1) Data analysis cutoff Date: 2025-12-31.
(2)Bivalent vaccine: EV-A71-CVA16 vaccine (EV-A71, Enterovirus 71; CVA16, Coxsackievirus A16).
(3)95% CI, 95% confidence interval.

**Table S17. Frequency of IFN-γ cells at each time point before and post immunization (3×10^6 PBMC cells) and their ratio to Baseline_EV-A71 type (CIS)**

| | Bivalent vaccine group (N=19) | EV-A71 vaccine group (N=19) | Test Method | Statistics | *P* value |
|---|---|---|---|---|---|
| **Pre-free** | | | | | |
| n (Missing) | 19 (0) | 18 (1) | | | |
| Mean (SD) | 400.5 (322.9) | 463.9 (422.9) | Wilcoxon Rank-sum Test | 0.015 | 0.9879 |
| Median | 280.0 | 370.0 | | | |
| Q1, Q3 | 150.0, 510.0 | 90.0, 750.0 | | | |
| Min, Max | 50, 1150 | 30, 1520 | | | |
| **14 days post First Dose Vaccination** | | | | | |
| n (Missing) | 16 (3) | 19 (0) | | | |
| Mean (SD) | 1,326.9 (979.4) | 1,933.2 (1,065.3) | Wilcoxon Rank-sum Test | -1.772 | 0.0764 |
| Median | 1,150.0 | 1,780.0 | | | |
| Q1, Q3 | 610.0, 1,985.0 | 1,300.0, 2,730.0 | | | |
| Min, Max | 50, 3300 | 150, 4020 | | | |
| **Ratio of 14 days post First Dose Vaccination to pre-immunization** | | | | | |
| n (Missing) | 16 (3) | 18 (1) | | | |
| Mean (SD) | 6.16 (7.24) | 13.85 (18.17) | Wilcoxon Rank-sum Test | -1.277 | 0.2017 |
| Median | 3.60 | 6.32 | | | |
| Q1, Q3 | 0.97, 8.76 | 1.75, 21.00 | | | |
| Min, Max | 0.3, 26.1 | 0.7, 61.0 | | | |
| **30 days post First Dose Vaccination (before the second dose Vaccination)** | | | | | |
| n (Missing) | 19 (0) | 19 (0) | | | |

| | Bivalent vaccine group (N=19) | EV-A71 vaccine group (N=19) | Test Method | Statistics | *P* value |
|---|---|---|---|---|---|
| Mean (SD) | 1,875.8 (1,338.4) | 1,754.2 (1,285.3) | Wilcoxon Rank-sum Test | 0.234 | 0.8153 |
| Median | 1,900.0 | 1,800.0 | | | |
| Q1, Q3 | 580.0, 2,980.0 | 450.0, 2,670.0 | | | |
| Min, Max | 60, 4460 | 230, 4590 | | | |
| **Ratio of First Dose Vaccination at 30 days post-vaccination (before the second dose) compared to pre-immunization** | | | | | |
| n (Missing) | 19 (0) | 18 (1) | | | |
| Mean (SD) | 7.02 (6.11) | 7.83 (9.79) | Wilcoxon Rank-sum Test | -0.593 | 0.5535 |
| Median | 5.96 | 3.84 | | | |
| Q1, Q3 | 3.38, 8.20 | 2.91, 7.80 | | | |
| Min, Max | 0.2, 24.2 | 0.3, 33.4 | | | |
| **14 days post the 2nd dose of Vaccination** | | | | | |
| n (Missing) | 19 (0) | 18 (1) | | | |
| Mean (SD) | 2,611.1 (1,005.1) | 2,577.8 (1,275.2) | Wilcoxon Rank-sum Test | -0.501 | 0.6161 |
| Median | 3,000.0 | 2,435.0 | | | |
| Q1, Q3 | 1,880.0, 3,390.0 | 1,800.0, 3,440.0 | | | |
| Min, Max | 540, 3890 | 430, 6110 | | | |
| **Ratio of 14 days post the second dose of Vaccination to pre-immunization** | | | | | |
| n (Missing) | 19 (0) | 17 (2) | | | |
| Mean (SD) | 14.10 (16.19) | 17.40 (23.30) | Wilcoxon Rank-sum Test | -0.032 | 0.9747 |
| Median | 5.88 | 6.75 | | | |

| | Bivalent vaccine group (N=19) | EV-A71 vaccine group (N=19) | Test Method | Statistics | *P* value |
|---|---|---|---|---|---|
| Q1, Q3 | 3.33, 21.61 | 2.89, 20.00 | | | |
| Min, Max | 1.3, 62.6 | 1.6, 88.7 | | | |

(1) Data analysis cutoff Date: 2025-12-31.

(2)Bivalent vaccine: EV-A71-CVA16 vaccine (EV-A71, Enterovirus 71; CVA16, Coxsackievirus A16).

**Table S18. Frequency of IFN-γ cells at each time point before and post immunization (3×10^6 PBMC cells) and their ratio to Baseline_CVA16 type (CIS)**

| | Bivalent vaccine group (N=19) | EV-A71 vaccine group (N=19) | Test Method | Statistics | *P* value |
|---|---|---|---|---|---|
| **Pre-free** | | | | | |
| n (Missing) | 17 (2) | 18 (1) | | | |
| Mean (SD) | 389.4 (255.7) | 578.9 (507.8) | Wilcoxon Rank-sum Test | -0.677 | 0.4986 |
| Median | 380.0 | 570.0 | | | |
| Q1, Q3 | 200.0, 520.0 | 120.0, 940.0 | | | |
| Min, Max | 20, 1040 | 50, 1800 | | | |
| **14 days post First Dose Vaccination** | | | | | |
| n (Missing) | 15 (4) | 19 (0) | | | |
| Mean (SD) | 1,566.0 (1,170.4) | 1,880.5 (927.0) | Wilcoxon Rank-sum Test | -1.145 | 0.2523 |
| Median | 1,290.0 | 1,640.0 | | | |
| Q1, Q3 | 650.0, 2,320.0 | 1,200.0, 2,610.0 | | | |
| Min, Max | 50, 3940 | 160, 3660 | | | |
| **Ratio of 14 days post First Dose Vaccination to pre-immunization** | | | | | |
| n (Missing) | 14 (5) | 18 (1) | | | |
| Mean (SD) | 6.69 (6.40) | 9.39 (10.74) | Wilcoxon Rank-sum Test | -0.133 | 0.8942 |
| Median | 4.17 | 4.03 | | | |
| Q1, Q3 | 2.69, 10.37 | 2.08, 13.33 | | | |
| Min, Max | 0.2, 20.9 | 0.7, 34.4 | | | |
| **30 days post First Dose Vaccination (before the second dose Vaccination)** | | | | | |
| n (Missing) | 19 (0) | 19 (0) | | | |

| | Bivalent vaccine group (N=19) | EV-A71 vaccine group (N=19) | Test Method | Statistics | *P* value |
|---|---|---|---|---|---|
| Mean (SD) | 2,040.5 (1,341.1) | 1,833.2 (1,189.9) | Wilcoxon Rank-sum Test | 0.482 | 0.6300 |
| Median | 1,950.0 | 1,990.0 | | | |
| Q1, Q3 | 710.0, 3,310.0 | 530.0, 2,560.0 | | | |
| Min, Max | 90, 4250 | 190, 4020 | | | |
| **Ratio of First Dose Vaccination at 30 days post-vaccination (before the second dose) compared to pre-immunization** | | | | | |
| n (Missing) | 17 (2) | 18 (1) | | | |
| Mean (SD) | 8.17 (8.12) | 6.82 (10.33) | Wilcoxon Rank-sum Test | 1.733 | 0.0831 |
| Median | 5.11 | 3.06 | | | |
| Q1, Q3 | 3.55, 7.92 | 2.12, 4.83 | | | |
| Min, Max | 0.2, 33.0 | 0.2, 38.0 | | | |
| **14 days post the 2nd dose of Vaccination** | | | | | |
| n (Missing) | 16 (3) | 18 (1) | | | |
| Mean (SD) | 2,596.9 (1,307.4) | 2,944.4 (1,344.8) | Wilcoxon Rank-sum Test | -0.518 | 0.6047 |
| Median | 2,740.0 | 2,620.0 | | | |
| Q1, Q3 | 1,905.0, 3,380.0 | 2,260.0, 3,770.0 | | | |
| Min, Max | 510, 5160 | 570, 6150 | | | |
| **Ratio of 14 days post the second dose of Vaccination to pre-immunization** | | | | | |
| n (Missing) | 14 (5) | 17 (2) | | | |
| Mean (SD) | 11.42 (9.50) | 14.31 (17.62) | Wilcoxon Rank-sum Test | 0.536 | 0.5920 |
| Median | 7.06 | 4.56 | | | |

| | Bivalent vaccine group (N=19) | EV-A71 vaccine group (N=19) | Test Method | Statistics | *P* value |
|---|---|---|---|---|---|
| Q1, Q3 | 5.52, 18.00 | 3.12, 17.38 | | | |
| Min, Max | 1.2, 34.0 | 1.3, 55.0 | | | |

(1) Data analysis cutoff Date: 2025-12-31.

(2)Bivalent vaccine: EV-A71-CVA16 vaccine (EV-A71, Enterovirus 71; CVA16, Coxsackievirus A16).

**Table S19. IL-4 cell frequency (3×10^6 PBMC cells) at each time point before and post immunization and its ratio to Baseline_EV-A71 type (CIS)**

| | Bivalent vaccine group (N=19) | EV-A71 vaccine group (N=19) | Test Method | Statistics | *P* value |
|---|---|---|---|---|---|
| **Pre-free** | | | | | |
| n (Missing) | 19 (0) | 19 (0) | | | |
| Mean (SD) | 27.6 (21.9) | 62.4 (76.7) | Wilcoxon Rank-sum Test | -1.100 | 0.2715 |
| Median | 30.0 | 30.0 | | | |
| Q1, Q3 | 15.0, 45.0 | 15.0, 90.0 | | | |
| Min, Max | 0, 75 | 0, 300 | | | |
| **14 days post First Dose Vaccination** | | | | | |
| n (Missing) | 18 (1) | 19 (0) | | | |
| Mean (SD) | 67.5 (109.4) | 85.3 (121.1) | Wilcoxon Rank-sum Test | -1.111 | 0.2666 |
| Median | 37.5 | 45.0 | | | |
| Q1, Q3 | 15.0, 60.0 | 30.0, 90.0 | | | |
| Min, Max | 0, 420 | 15, 495 | | | |
| **Ratio of 14 days post First Dose Vaccination to pre-immunization** | | | | | |
| n (Missing) | 14 (5) | 16 (3) | | | |
| Mean (SD) | 2.06 (2.47) | 1.78 (2.50) | Wilcoxon Rank-sum Test | 0.334 | 0.7384 |
| Median | 1.50 | 1.00 | | | |
| Q1, Q3 | 0.40, 2.50 | 0.42, 2.00 | | | |
| Min, Max | 0.0, 9.3 | 0.2, 10.5 | | | |
| **30 days post First Dose Vaccination (before the second dose Vaccination)** | | | | | |
| n (Missing) | 19 (0) | 19 (0) | | | |

| | Bivalent vaccine group (N=19) | EV-A71 vaccine group (N=19) | Test Method | Statistics | *P* value |
|---|---|---|---|---|---|
| Mean (SD) | 53.7 (51.8) | 64.7 (86.3) | Wilcoxon Rank-sum Test | 0.459 | 0.6462 |
| Median | 30.0 | 15.0 | | | |
| Q1, Q3 | 15.0, 75.0 | 0.0, 105.0 | | | |
| Min, Max | 0, 165 | 0, 315 | | | |
| **Ratio of First Dose Vaccination at 30 days post-vaccination (before the second dose) compared to pre-immunization** | | | | | |
| n (Missing) | 15 (4) | 16 (3) | | | |
| Mean (SD) | 2.11 (1.98) | 1.60 (2.74) | Wilcoxon Rank-sum Test | 1.546 | 0.1221 |
| Median | 1.33 | 0.53 | | | |
| Q1, Q3 | 0.50, 4.00 | 0.06, 1.92 | | | |
| Min, Max | 0.0, 6.0 | 0.0, 10.5 | | | |
| **14 days post the 2nd dose of Vaccination** | | | | | |
| n (Missing) | 19 (0) | 19 (0) | | | |
| Mean (SD) | 33.2 (38.3) | 183.2 (456.9) | Wilcoxon Rank-sum Test | -1.292 | 0.1963 |
| Median | 30.0 | 75.0 | | | |
| Q1, Q3 | 0.0, 45.0 | 0.0, 120.0 | | | |
| Min, Max | 0, 150 | 0, 2025 | | | |
| **Ratio of 14 days post the second dose of Vaccination to pre-immunization** | | | | | |
| n (Missing) | 15 (4) | 16 (3) | | | |
| Mean (SD) | 1.71 (2.51) | 6.74 (17.08) | Wilcoxon Rank-sum Test | -0.040 | 0.9683 |
| Median | 1.00 | 1.26 | | | |

| | Bivalent vaccine group (N=19) | EV-A71 vaccine group (N=19) | Test Method | Statistics | *P* value |
|---|---|---|---|---|---|
| Q1, Q3 | 0.50, 2.00 | 0.00, 3.50 | | | |
| Min, Max | 0.0, 10.0 | 0.0, 67.5 | | | |

(1) Data analysis cutoff Date: 2025-12-31.

(2)Bivalent vaccine: EV-A71-CVA16 vaccine (EV-A71, Enterovirus 71; CVA16, Coxsackievirus A16).

**Table S20. IL-4 cell frequency (3×10^6 PBMC cells) at each time point before and post immunization and its ratio to Baseline_CVA16 type (CIS)**

| | Bivalent vaccine group (N=19) | EV-A71 vaccine group (N=19) | Test Method | Statistics | *P* value |
|---|---|---|---|---|---|
| Pre-free | | | | | |
| n (Missing) | 19 (0) | 19 (0) | | | |
| Mean (SD) | 41.8 (26.7) | 76.6 (90.8) | Wilcoxon Rank-sum Test | -0.297 | 0.7667 |
| Median | 30.0 | 30.0 | | | |
| Q1, Q3 | 30.0, 60.0 | 15.0, 90.0 | | | |
| Min, Max | 0, 105 | 0, 285 | | | |
| 14 days post First Dose Vaccination | | | | | |
| n (Missing) | 18 (1) | 19 (0) | | | |
| Mean (SD) | 61.7 (88.8) | 79.7 (112.7) | Wilcoxon Rank-sum Test | -0.568 | 0.5697 |
| Median | 30.0 | 30.0 | | | |
| Q1, Q3 | 0.0, 75.0 | 15.0, 90.0 | | | |
| Min, Max | 0, 360 | 0, 465 | | | |
| Ratio of 14 days post First Dose Vaccination to pre-immunization | | | | | |
| n (Missing) | 16 (3) | 18 (1) | | | |
| Mean (SD) | 1.51 (2.90) | 2.19 (3.34) | Wilcoxon Rank-sum Test | -0.452 | 0.6513 |
| Median | 0.79 | 0.83 | | | |
| Q1, Q3 | 0.13, 1.46 | 0.20, 2.50 | | | |
| Min, Max | 0.0, 12.0 | 0.0, 13.0 | | | |
| 30 days post First Dose Vaccination (before the second dose Vaccination) | | | | | |
| n (Missing) | 19 (0) | 19 (0) | | | |

| | Bivalent vaccine group (N=19) | EV-A71 vaccine group (N=19) | Test Method | Statistics | *P* value |
|---|---|---|---|---|---|
| Mean (SD) | 56.1 (61.6) | 82.1 (115.4) | Wilcoxon Rank-sum Test | -0.296 | 0.7675 |
| Median | 30.0 | 30.0 | | | |
| Q1, Q3 | 15.0, 75.0 | 15.0, 135.0 | | | |
| Min, Max | 0, 225 | 0, 495 | | | |
| Ratio of First Dose Vaccination at 30 days post-vaccination (before the second dose) compared to pre-immunization | | | | | |
| n (Missing) | 17 (2) | 18 (1) | | | |
| Mean (SD) | 1.31 (1.06) | 2.77 (4.33) | Wilcoxon Rank-sum Test | -0.066 | 0.9472 |
| Median | 1.00 | 0.63 | | | |
| Q1, Q3 | 0.33, 2.25 | 0.26, 4.00 | | | |
| Min, Max | 0.0, 3.0 | 0.0, 16.5 | | | |
| 14 days post the 2nd dose of Vaccination | | | | | |
| n (Missing) | 19 (0) | 18 (1) | | | |
| Mean (SD) | 45.8 (56.9) | 107.5 (164.0) | Wilcoxon Rank-sum Test | 1.259 | 0.2081 |
| Median | 30.0 | 52.5 | | | |
| Q1, Q3 | 15.0, 60.0 | 15.0, 105.0 | | | |
| Min, Max | 0, 180 | 0, 660 | | | |
| Ratio of 14 days post the second dose of Vaccination to pre-immunization | | | | | |
| n (Missing) | 17 (2) | 17 (2) | | | |
| Mean (SD) | 1.37 (1.86) | 2.88 (5.06) | Wilcoxon Rank-sum Test | -0.606 | 0.5443 |
| Median | 0.57 | 1.00 | | | |

| | **Bivalent vaccine group (N=19)** | **EV-A71 vaccine group (N=19)** | **Test Method** | **Statistics** | ***P* value** |
|---|---|---|---|---|---|
| Q1, Q3 | 0.25, 1.50 | 0.11, 2.00 | | | |
| Min, Max | 0.0, 6.0 | 0.0, 19.0 | | | |

(1) Data analysis cutoff Date: 2025-12-31.

(2)Bivalent vaccine: EV-A71-CVA16 vaccine (EV-A71, Enterovirus 71; CVA16, Coxsackievirus A16).

**Table S21. Serious Adverse Event Summary Table_6-71 Month(s) of age (SSS)**

| | Bivalent vaccine group (N=3,989) | | | EV-A71 vaccine group (N=3,986) | | | Total (N=7,975) | | | |
|---|---|---|---|---|---|---|---|---|---|---|
| | **cases** | **Number of cases** | **Incidence rate (%)** | **cases** | **Number of cases** | **Incidence rate (%)** | **cases** | **Number of cases** | **Incidence rate (%)** | ***P* value[1]** |
| **All Serious Adverse Event** | 349 | 248 | 6.22 | 430 | 301 | 7.55 | 779 | 549 | 6.88 | 0.0191 |
| Serious Adverse Event related to the investigational vaccine | 1 | 1 | 0.03 | 0 | 0 | 0.00 | 1 | 1 | 0.01 | 1.0000 |
| Serious Adverse Event Not Related to the Investigational Vaccine | 348 | 248 | 6.22 | 430 | 301 | 7.55 | 778 | 549 | 6.88 | 0.0191 |
| **Serious Adverse Event within 30 days of Vaccination** | 146 | 110 | 2.76 | 180 | 130 | 3.26 | 326 | 240 | 3.01 | 0.1906 |
| Serious Adverse Event related to the investigational vaccine | 1 | 1 | 0.03 | 0 | 0 | 0.00 | 1 | 1 | 0.01 | 1.0000 |
| Serious Adverse Event Not Related to the Investigational Vaccine | 145 | 110 | 2.76 | 180 | 130 | 3.26 | 325 | 240 | 3.01 | 0.1906 |

[1] ***P*** values were calculated using Fisher's exact test.

(1) Serious Adverse events were coded using MedDRA version 27.1.

(2) Related to the investigational Vaccine means the relationship between the Adverse event and the investigational Vaccine is assessed as "possibly related," "probably related," or "definitely related."

(3) Not Related to the investigational Vaccine means the relationship between the Adverse event and the investigational Vaccine is assessed as "Unlikely Related" or "Not Related."

(4) For the non-Reactogenicity subgroup, participant with randomization number R22186 was planned to be in the Bivalent vaccine Group but was erroneously administered two doses of the EV-A71 vaccine Group Vaccine; therefore, in the SSS-related analysis, the participant was analyzed according to the actual group (EV-A71 vaccine Group).

(5) A summary analysis was performed for serious Adverse events occurring within 6 months post the full vaccination course.

(6) Data analysis cutoff Date: 2025-12-31.

(7)Bivalent vaccine: EV-A71-CVA16 vaccine (EV-A71, Enterovirus 71; CVA16, Coxsackievirus A16).

**Table S22. Number and Frequency of Serious Adverse Events_6-71 Month(s) of Age (SSS)**

| Serious Adverse Event Name | Bivalent vaccine group (N=3,989) | | | EV-A71 vaccine group (N=3,986) | | | Total (N=7,975) | | | *P* value[1] |
|---|---|---|---|---|---|---|---|---|---|---|
| | cases | Number of cases | Incidence rate (%) | cases | Number of cases | Incidence rate (%) | cases | Number of cases | Incidence rate (%) | |
| **SAE** | 349 | 248 | 6.22 | 430 | 301 | 7.55 | 779 | 549 | 6.88 | 0.0191 |
| Infections and infestations | 293 | 225 | 5.64 | 358 | 268 | 6.72 | 651 | 493 | 6.18 | 0.0456 |
| Pneumonia | 107 | 101 | 2.53 | 131 | 125 | 3.14 | 238 | 226 | 2.83 | 0.1058 |
| Bronchitis | 29 | 27 | 0.68 | 38 | 36 | 0.90 | 67 | 63 | 0.79 | 0.2584 |
| Upper respiratory tract infection | 17 | 17 | 0.43 | 27 | 26 | 0.65 | 44 | 43 | 0.54 | 0.1729 |
| Pneumonia mycoplasmal | 14 | 14 | 0.35 | 13 | 13 | 0.33 | 27 | 27 | 0.34 | 1.0000 |
| Pharyngitis | 13 | 11 | 0.28 | 19 | 18 | 0.45 | 32 | 29 | 0.36 | 0.1995 |
| Pneumonia viral | 10 | 10 | 0.25 | 12 | 12 | 0.30 | 22 | 22 | 0.28 | 0.6771 |
| Laryngitis | 10 | 9 | 0.23 | 6 | 6 | 0.15 | 16 | 15 | 0.19 | 0.6069 |
| Hand-foot-and-mouth disease | 8 | 8 | 0.20 | 14 | 14 | 0.35 | 22 | 22 | 0.28 | 0.2093 |
| Gastroenteritis rotavirus | 7 | 7 | 0.18 | 10 | 10 | 0.25 | 17 | 17 | 0.21 | 0.4801 |
| Tonsillitis bacterial | 6 | 6 | 0.15 | 6 | 6 | 0.15 | 12 | 12 | 0.15 | 1.0000 |
| Gastroenteritis | 6 | 6 | 0.15 | 6 | 6 | 0.15 | 12 | 12 | 0.15 | 1.0000 |
| Pneumonia haemophilus | 5 | 5 | 0.13 | 1 | 1 | 0.03 | 6 | 6 | 0.08 | 0.2186 |
| Tonsillitis | 4 | 4 | 0.10 | 7 | 6 | 0.15 | 11 | 10 | 0.13 | 0.5484 |
| Influenza | 4 | 4 | 0.10 | 3 | 3 | 0.08 | 7 | 7 | 0.09 | 1.0000 |
| Viral upper respiratory tract infection | 4 | 4 | 0.10 | 2 | 2 | 0.05 | 6 | 6 | 0.08 | 0.6874 |
| Pneumonia respiratory syncytial viral | 3 | 3 | 0.08 | 3 | 3 | 0.08 | 6 | 6 | 0.08 | 1.0000 |
| Laryngopharyngitis | 3 | 3 | 0.08 | 2 | 2 | 0.05 | 5 | 5 | 0.06 | 1.0000 |
| Herpangina | 3 | 3 | 0.08 | 5 | 5 | 0.13 | 8 | 8 | 0.10 | 0.5075 |
| Nasopharyngitis | 3 | 3 | 0.08 | 0 | 0 | 0.00 | 3 | 3 | 0.04 | 0.2499 |
| Oral herpes | 2 | 2 | 0.05 | 0 | 0 | 0.00 | 2 | 2 | 0.03 | 0.4999 |
| Bronchitis mycoplasmal | 2 | 2 | 0.05 | 1 | 1 | 0.03 | 3 | 3 | 0.04 | 1.0000 |
| Bronchiolitis | 2 | 2 | 0.05 | 2 | 2 | 0.05 | 4 | 4 | 0.05 | 1.0000 |

| Serious Adverse Event Name | Bivalent vaccine group (N=3,989) | | | EV-A71 vaccine group (N=3,986) | | | Total (N=7,975) | | | *P* value[1] |
|---|---|---|---|---|---|---|---|---|---|---|
| | cases | Number of cases | Incidence rate (%) | cases | Number of cases | Incidence rate (%) | cases | Number of cases | Incidence rate (%) | |
| Pneumonia bacterial | 2 | 2 | 0.05 | 11 | 11 | 0.28 | 13 | 13 | 0.16 | 0.0128 |
| Enterovirus infection | 2 | 2 | 0.05 | 1 | 1 | 0.03 | 3 | 3 | 0.04 | 1.0000 |
| Pneumonia adenoviral | 2 | 2 | 0.05 | 0 | 0 | 0.00 | 2 | 2 | 0.03 | 0.4999 |
| Pneumonia streptococcal | 2 | 2 | 0.05 | 2 | 2 | 0.05 | 4 | 4 | 0.05 | 1.0000 |
| Rhinitis | 2 | 2 | 0.05 | 0 | 0 | 0.00 | 2 | 2 | 0.03 | 0.4999 |
| COVID-19 | 1 | 1 | 0.03 | 0 | 0 | 0.00 | 1 | 1 | 0.01 | 1.0000 |
| H1N1 influenza | 1 | 1 | 0.03 | 1 | 1 | 0.03 | 2 | 2 | 0.03 | 1.0000 |
| Infectious mononucleosis | 1 | 1 | 0.03 | 0 | 0 | 0.00 | 1 | 1 | 0.01 | 1.0000 |
| Pneumonia klebsiella | 1 | 1 | 0.03 | 0 | 0 | 0.00 | 1 | 1 | 0.01 | 1.0000 |
| Pneumonia parainfluenzae viral | 1 | 1 | 0.03 | 1 | 1 | 0.03 | 2 | 2 | 0.03 | 1.0000 |
| Respiratory tract infection | 1 | 1 | 0.03 | 4 | 4 | 0.10 | 5 | 5 | 0.06 | 0.2185 |
| Bronchitis haemophilus | 1 | 1 | 0.03 | 0 | 0 | 0.00 | 1 | 1 | 0.01 | 1.0000 |
| Exanthema subitum | 1 | 1 | 0.03 | 0 | 0 | 0.00 | 1 | 1 | 0.01 | 1.0000 |
| Diarrhea infectious | 1 | 1 | 0.03 | 3 | 3 | 0.08 | 4 | 4 | 0.05 | 0.3747 |
| Tracheobronchitis mycoplasmal | 1 | 1 | 0.03 | 0 | 0 | 0.00 | 1 | 1 | 0.01 | 1.0000 |
| Mycoplasma infection | 1 | 1 | 0.03 | 1 | 1 | 0.03 | 2 | 2 | 0.03 | 1.0000 |
| Tracheitis | 1 | 1 | 0.03 | 1 | 1 | 0.03 | 2 | 2 | 0.03 | 1.0000 |
| Varicella | 1 | 1 | 0.03 | 0 | 0 | 0.00 | 1 | 1 | 0.01 | 1.0000 |
| Encephalitis herpes | 1 | 1 | 0.03 | 0 | 0 | 0.00 | 1 | 1 | 0.01 | 1.0000 |
| Gastroenteritis viral | 1 | 1 | 0.03 | 1 | 1 | 0.03 | 2 | 2 | 0.03 | 1.0000 |
| Bacterial infection | 1 | 1 | 0.03 | 1 | 1 | 0.03 | 2 | 2 | 0.03 | 1.0000 |
| Tuberculosis | 1 | 1 | 0.03 | 0 | 0 | 0.00 | 1 | 1 | 0.01 | 1.0000 |
| Pneumonia pneumococcal | 1 | 1 | 0.03 | 1 | 1 | 0.03 | 2 | 2 | 0.03 | 1.0000 |
| Encephalitis | 1 | 1 | 0.03 | 0 | 0 | 0.00 | 1 | 1 | 0.01 | 1.0000 |
| Pneumonia chlamydial | 1 | 1 | 0.03 | 0 | 0 | 0.00 | 1 | 1 | 0.01 | 1.0000 |

| Serious Adverse Event Name | Bivalent vaccine group (N=3,989) | | | EV-A71 vaccine group (N=3,986) | | | Total (N=7,975) | | | *P* value[1] |
|---|---|---|---|---|---|---|---|---|---|---|
| | cases | Number of cases | Incidence rate (%) | cases | Number of cases | Incidence rate (%) | cases | Number of cases | Incidence rate (%) | |
| Sinusitis | 1 | 1 | 0.03 | 1 | 1 | 0.03 | 2 | 2 | 0.03 | 1.0000 |
| Lower respiratory tract infection | 0 | 0 | 0.00 | 1 | 1 | 0.03 | 1 | 1 | 0.01 | 0.4998 |
| Otitis media | 0 | 0 | 0.00 | 1 | 1 | 0.03 | 1 | 1 | 0.01 | 0.4998 |
| Paratyphoid fever | 0 | 0 | 0.00 | 1 | 1 | 0.03 | 1 | 1 | 0.01 | 0.4998 |
| Complicated appendicitis | 0 | 0 | 0.00 | 2 | 2 | 0.05 | 2 | 2 | 0.03 | 0.2498 |
| Otitis media acute | 0 | 0 | 0.00 | 1 | 1 | 0.03 | 1 | 1 | 0.01 | 0.4998 |
| Enteritis infectious | 0 | 0 | 0.00 | 1 | 1 | 0.03 | 1 | 1 | 0.01 | 0.4998 |
| Peritonsillar abscess | 0 | 0 | 0.00 | 1 | 1 | 0.03 | 1 | 1 | 0.01 | 0.4998 |
| Pneumonia influenzal | 0 | 0 | 0.00 | 3 | 3 | 0.08 | 3 | 3 | 0.04 | 0.1248 |
| Pertussis-like syndrome | 0 | 0 | 0.00 | 1 | 1 | 0.03 | 1 | 1 | 0.01 | 0.4998 |
| Pharyngitis bacterial | 0 | 0 | 0.00 | 1 | 1 | 0.03 | 1 | 1 | 0.01 | 0.4998 |
| Enterobacter pneumonia | 0 | 0 | 0.00 | 1 | 1 | 0.03 | 1 | 1 | 0.01 | 0.4998 |
| Gastrointestinal infection | 0 | 0 | 0.00 | 1 | 1 | 0.03 | 1 | 1 | 0.01 | 0.4998 |
| Gastrointestinal bacterial infection | 0 | 0 | 0.00 | 1 | 1 | 0.03 | 1 | 1 | 0.01 | 0.4998 |
| Sepsis | 0 | 0 | 0.00 | 1 | 1 | 0.03 | 1 | 1 | 0.01 | 0.4998 |
| Peritonitis | 0 | 0 | 0.00 | 1 | 1 | 0.03 | 1 | 1 | 0.01 | 0.4998 |
| Adenovirus infection | 0 | 0 | 0.00 | 1 | 1 | 0.03 | 1 | 1 | 0.01 | 0.4998 |
| Gastroenteritis norovirus | 0 | 0 | 0.00 | 2 | 2 | 0.05 | 2 | 2 | 0.03 | 0.2498 |
| Gastrointestinal disorders | 23 | 20 | 0.50 | 30 | 29 | 0.73 | 53 | 49 | 0.61 | 0.2011 |
| Enteritis | 8 | 8 | 0.20 | 8 | 8 | 0.20 | 16 | 16 | 0.20 | 1.0000 |
| Intestinal obstruction | 5 | 5 | 0.13 | 4 | 4 | 0.10 | 9 | 9 | 0.11 | 1.0000 |
| Intussusception | 3 | 3 | 0.08 | 3 | 3 | 0.08 | 6 | 6 | 0.08 | 1.0000 |
| Inguinal hernia | 3 | 3 | 0.08 | 1 | 1 | 0.03 | 4 | 4 | 0.05 | 0.6249 |
| Gastritis | 2 | 2 | 0.05 | 4 | 4 | 0.10 | 6 | 6 | 0.08 | 0.4528 |
| Functional gastrointestinal disorder | 1 | 1 | 0.03 | 5 | 5 | 0.13 | 6 | 6 | 0.08 | 0.1247 |

| Serious Adverse Event Name | Bivalent vaccine group (N=3,989) | | | EV-A71 vaccine group (N=3,986) | | | Total (N=7,975) | | | *P* value[1] |
|---|---|---|---|---|---|---|---|---|---|---|
| | cases | Number of cases | Incidence rate (%) | cases | Number of cases | Incidence rate (%) | cases | Number of cases | Incidence rate (%) | |
| Diarrhea | 1 | 1 | 0.03 | 0 | 0 | 0.00 | 1 | 1 | 0.01 | 1.0000 |
| Mouth ulceration | 0 | 0 | 0.00 | 2 | 2 | 0.05 | 2 | 2 | 0.03 | 0.2498 |
| Stomatitis | 0 | 0 | 0.00 | 2 | 2 | 0.05 | 2 | 2 | 0.03 | 0.2498 |
| Infantile Diarrhea | 0 | 0 | 0.00 | 1 | 1 | 0.03 | 1 | 1 | 0.01 | 0.4998 |
| Nervous system disorders | 19 | 18 | 0.45 | 19 | 18 | 0.45 | 38 | 36 | 0.45 | 1.0000 |
| Febrile convulsion | 13 | 12 | 0.30 | 17 | 16 | 0.40 | 30 | 28 | 0.35 | 0.4573 |
| Seizure | 5 | 5 | 0.13 | 2 | 2 | 0.05 | 7 | 7 | 0.09 | 0.4529 |
| Status epilepticus | 1 | 1 | 0.03 | 0 | 0 | 0.00 | 1 | 1 | 0.01 | 1.0000 |
| Injury, poisoning and procedural complications | 5 | 5 | 0.13 | 6 | 6 | 0.15 | 11 | 11 | 0.14 | 0.7742 |
| Toxicity to various agents | 1 | 1 | 0.03 | 0 | 0 | 0.00 | 1 | 1 | 0.01 | 1.0000 |
| Thermal burn | 1 | 1 | 0.03 | 1 | 1 | 0.03 | 2 | 2 | 0.03 | 1.0000 |
| Kidney rupture | 1 | 1 | 0.03 | 0 | 0 | 0.00 | 1 | 1 | 0.01 | 1.0000 |
| Tibia fracture | 1 | 1 | 0.03 | 0 | 0 | 0.00 | 1 | 1 | 0.01 | 1.0000 |
| Arthropod bite | 1 | 1 | 0.03 | 0 | 0 | 0.00 | 1 | 1 | 0.01 | 1.0000 |
| Foreign body in respiratory tract | 0 | 0 | 0.00 | 1 | 1 | 0.03 | 1 | 1 | 0.01 | 0.4998 |
| Hand fracture | 0 | 0 | 0.00 | 1 | 1 | 0.03 | 1 | 1 | 0.01 | 0.4998 |
| Exposure to toxic agent | 0 | 0 | 0.00 | 1 | 1 | 0.03 | 1 | 1 | 0.01 | 0.4998 |
| Limb traumatic amputation | 0 | 0 | 0.00 | 1 | 1 | 0.03 | 1 | 1 | 0.01 | 0.4998 |
| Scrotal injury | 0 | 0 | 0.00 | 1 | 1 | 0.03 | 1 | 1 | 0.01 | 0.4998 |
| General disorders and administration site conditions | 2 | 2 | 0.05 | 2 | 2 | 0.05 | 4 | 4 | 0.05 | 1.0000 |
| Fever | 2 | 2 | 0.05 | 2 | 2 | 0.05 | 4 | 4 | 0.05 | 1.0000 |
| Congenital, familial and genetic disorders | 2 | 2 | 0.05 | 2 | 2 | 0.05 | 4 | 4 | 0.05 | 1.0000 |
| Congenital megacolon | 1 | 1 | 0.03 | 0 | 0 | 0.00 | 1 | 1 | 0.01 | 1.0000 |
| Polydactyly | 1 | 1 | 0.03 | 0 | 0 | 0.00 | 1 | 1 | 0.01 | 1.0000 |
| Hydrocele | 0 | 0 | 0.00 | 2 | 2 | 0.05 | 2 | 2 | 0.03 | 0.2498 |

| Serious Adverse Event Name | Bivalent vaccine group (N=3,989) | | | EV-A71 vaccine group (N=3,986) | | | Total (N=7,975) | | | |
|---|---|---|---|---|---|---|---|---|---|---|
| | cases | Number of cases | Incidence rate (%) | cases | Number of cases | Incidence rate (%) | cases | Number of cases | Incidence rate (%) | *P* value[1] |
| Respiratory, thoracic and mediastinal disorders | 2 | 2 | 0.05 | 2 | 2 | 0.05 | 4 | 4 | 0.05 | 1.0000 |
| Asthma | 1 | 1 | 0.03 | 0 | 0 | 0.00 | 1 | 1 | 0.01 | 1.0000 |
| Adenoidal hypertrophy | 1 | 1 | 0.03 | 0 | 0 | 0.00 | 1 | 1 | 0.01 | 1.0000 |
| Respiratory disorder | 0 | 0 | 0.00 | 1 | 1 | 0.03 | 1 | 1 | 0.01 | 0.4998 |
| Pleurisy | 0 | 0 | 0.00 | 1 | 1 | 0.03 | 1 | 1 | 0.01 | 0.4998 |
| Skin and subcutaneous tissue disorders | 2 | 2 | 0.05 | 2 | 2 | 0.05 | 4 | 4 | 0.05 | 1.0000 |
| Dermatitis contact | 1 | 1 | 0.03 | 0 | 0 | 0.00 | 1 | 1 | 0.01 | 1.0000 |
| Dermatitis allergic | 1 | 1 | 0.03 | 1 | 1 | 0.03 | 2 | 2 | 0.03 | 1.0000 |
| Urticaria | 0 | 0 | 0.00 | 1 | 1 | 0.03 | 1 | 1 | 0.01 | 0.4998 |
| Musculoskeletal and connective tissue disorders | 1 | 1 | 0.03 | 0 | 0 | 0.00 | 1 | 1 | 0.01 | 1.0000 |
| Tenosynovitis stenosans | 1 | 1 | 0.03 | 0 | 0 | 0.00 | 1 | 1 | 0.01 | 1.0000 |
| **Neoplasms benign, malignant and unspecified (incl cysts and polyps)** | 0 | 0 | 0.00 | 1 | 1 | 0.03 | 1 | 1 | 0.01 | 0.4998 |
| Benign neoplasm | 0 | 0 | 0.00 | 1 | 1 | 0.03 | 1 | 1 | 0.01 | 0.4998 |
| Blood and lymphatic system disorders | 0 | 0 | 0.00 | 7 | 7 | 0.18 | 7 | 7 | 0.09 | 0.0078 |
| Immune thrombocytopenia | 0 | 0 | 0.00 | 3 | 3 | 0.08 | 3 | 3 | 0.04 | 0.1248 |
| Mesenteric lymphadenitis | 0 | 0 | 0.00 | 4 | 4 | 0.10 | 4 | 4 | 0.05 | 0.0624 |
| Vascular disorders | 0 | 0 | 0.00 | 1 | 1 | 0.03 | 1 | 1 | 0.01 | 0.4998 |
| Kawasaki's disease | 0 | 0 | 0.00 | 1 | 1 | 0.03 | 1 | 1 | 0.01 | 0.4998 |

[1] *P* values were calculated using Fisher's exact test.

(1) Serious Adverse events were coded using MedDRA version 27.1.

(2) For non-Reactogenicity subgroup, participant with randomization number R22186 was planned to be in the Bivalent vaccine Group but was incorrectly administered two doses of EV-A71 vaccine Group Vaccine; therefore, in the SSS elated analysis, the participant was analyzed according to the actual group (EV-A71 vaccine Group).

(3) A summary analysis was performed for serious Adverse events occurring within 6 months post full-course vaccination.

(4)Data analysis cutoff Date: 2025-12-31.
(5)Bivalent vaccine: EV-A71-CVA16 vaccine (EV-A71, Enterovirus 71; CVA16, Coxsackievirus A16).

**Table S23. Adverse Event Summary_6-71 Month(s) of age Reactogenicity subgroup (ASS)**

| | Bivalent vaccine group (N=993) | | | EV-A71 vaccine group (N=998) | | | Total (N=1,991) | | | |
|---|---|---|---|---|---|---|---|---|---|---|
| | Example Second | Number of cases | Incidence rate (%) | Example Second | Number of cases | Incidence rate (%) | Example Second | Example Number | Incidence rate (%) | ***P*** **value[1]** |
| Adverse event occurring post vaccination | 2,163 | 704 | 70.90 | 2,014 | 657 | 65.83 | 4,177 | 1,361 | 68.36 | 0.0160 |
| Adverse event related to the investigational vaccine | 1,037 | 464 | 46.73 | 756 | 360 | 36.07 | 1,793 | 824 | 41.39 | <0.0001 |
| Adverse event within 30 min | 10 | 10 | 1.01 | 17 | 16 | 1.60 | 27 | 26 | 1.31 | 0.3238 |
| 0-7 days Adverse event | 1,037 | 464 | 46.73 | 754 | 360 | 36.07 | 1,791 | 824 | 41.39 | <0.0001 |
| 8-30 days Adverse event | 0 | 0 | 0.00 | 2 | 1 | 0.10 | 2 | 1 | 0.05 | 1.0000 |
| Grade 1 Adverse event | 423 | 261 | 26.28 | 342 | 202 | 20.24 | 765 | 463 | 23.25 | 0.0015 |
| Grade 2 Adverse event | 567 | 322 | 32.43 | 387 | 244 | 24.45 | 954 | 566 | 28.43 | <0.0001 |
| Grade 3 Adverse event | 47 | 43 | 4.33 | 27 | 24 | 2.40 | 74 | 67 | 3.37 | 0.0182 |
| Grade 4 Adverse event | 0 | 0 | 0.00 | 0 | 0 | 0.00 | 0 | 0 | 0.00 | 1.0000 |
| Grade 5 Adverse event | 0 | 0 | 0.00 | 0 | 0 | 0.00 | 0 | 0 | 0.00 | 1.0000 |
| Solicited Adverse Event | 815 | 405 | 40.79 | 542 | 285 | 28.56 | 1,357 | 690 | 34.66 | <0.0001 |
| Unsolicited Adverse Event | 222 | 149 | 15.01 | 214 | 154 | 15.43 | 436 | 303 | 15.22 | 0.8032 |
| Local (Injection Site) Adverse event | 133 | 95 | 9.57 | 121 | 77 | 7.72 | 254 | 172 | 8.64 | 0.1513 |
| Systemic (non-Injection Site) Adverse event | 904 | 423 | 42.60 | 635 | 323 | 32.36 | 1,539 | 746 | 37.47 | <0.0001 |
| Adverse event not related to the investigational vaccine | 1,126 | 490 | 49.35 | 1,258 | 517 | 51.80 | 2,384 | 1,007 | 50.58 | 0.2821 |
| Grade ≥3 adverse event | 188 | 134 | 13.49 | 225 | 148 | 14.83 | 413 | 282 | 14.16 | 0.4039 |
| ≥Grade 3 Adverse Event Related to the Investigational Vaccine | 47 | 43 | 4.33 | 27 | 24 | 2.40 | 74 | 67 | 3.37 | 0.0182 |
| Adverse event leading to withdrawal | 0 | 0 | 0.00 | 1 | 1 | 0.10 | 1 | 1 | 0.05 | 1.0000 |
| Adverse event leading to withdrawal related to the investigational vaccine | 0 | 0 | 0.00 | 0 | 0 | 0.00 | 0 | 0 | 0.00 | 1.0000 |

[1] ***P*** values were calculated using Fisher's exact test.

(1) Unless otherwise specified, Adverse events were coded using MedDRA version 27.1; solicited Adverse events were summarized using the classifications specified in the protocol.

(2) Related to the investigational Vaccine refers to Adverse events with a relationship to the investigational Vaccine assessed as "definitely related," "probably related," or "possibly related."

(3) Not Related to the investigational Vaccine refers to Adverse events with a relationship to the investigational Vaccine assessed as "Unlikely Related" or "Not Related."

(4) Adverse events occurring post vaccination were defined as those occurring on or post the first dose administration.

(5) Adverse event onset Time (days) = Adverse event start date - date of the dose administration associated with the Adverse event.

(6) A summary analysis was performed for Adverse events occurring within 6 months post the full vaccination course.

(7)Data analysis cutoff Date: 2025-12-31.

(8)Bivalent vaccine: EV-A71-CVA16 vaccine (EV-A71, Enterovirus 71; CVA16, Coxsackievirus A16).

**Table S24. Incidence of AEs post Vaccination (by SOC and PT)_6-71 Month(s) of age Reactogenicity Subgroup (ASS)**

| System Organ Class | Bivalent vaccine group (N=993) | | | EV-A71 vaccine group (N=998) | | | Total (N=1,991) | | | |
|---|---|---|---|---|---|---|---|---|---|---|
| Preferred term | cases | Number of cases | Incidence rate (%) | episodes | Number of cases | Incidence rate (%) | episodes | Number of cases | Incidence rate (%) | *P* value[1] |
| **TEAE** | 1,989 | 678 | 68.28 | 1,775 | 633 | 63.43 | 3,764 | 1,311 | 65.85 | 0.0233 |
| **General disorders and administration site conditions** | 752 | 466 | 46.93 | 514 | 350 | 35.07 | 1,266 | 816 | 40.98 | <0.0001 |
| Fever | 543 | 403 | 40.58 | 337 | 287 | 28.76 | 880 | 690 | 34.66 | <0.0001 |
| Vaccination site pain | 87 | 74 | 7.45 | 58 | 53 | 5.31 | 145 | 127 | 6.38 | 0.0542 |
| Irritability postvaccinal | 40 | 32 | 3.22 | 28 | 24 | 2.40 | 68 | 56 | 2.81 | 0.2813 |
| Decreased activity | 25 | 23 | 2.32 | 16 | 14 | 1.40 | 41 | 37 | 1.86 | 0.1387 |
| Vaccination site swelling | 21 | 20 | 2.01 | 22 | 20 | 2.00 | 43 | 40 | 2.01 | 1.0000 |
| Vaccination site erythema | 14 | 14 | 1.41 | 23 | 22 | 2.20 | 37 | 36 | 1.81 | 0.2387 |
| Fatigue | 11 | 9 | 0.91 | 10 | 10 | 1.00 | 21 | 19 | 0.95 | 1.0000 |
| Vaccination site Itching | 6 | 6 | 0.60 | 9 | 7 | 0.70 | 15 | 13 | 0.65 | 1.0000 |
| Vaccination site induration | 3 | 3 | 0.30 | 7 | 6 | 0.60 | 10 | 9 | 0.45 | 0.5068 |
| Vaccination site rash | 1 | 1 | 0.10 | 2 | 2 | 0.20 | 3 | 3 | 0.15 | 1.0000 |
| acial edema | 1 | 1 | 0.10 | 0 | 0 | 0.00 | 1 | 1 | 0.05 | 0.4987 |
| Chills | 0 | 0 | 0.00 | 1 | 1 | 0.10 | 1 | 1 | 0.05 | 1.0000 |
| Temperature intolerance | 0 | 0 | 0.00 | 1 | 1 | 0.10 | 1 | 1 | 0.05 | 1.0000 |
| **Respiratory, thoracic and mediastinal disorders** | 621 | 345 | 34.74 | 621 | 352 | 35.27 | 1,242 | 697 | 35.01 | 0.8144 |
| Cough | 310 | 257 | 25.88 | 301 | 263 | 26.35 | 611 | 520 | 26.12 | 0.8383 |
| Rhinorrhoea | 244 | 209 | 21.05 | 260 | 215 | 21.54 | 504 | 424 | 21.30 | 0.8267 |
| Nasal obstruction | 20 | 19 | 1.91 | 17 | 17 | 1.70 | 37 | 36 | 1.81 | 0.7400 |
| Oropharyngeal pain | 12 | 12 | 1.21 | 18 | 17 | 1.70 | 30 | 29 | 1.46 | 0.4550 |
| Productive cough | 10 | 10 | 1.01 | 6 | 6 | 0.60 | 16 | 16 | 0.80 | 0.3292 |
| Pharyngeal erythema | 7 | 7 | 0.70 | 4 | 4 | 0.40 | 11 | 11 | 0.55 | 0.3853 |

| System Organ Class<br>Preferred term | Bivalent vaccine group (N=993) | | | EV-A71 vaccine group (N=998) | | | Total (N=1,991) | | | *P* value[1] |
|---|---|---|---|---|---|---|---|---|---|---|
| | cases | Number of cases | Incidence rate (%) | episodes | Number of cases | Incidence rate (%) | episodes | Number of cases | Incidence rate (%) | |
| Sneezing | 6 | 6 | 0.60 | 3 | 2 | 0.20 | 9 | 8 | 0.40 | 0.1778 |
| Allergic rhinitis | 3 | 3 | 0.30 | 1 | 1 | 0.10 | 4 | 4 | 0.20 | 0.3734 |
| Dysphonia | 2 | 2 | 0.20 | 2 | 2 | 0.20 | 4 | 4 | 0.20 | 1.0000 |
| Dyspnoea | 1 | 1 | 0.10 | 2 | 2 | 0.20 | 3 | 3 | 0.15 | 1.0000 |
| Respiratory disorder | 1 | 1 | 0.10 | 1 | 1 | 0.10 | 2 | 2 | 0.10 | 1.0000 |
| Laryngeal inflammation | 1 | 1 | 0.10 | 1 | 1 | 0.10 | 2 | 2 | 0.10 | 1.0000 |
| Sputum retention | 1 | 1 | 0.10 | 0 | 0 | 0.00 | 1 | 1 | 0.05 | 0.4987 |
| Adenoidal hypertrophy | 1 | 1 | 0.10 | 0 | 0 | 0.00 | 1 | 1 | 0.05 | 0.4987 |
| Nasal Itching | 1 | 1 | 0.10 | 0 | 0 | 0.00 | 1 | 1 | 0.05 | 0.4987 |
| Epistaxis | 1 | 1 | 0.10 | 1 | 1 | 0.10 | 2 | 2 | 0.10 | 1.0000 |
| Pharyngeal swelling | 0 | 0 | 0.00 | 1 | 1 | 0.10 | 1 | 1 | 0.05 | 1.0000 |
| Tonsillar erythema | 0 | 0 | 0.00 | 1 | 1 | 0.10 | 1 | 1 | 0.05 | 1.0000 |
| Tonsillar hypertrophy | 0 | 0 | 0.00 | 1 | 1 | 0.10 | 1 | 1 | 0.05 | 1.0000 |
| Allergic cough | 0 | 0 | 0.00 | 1 | 1 | 0.10 | 1 | 1 | 0.05 | 1.0000 |
| **Gastrointestinal disorders** | 312 | 201 | 20.24 | 297 | 200 | 20.04 | 609 | 401 | 20.14 | 0.9112 |
| Diarrhea | 155 | 128 | 12.89 | 138 | 126 | 12.63 | 293 | 254 | 12.76 | 0.8932 |
| Vomiting | 101 | 92 | 9.26 | 103 | 99 | 9.92 | 204 | 191 | 9.59 | 0.6483 |
| Abdominal pain | 12 | 12 | 1.21 | 16 | 16 | 1.60 | 28 | 28 | 1.41 | 0.5689 |
| Nausea | 8 | 7 | 0.70 | 3 | 3 | 0.30 | 11 | 10 | 0.50 | 0.2243 |
| Functional gastrointestinal disorder | 7 | 6 | 0.60 | 14 | 14 | 1.40 | 21 | 20 | 1.00 | 0.1135 |
| Dyspepsia | 6 | 6 | 0.60 | 6 | 6 | 0.60 | 12 | 12 | 0.60 | 1.0000 |
| Mouth ulceration | 5 | 5 | 0.50 | 1 | 1 | 0.10 | 6 | 6 | 0.30 | 0.1236 |
| Enteritis | 3 | 3 | 0.30 | 1 | 1 | 0.10 | 4 | 4 | 0.20 | 0.3734 |
| Infrequent bowel movements | 2 | 2 | 0.20 | 0 | 0 | 0.00 | 2 | 2 | 0.10 | 0.2486 |

| System Organ Class<br>Preferred term | Bivalent vaccine group (N=993) | | | EV-A71 vaccine group (N=998) | | | Total (N=1,991) | | | *P* value[1] |
|---|---|---|---|---|---|---|---|---|---|---|
| | cases | Number of cases | Incidence rate (%) | episodes | Number of cases | Incidence rate (%) | episodes | Number of cases | Incidence rate (%) | |
| Change of bowel habit | 2 | 2 | 0.20 | 2 | 2 | 0.20 | 4 | 4 | 0.20 | 1.0000 |
| Abdominal distension | 2 | 2 | 0.20 | 2 | 2 | 0.20 | 4 | 4 | 0.20 | 1.0000 |
| Abdominal pain upper | 1 | 1 | 0.10 | 0 | 0 | 0.00 | 1 | 1 | 0.05 | 0.4987 |
| Constipation | 1 | 1 | 0.10 | 2 | 2 | 0.20 | 3 | 3 | 0.15 | 1.0000 |
| Infantile spitting up | 1 | 1 | 0.10 | 1 | 1 | 0.10 | 2 | 2 | 0.10 | 1.0000 |
| Toothache | 1 | 1 | 0.10 | 0 | 0 | 0.00 | 1 | 1 | 0.05 | 0.4987 |
| Faeces hard | 1 | 1 | 0.10 | 2 | 2 | 0.20 | 3 | 3 | 0.15 | 1.0000 |
| Intussusception | 1 | 1 | 0.10 | 1 | 1 | 0.10 | 2 | 2 | 0.10 | 1.0000 |
| Intestinal obstruction | 1 | 1 | 0.10 | 0 | 0 | 0.00 | 1 | 1 | 0.05 | 0.4987 |
| Gastritis | 1 | 1 | 0.10 | 2 | 2 | 0.20 | 3 | 3 | 0.15 | 1.0000 |
| Noninfective gingivitis | 1 | 1 | 0.10 | 0 | 0 | 0.00 | 1 | 1 | 0.05 | 0.4987 |
| Dysbiosis | 0 | 0 | 0.00 | 1 | 1 | 0.10 | 1 | 1 | 0.05 | 1.0000 |
| Intra-abdominal fluid collection | 0 | 0 | 0.00 | 1 | 1 | 0.10 | 1 | 1 | 0.05 | 1.0000 |
| Inguinal hernia | 0 | 0 | 0.00 | 1 | 1 | 0.10 | 1 | 1 | 0.05 | 1.0000 |
| **Metabolism and nutrition disorders** | 107 | 93 | 9.37 | 90 | 83 | 8.32 | 197 | 176 | 8.84 | 0.4303 |
| Decreased appetite | 105 | 92 | 9.26 | 83 | 77 | 7.72 | 188 | 169 | 8.49 | 0.2282 |
| Hypomagnesaemia | 1 | 1 | 0.10 | 0 | 0 | 0.00 | 1 | 1 | 0.05 | 0.4987 |
| Dehydration | 1 | 1 | 0.10 | 1 | 1 | 0.10 | 2 | 2 | 0.10 | 1.0000 |
| Hyponatraemia | 0 | 0 | 0.00 | 3 | 3 | 0.30 | 3 | 3 | 0.15 | 0.2496 |
| Hypokalaemia | 0 | 0 | 0.00 | 1 | 1 | 0.10 | 1 | 1 | 0.05 | 1.0000 |
| Hyperuricaemia | 0 | 0 | 0.00 | 1 | 1 | 0.10 | 1 | 1 | 0.05 | 1.0000 |
| Hyperlipidaemia | 0 | 0 | 0.00 | 1 | 1 | 0.10 | 1 | 1 | 0.05 | 1.0000 |
| **Infections and infestations** | 125 | 78 | 7.85 | 162 | 93 | 9.32 | 287 | 171 | 8.59 | 0.2630 |

| System Organ Class | Bivalent vaccine group (N=993) | | | EV-A71 vaccine group (N=998) | | | Total (N=1,991) | | | |
|---|---|---|---|---|---|---|---|---|---|---|
| **Preferred term** | **cases** | **Number of cases** | **Incidence rate (%)** | **episodes** | **Number of cases** | **Incidence rate (%)** | **episodes** | **Number of cases** | **Incidence rate (%)** | ***P* value[1]** |
| Upper respiratory tract infection | 27 | 26 | 2.62 | 22 | 22 | 2.20 | 49 | 48 | 2.41 | 0.5626 |
| Bronchitis | 22 | 20 | 2.01 | 25 | 25 | 2.51 | 47 | 45 | 2.26 | 0.5469 |
| Pneumonia | 19 | 19 | 1.91 | 20 | 20 | 2.00 | 39 | 39 | 1.96 | 1.0000 |
| Mycoplasma infection | 8 | 8 | 0.81 | 9 | 9 | 0.90 | 17 | 17 | 0.85 | 1.0000 |
| Pharyngitis | 7 | 7 | 0.70 | 11 | 11 | 1.10 | 18 | 18 | 0.90 | 0.4787 |
| Gastroenteritis | 5 | 5 | 0.50 | 4 | 4 | 0.40 | 9 | 9 | 0.45 | 0.7528 |
| Tonsillitis | 4 | 4 | 0.40 | 5 | 5 | 0.50 | 9 | 9 | 0.45 | 1.0000 |
| Oral herpes | 3 | 3 | 0.30 | 2 | 2 | 0.20 | 5 | 5 | 0.25 | 0.6864 |
| Laryngitis | 3 | 3 | 0.30 | 3 | 3 | 0.30 | 6 | 6 | 0.30 | 1.0000 |
| Rhinitis | 3 | 3 | 0.30 | 1 | 1 | 0.10 | 4 | 4 | 0.20 | 0.3734 |
| Influenza | 2 | 2 | 0.20 | 9 | 9 | 0.90 | 11 | 11 | 0.55 | 0.0647 |
| Herpes virus infection | 2 | 2 | 0.20 | 1 | 1 | 0.10 | 3 | 3 | 0.15 | 0.6239 |
| Tonsillitis bacterial | 2 | 2 | 0.20 | 1 | 1 | 0.10 | 3 | 3 | 0.15 | 0.6239 |
| Conjunctivitis | 2 | 2 | 0.20 | 2 | 2 | 0.20 | 4 | 4 | 0.20 | 1.0000 |
| Enterovirus infection | 2 | 2 | 0.20 | 1 | 1 | 0.10 | 3 | 3 | 0.15 | 0.6239 |
| Lower respiratory tract infection | 1 | 1 | 0.10 | 1 | 1 | 0.10 | 2 | 2 | 0.10 | 1.0000 |
| Respiratory tract infection | 1 | 1 | 0.10 | 3 | 3 | 0.30 | 4 | 4 | 0.20 | 0.6246 |
| Laryngopharyngitis | 1 | 1 | 0.10 | 0 | 0 | 0.00 | 1 | 1 | 0.05 | 0.4987 |
| Exanthema subitum | 1 | 1 | 0.10 | 0 | 0 | 0.00 | 1 | 1 | 0.05 | 0.4987 |
| Acute sinusitis | 1 | 1 | 0.10 | 0 | 0 | 0.00 | 1 | 1 | 0.05 | 0.4987 |
| Balanoposthitis infective | 1 | 1 | 0.10 | 0 | 0 | 0.00 | 1 | 1 | 0.05 | 0.4987 |
| Tracheobronchitis mycoplasmal | 1 | 1 | 0.10 | 0 | 0 | 0.00 | 1 | 1 | 0.05 | 0.4987 |

| System Organ Class | Bivalent vaccine group (N=993) | | | EV-A71 vaccine group (N=998) | | | Total (N=1,991) | | | |
|---|---|---|---|---|---|---|---|---|---|---|
| Preferred term | cases | Number of cases | Incidence rate (%) | episodes | Number of cases | Incidence rate (%) | episodes | Number of cases | Incidence rate (%) | *P* value[1] |
| Herpangina | 1 | 1 | 0.10 | 2 | 2 | 0.20 | 3 | 3 | 0.15 | 1.0000 |
| Viral rash | 1 | 1 | 0.10 | 1 | 1 | 0.10 | 2 | 2 | 0.10 | 1.0000 |
| Gastroenteritis viral | 1 | 1 | 0.10 | 3 | 3 | 0.30 | 4 | 4 | 0.20 | 0.6246 |
| Hordeolum | 1 | 1 | 0.10 | 0 | 0 | 0.00 | 1 | 1 | 0.05 | 0.4987 |
| Bacterial infection | 2 | 1 | 0.10 | 2 | 2 | 0.20 | 4 | 3 | 0.15 | 1.0000 |
| Adenovirus infection | 1 | 1 | 0.10 | 5 | 5 | 0.50 | 6 | 6 | 0.30 | 0.2181 |
| Pneumonia parainfluenzae viral | 0 | 0 | 0.00 | 1 | 1 | 0.10 | 1 | 1 | 0.05 | 1.0000 |
| Parainfluenzae virus infection | 0 | 0 | 0.00 | 1 | 1 | 0.10 | 1 | 1 | 0.05 | 1.0000 |
| Oral infection | 0 | 0 | 0.00 | 1 | 1 | 0.10 | 1 | 1 | 0.05 | 1.0000 |
| Oral viral infection | 0 | 0 | 0.00 | 1 | 1 | 0.10 | 1 | 1 | 0.05 | 1.0000 |
| Pneumonia respiratory syncytial viral | 0 | 0 | 0.00 | 1 | 1 | 0.10 | 1 | 1 | 0.05 | 1.0000 |
| Pneumonia haemophilus | 0 | 0 | 0.00 | 1 | 1 | 0.10 | 1 | 1 | 0.05 | 1.0000 |
| Complicated appendicitis | 0 | 0 | 0.00 | 1 | 1 | 0.10 | 1 | 1 | 0.05 | 1.0000 |
| Urinary meatitis | 0 | 0 | 0.00 | 1 | 1 | 0.10 | 1 | 1 | 0.05 | 1.0000 |
| Myocarditis infectious | 0 | 0 | 0.00 | 1 | 1 | 0.10 | 1 | 1 | 0.05 | 1.0000 |
| Moraxella infection | 0 | 0 | 0.00 | 1 | 1 | 0.10 | 1 | 1 | 0.05 | 1.0000 |
| Pneumonia mycoplasmal | 0 | 0 | 0.00 | 5 | 5 | 0.50 | 5 | 5 | 0.25 | 0.0622 |
| Pneumonia influenzal | 0 | 0 | 0.00 | 2 | 2 | 0.20 | 2 | 2 | 0.10 | 0.4998 |
| Pneumonia viral | 0 | 0 | 0.00 | 1 | 1 | 0.10 | 1 | 1 | 0.05 | 1.0000 |
| Viral infection | 0 | 0 | 0.00 | 2 | 2 | 0.20 | 2 | 2 | 0.10 | 0.4998 |
| Bronchiolitis | 0 | 0 | 0.00 | 1 | 1 | 0.10 | 1 | 1 | 0.05 | 1.0000 |
| Peritonitis | 0 | 0 | 0.00 | 1 | 1 | 0.10 | 1 | 1 | 0.05 | 1.0000 |
| Chlamydial infection | 0 | 0 | 0.00 | 1 | 1 | 0.10 | 1 | 1 | 0.05 | 1.0000 |

| System Organ Class | Bivalent vaccine group (N=993) | | | EV-A71 vaccine group (N=998) | | | Total (N=1,991) | | | |
|---|---|---|---|---|---|---|---|---|---|---|
| Preferred term | cases | Number of cases | Incidence rate (%) | episodes | Number of cases | Incidence rate (%) | episodes | Number of cases | Incidence rate (%) | *P* value[1] |
| Gastroenteritis rotavirus | 0 | 0 | 0.00 | 1 | 1 | 0.10 | 1 | 1 | 0.05 | 1.0000 |
| Pneumonia streptococcal | 0 | 0 | 0.00 | 1 | 1 | 0.10 | 1 | 1 | 0.05 | 1.0000 |
| Amoebic dysentery | 0 | 0 | 0.00 | 1 | 1 | 0.10 | 1 | 1 | 0.05 | 1.0000 |
| Nasopharyngitis | 0 | 0 | 0.00 | 1 | 1 | 0.10 | 1 | 1 | 0.05 | 1.0000 |
| Rhinovirus infection | 0 | 0 | 0.00 | 2 | 2 | 0.20 | 2 | 2 | 0.10 | 0.4998 |
| **Skin and subcutaneous tissue disorders** | 39 | 36 | 3.63 | 50 | 45 | 4.51 | 89 | 81 | 4.07 | 0.3643 |
| Rash | 19 | 19 | 1.91 | 27 | 26 | 2.61 | 46 | 45 | 2.26 | 0.3659 |
| Rash erythematous | 8 | 8 | 0.81 | 12 | 12 | 1.20 | 20 | 20 | 1.00 | 0.5013 |
| Itching | 4 | 4 | 0.40 | 3 | 3 | 0.30 | 7 | 7 | 0.35 | 0.7255 |
| Erythema | 3 | 3 | 0.30 | 1 | 1 | 0.10 | 4 | 4 | 0.20 | 0.3734 |
| Papule | 1 | 1 | 0.10 | 2 | 2 | 0.20 | 3 | 3 | 0.15 | 1.0000 |
| Dermatitis diaper | 1 | 1 | 0.10 | 2 | 2 | 0.20 | 3 | 3 | 0.15 | 1.0000 |
| Eczema | 1 | 1 | 0.10 | 1 | 1 | 0.10 | 2 | 2 | 0.10 | 1.0000 |
| Urticaria | 1 | 1 | 0.10 | 0 | 0 | 0.00 | 1 | 1 | 0.05 | 0.4987 |
| Dermatitis allergic | 1 | 1 | 0.10 | 0 | 0 | 0.00 | 1 | 1 | 0.05 | 0.4987 |
| Dermatitis | 0 | 0 | 0.00 | 1 | 1 | 0.10 | 1 | 1 | 0.05 | 1.0000 |
| Alopecia | 0 | 0 | 0.00 | 1 | 1 | 0.10 | 1 | 1 | 0.05 | 1.0000 |
| **Investigations** | 8 | 8 | 0.81 | 11 | 11 | 1.10 | 19 | 19 | 0.95 | 0.6460 |
| Enterovirus test positive | 4 | 4 | 0.40 | 5 | 5 | 0.50 | 9 | 9 | 0.45 | 1.0000 |
| Breath sounds abnormal | 3 | 3 | 0.30 | 2 | 2 | 0.20 | 5 | 5 | 0.25 | 0.6864 |
| Coxsackie virus test positive | 1 | 1 | 0.10 | 0 | 0 | 0.00 | 1 | 1 | 0.05 | 0.4987 |
| Myocardial necrosis marker increased | 0 | 0 | 0.00 | 3 | 3 | 0.30 | 3 | 3 | 0.15 | 0.2496 |
| White blood cell count | 0 | 0 | 0.00 | 1 | 1 | 0.10 | 1 | 1 | 0.05 | 1.0000 |

| System Organ Class | Bivalent vaccine group (N=993) | | | EV-A71 vaccine group (N=998) | | | Total (N=1,991) | | | |
|---|---|---|---|---|---|---|---|---|---|---|
| Preferred term | cases | Number of cases | Incidence rate (%) | episodes | Number of cases | Incidence rate (%) | episodes | Number of cases | Incidence rate (%) | *P* value[1] |
| decreased | | | | | | | | | | |
| **Immune system disorders** | 7 | 7 | 0.70 | 2 | 2 | 0.20 | 9 | 9 | 0.45 | 0.1078 |
| Hypersensitivity | 7 | 7 | 0.70 | 2 | 2 | 0.20 | 9 | 9 | 0.45 | 0.1078 |
| **Eye disorders** | 6 | 5 | 0.50 | 2 | 2 | 0.20 | 8 | 7 | 0.35 | 0.2871 |
| Lacrimation increased | 3 | 3 | 0.30 | 0 | 0 | 0.00 | 3 | 3 | 0.15 | 0.1239 |
| Eye discharge | 1 | 1 | 0.10 | 2 | 2 | 0.20 | 3 | 3 | 0.15 | 1.0000 |
| Erythema of eyelid | 1 | 1 | 0.10 | 0 | 0 | 0.00 | 1 | 1 | 0.05 | 0.4987 |
| Swelling of eyelid | 1 | 1 | 0.10 | 0 | 0 | 0.00 | 1 | 1 | 0.05 | 0.4987 |
| **Injury, poisoning and procedural complications** | 3 | 3 | 0.30 | 6 | 5 | 0.50 | 9 | 8 | 0.40 | 0.7260 |
| Animal bite | 1 | 1 | 0.10 | 1 | 1 | 0.10 | 2 | 2 | 0.10 | 1.0000 |
| Head injury | 1 | 1 | 0.10 | 0 | 0 | 0.00 | 1 | 1 | 0.05 | 0.4987 |
| Face injury | 1 | 1 | 0.10 | 0 | 0 | 0.00 | 1 | 1 | 0.05 | 0.4987 |
| Lip injury | 0 | 0 | 0.00 | 1 | 1 | 0.10 | 1 | 1 | 0.05 | 1.0000 |
| Mouth injury | 0 | 0 | 0.00 | 1 | 1 | 0.10 | 1 | 1 | 0.05 | 1.0000 |
| Tooth dislocation | 0 | 0 | 0.00 | 1 | 1 | 0.10 | 1 | 1 | 0.05 | 1.0000 |
| Tibia fracture | 0 | 0 | 0.00 | 1 | 1 | 0.10 | 1 | 1 | 0.05 | 1.0000 |
| Skull fracture | 0 | 0 | 0.00 | 1 | 1 | 0.10 | 1 | 1 | 0.05 | 1.0000 |
| **Nervous system disorders** | 2 | 2 | 0.20 | 4 | 4 | 0.40 | 6 | 6 | 0.30 | 0.6870 |
| Febrile convulsion | 1 | 1 | 0.10 | 3 | 3 | 0.30 | 4 | 4 | 0.20 | 0.6246 |
| Seizure | 1 | 1 | 0.10 | 0 | 0 | 0.00 | 1 | 1 | 0.05 | 0.4987 |
| Headache | 0 | 0 | 0.00 | 1 | 1 | 0.10 | 1 | 1 | 0.05 | 1.0000 |
| **Cardiac disorders** | 2 | 2 | 0.20 | 4 | 4 | 0.40 | 6 | 6 | 0.30 | 0.6870 |
| Myocardial injury | 2 | 2 | 0.20 | 4 | 4 | 0.40 | 6 | 6 | 0.30 | 0.6870 |
| **Ear and labyrinth disorders** | 3 | 2 | 0.20 | 0 | 0 | 0.00 | 3 | 2 | 0.10 | 0.2486 |

| System Organ Class | Bivalent vaccine group (N=993) | | | EV-A71 vaccine group (N=998) | | | Total (N=1,991) | | | |
|---|---|---|---|---|---|---|---|---|---|---|
| **Preferred term** | **cases** | **Number of cases** | **Incidence rate (%)** | **episodes** | **Number of cases** | **Incidence rate (%)** | **episodes** | **Number of cases** | **Incidence rate (%)** | ***P* value[1]** |
| External ear pain | 1 | 1 | 0.10 | 0 | 0 | 0.00 | 1 | 1 | 0.05 | 0.4987 |
| Ear pain | 1 | 1 | 0.10 | 0 | 0 | 0.00 | 1 | 1 | 0.05 | 0.4987 |
| Ear swelling | 1 | 1 | 0.10 | 0 | 0 | 0.00 | 1 | 1 | 0.05 | 0.4987 |
| **Congenital, familial and genetic disorders** | 1 | 1 | 0.10 | 1 | 1 | 0.10 | 2 | 2 | 0.10 | 1.0000 |
| Congenital megacolon | 1 | 1 | 0.10 | 0 | 0 | 0.00 | 1 | 1 | 0.05 | 0.4987 |
| Thalassaemia | 0 | 0 | 0.00 | 1 | 1 | 0.10 | 1 | 1 | 0.05 | 1.0000 |
| **Blood and lymphatic system disorders** | 1 | 1 | 0.10 | 5 | 5 | 0.50 | 6 | 6 | 0.30 | 0.2181 |
| Lymphadenitis | 1 | 1 | 0.10 | 1 | 1 | 0.10 | 2 | 2 | 0.10 | 1.0000 |
| Neutropenia | 0 | 0 | 0.00 | 2 | 2 | 0.20 | 2 | 2 | 0.10 | 0.4998 |
| Anaemia | 0 | 0 | 0.00 | 2 | 2 | 0.20 | 2 | 2 | 0.10 | 0.4998 |
| **Musculoskeletal and connective tissue disorders** | 0 | 0 | 0.00 | 2 | 2 | 0.20 | 2 | 2 | 0.10 | 0.4998 |
| Arthralgia | 0 | 0 | 0.00 | 1 | 1 | 0.10 | 1 | 1 | 0.05 | 1.0000 |
| Myalgia | 0 | 0 | 0.00 | 1 | 1 | 0.10 | 1 | 1 | 0.05 | 1.0000 |
| **Reproductive system and breast disorders** | 0 | 0 | 0.00 | 1 | 1 | 0.10 | 1 | 1 | 0.05 | 1.0000 |
| Pelvic fluid collection | 0 | 0 | 0.00 | 1 | 1 | 0.10 | 1 | 1 | 0.05 | 1.0000 |
| **Vascular disorders** | 0 | 0 | 0.00 | 3 | 3 | 0.30 | 3 | 3 | 0.15 | 0.2496 |
| Scalp haematoma | 0 | 0 | 0.00 | 1 | 1 | 0.10 | 1 | 1 | 0.05 | 1.0000 |
| Flushing | 0 | 0 | 0.00 | 2 | 2 | 0.20 | 2 | 2 | 0.10 | 0.4998 |

[1] ***P*** values were calculated using Fisher's exact test.

(1) Adverse events were coded using MedDRA version 27.1.

(2) A summary analysis was performed for adverse events occurring within 6 months post the full course of vaccination.

(3)Data analysis cutoff Date: 2025-12-31.
(4)Bivalent vaccine: EV-A71-CVA16 vaccine (EV-A71, Enterovirus 71; CVA16, Coxsackievirus A16).

#### Table S25. Adverse Event Summary_6-71 Month(s) of age All randomized Enrollment participants (SSS)

| | Bivalent vaccine group (N=3,989) | | | EV-A71 vaccine group (N=3,986) | | | Total (N=7,975) | | | |
|---|---|---|---|---|---|---|---|---|---|---|
| | Example Second | Example Number | Incidence rate (%) | Example Second | Example Number | Incidence rate (%) | Example Second | Example Number | Incidence rate (%) | *P* value[1] |
| Adverse event occurring post vaccination | 4,058 | 1,489 | 37.33 | 3,806 | 1,380 | 34.62 | 7,864 | 2,869 | 35.97 | 0.0125 |
| Adverse event related to the investigational vaccine | 1,394 | 724 | 18.15 | 1,033 | 537 | 13.47 | 2,427 | 1,261 | 15.81 | <0.0001 |
| Adverse event within 30 min | 45 | 44 | 1.10 | 49 | 46 | 1.15 | 94 | 90 | 1.13 | 0.8331 |
| 0-7 days Adverse event | 1,393 | 724 | 18.15 | 1,029 | 536 | 13.45 | 2,422 | 1,260 | 15.80 | <0.0001 |
| 8-30 days Adverse event | 1 | 1 | 0.03 | 3 | 2 | 0.05 | 4 | 3 | 0.04 | 0.6248 |
| Grade 1 Adverse event | 544 | 366 | 9.18 | 447 | 291 | 7.30 | 991 | 657 | 8.24 | 0.0026 |
| Grade 2 Adverse event | 779 | 487 | 12.21 | 543 | 352 | 8.83 | 1,322 | 839 | 10.52 | <0.0001 |
| Grade 3 Adverse event | 71 | 66 | 1.65 | 43 | 39 | 0.98 | 114 | 105 | 1.32 | 0.0103 |
| Grade 4 Adverse event | 0 | 0 | 0.00 | 0 | 0 | 0.00 | 0 | 0 | 0.00 | 1.0000 |
| Grade 5 Adverse event | 0 | 0 | 0.00 | 0 | 0 | 0.00 | 0 | 0 | 0.00 | 1.0000 |
| Local (Injection Site) Adverse event | 159 | 120 | 3.01 | 142 | 96 | 2.41 | 301 | 216 | 2.71 | 0.1124 |
| Systemic (non-Injection Site) Adverse event | 1,235 | 663 | 16.62 | 891 | 484 | 12.14 | 2,126 | 1,147 | 14.38 | <0.0001 |
| Adverse event not related to the investigational vaccine | 2,664 | 1,120 | 28.08 | 2,773 | 1,134 | 28.45 | 5,437 | 2,254 | 28.26 | 0.7277 |
| Grade ≥3 adverse event | 556 | 347 | 8.70 | 611 | 370 | 9.28 | 1,167 | 717 | 8.99 | 0.3681 |
| ≥Grade 3 Adverse Event Related to the Investigational Vaccine | 71 | 66 | 1.65 | 43 | 39 | 0.98 | 114 | 105 | 1.32 | 0.0103 |
| Adverse event leading to withdrawal | 2 | 2 | 0.05 | 2 | 2 | 0.05 | 4 | 4 | 0.05 | 1.0000 |
| Adverse event leading to withdrawal related to the investigational vaccine | 0 | 0 | 0.00 | 0 | 0 | 0.00 | 0 | 0 | 0.00 | 1.0000 |

[1] ***P*** values were calculated using Fisher's exact test.

(1) Unless otherwise specified, Adverse events were coded using MedDRA version 27.1.

(2) Related to the investigational Vaccine refers to Adverse events with a relationship to the investigational Vaccine classified as "definitely related," "probably related," or "possibly related."

(3) Not Related to the investigational Vaccine refers to Adverse events with a relationship to the investigational Vaccine classified as "Unlikely Related" or "Not Related."

(4) Adverse events occurring post vaccination were defined as those occurring on or post the first dose.

(5) Adverse event onset Time (days) = Adverse event start date - Date of the dose associated with the Adverse event.

(6) For the non-Reactogenicity subgroup, participant with randomization number R22186 was assigned to the Bivalent vaccine Group in the planned group but received two doses of EV-A71 vaccine Group Vaccine erroneously; therefore, in the SSS-related analysis, this participant was analyzed according to the actual group (EV-A71 vaccine Group).

(7) A total of 8 Adverse events occurred in 8 participants (4 events in 4 participants in the Bivalent vaccine Group, 4 events in 4 participants in the EV-A71 vaccine Group) for which the onset time could not be calculated due to missing start dates; these were included in the calculation as Adverse events occurring post vaccination. Among these, one Adverse event in one participant in the EV-A71 vaccine Group was related to the investigational Vaccine, while all other Adverse events were Not Related to the investigational Vaccine.

(8) A summary analysis was performed for Adverse events occurring within 6 months post the full vaccination course.

(9)Data analysis cutoff Date: 2025-12-31.

(10)Bivalent vaccine: EV-A71-CVA16 vaccine (EV-A71, Enterovirus 71; CVA16, Coxsackievirus A16).

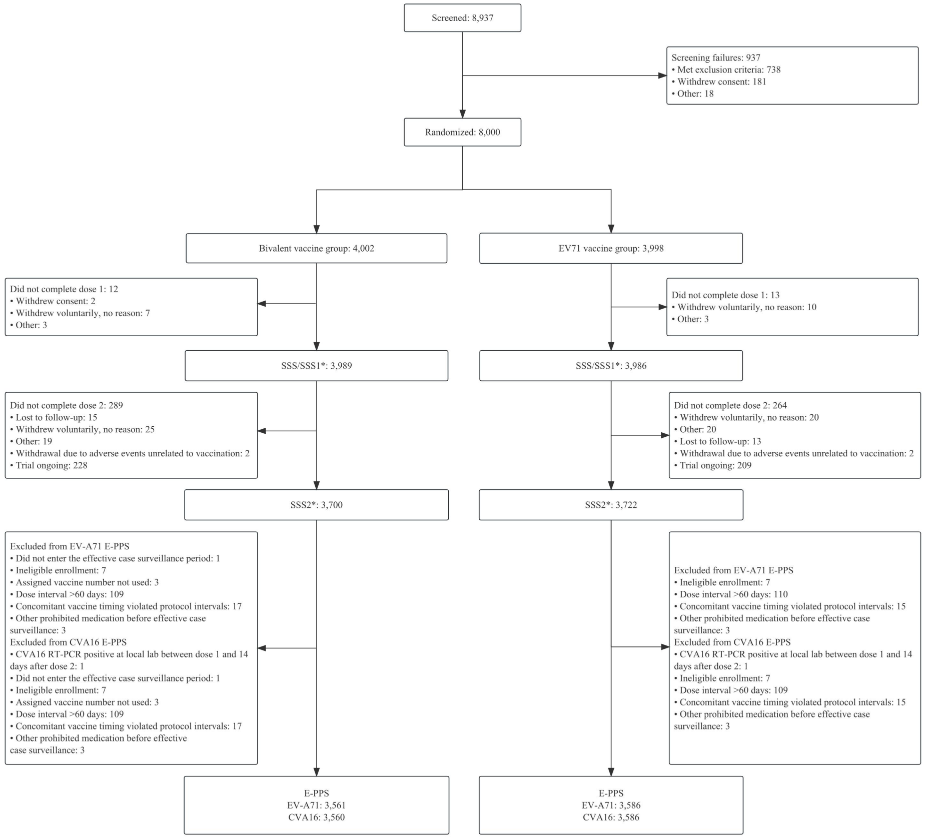

**Figure S1. Screening, Randomization, and Inclusion in the EV-A71-CVA16 Bivalent Vaccine Clinical Trial (E-PPS)**

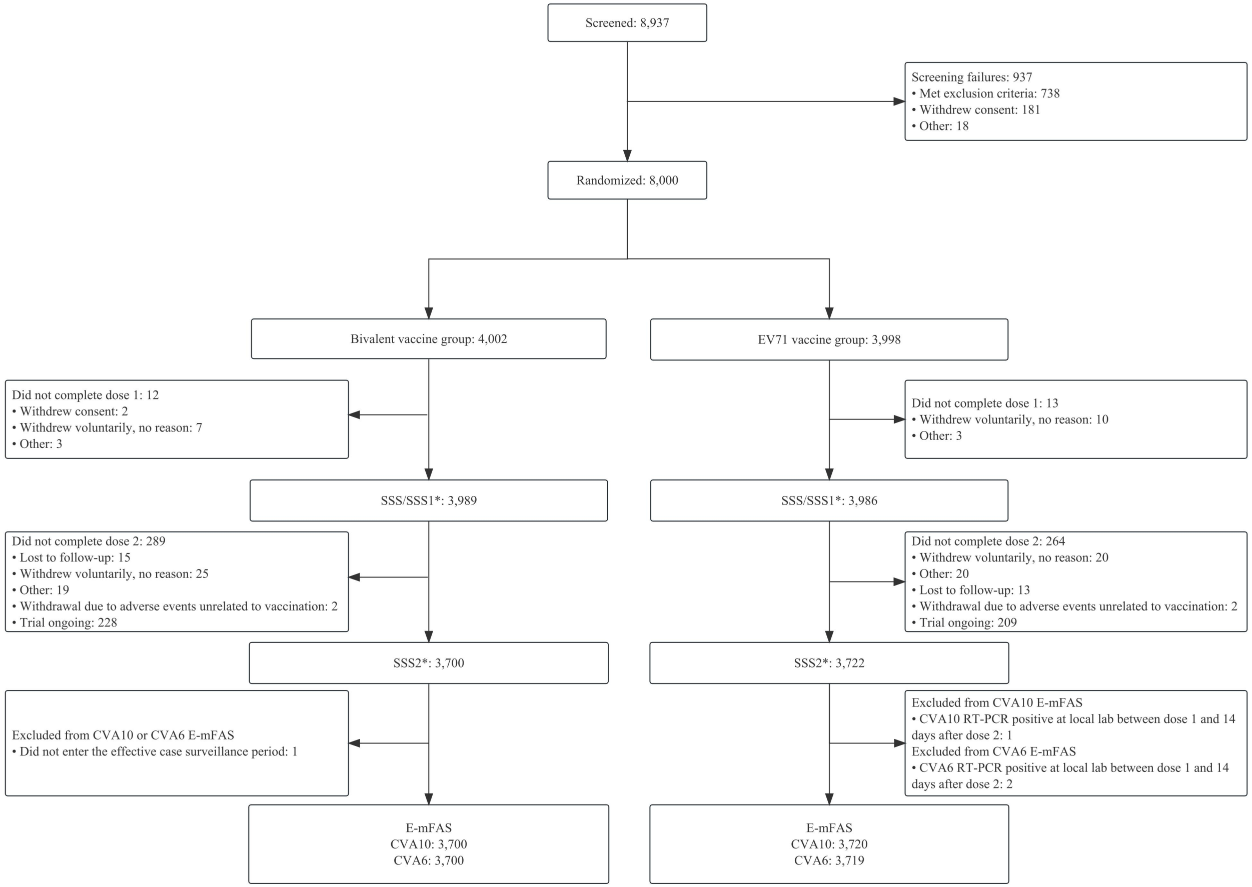

**Figure S2. Screening, randomization, and inclusion in the CVA10 and CVA6 of the bivalent vaccine clinical trial (E-mFAS)**

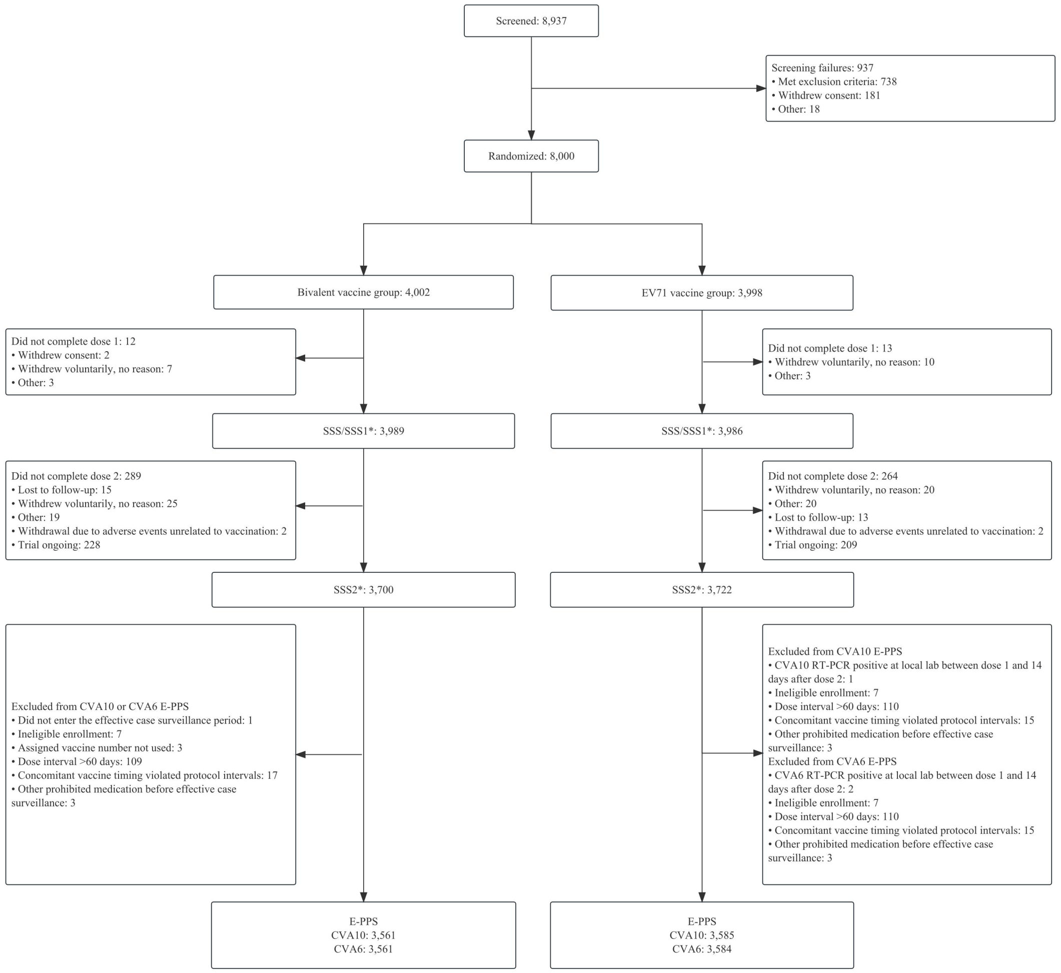

**Figure S3. Screening, randomization, and inclusion in the CVA10 and CVA6 of the bivalent vaccine clinical trial (E-PPS)**

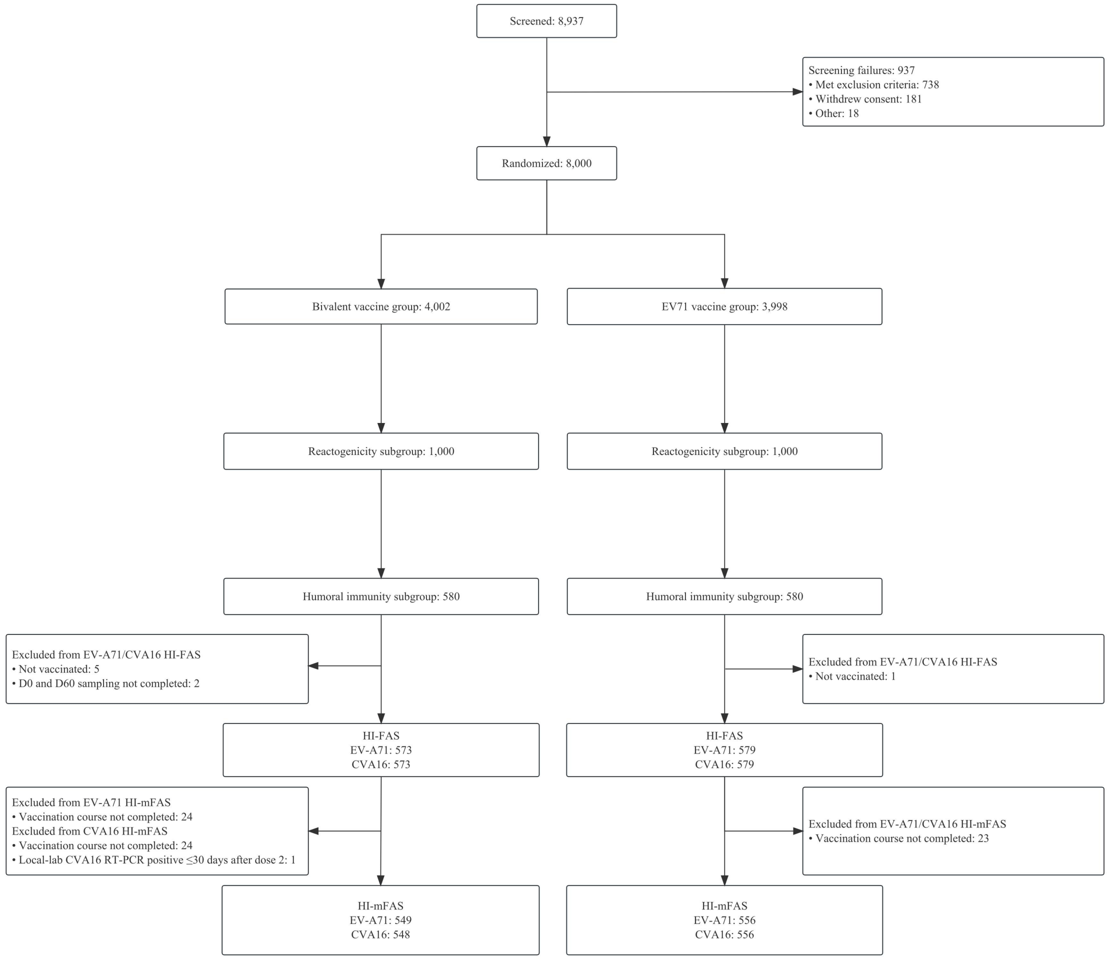

**Figure S4. Participant flow for the humoral immunogenicity analysis sets (HI-mFAS)**

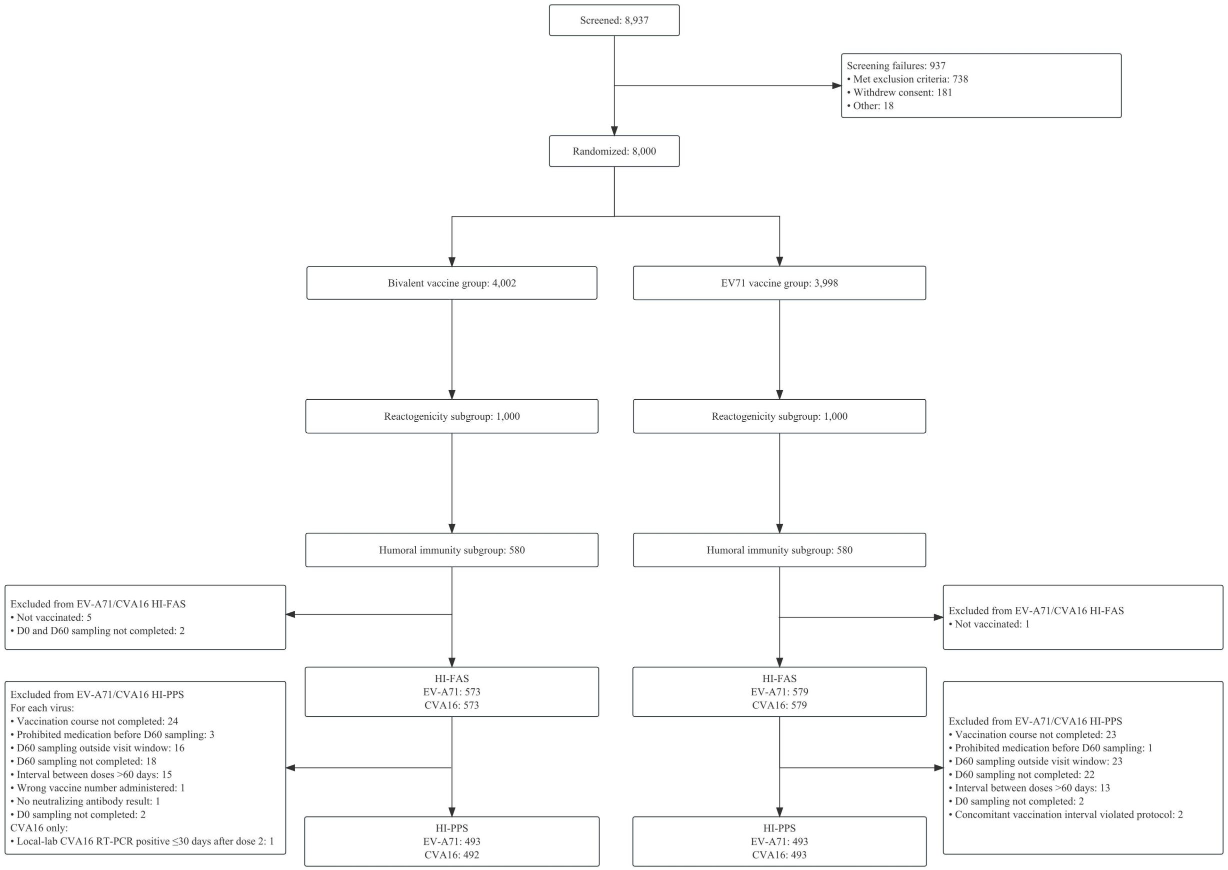

**Figure S5. Participant flow for the humoral immunogenicity per-protocol set (HI-PPS)**

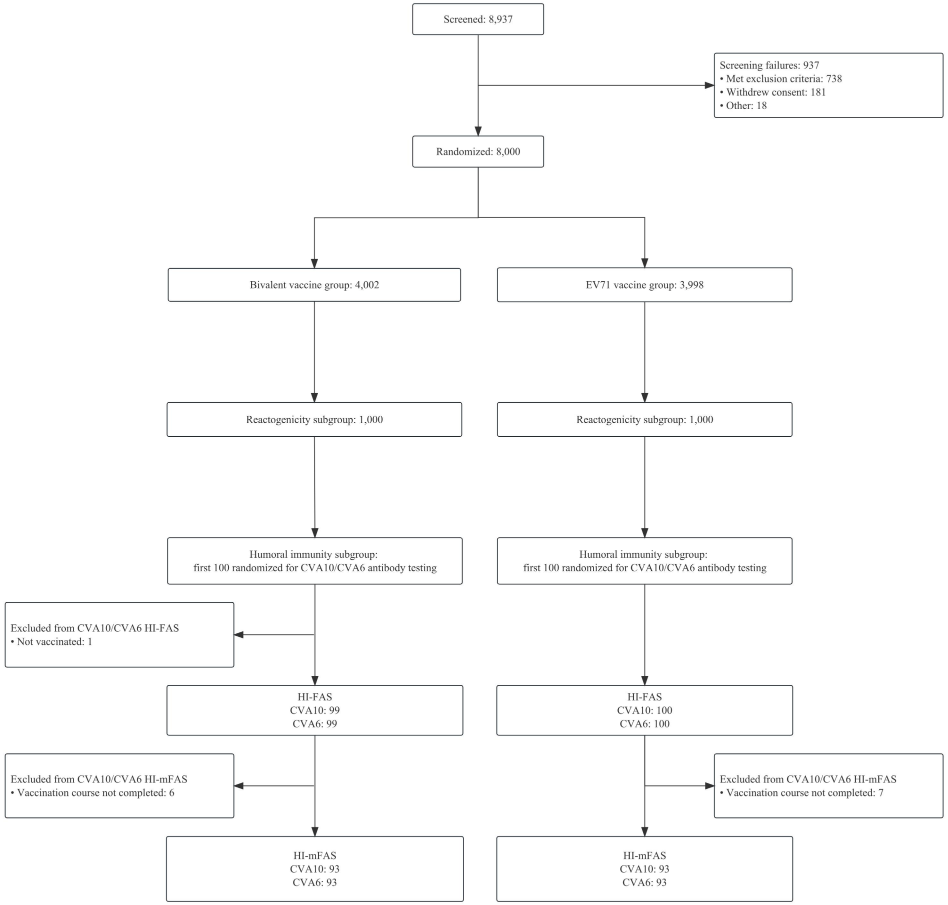

**Figure S6. Participant flow for the cross-reactive immunogenicity against CVA10 and CVA6 (HI-mFAS)**

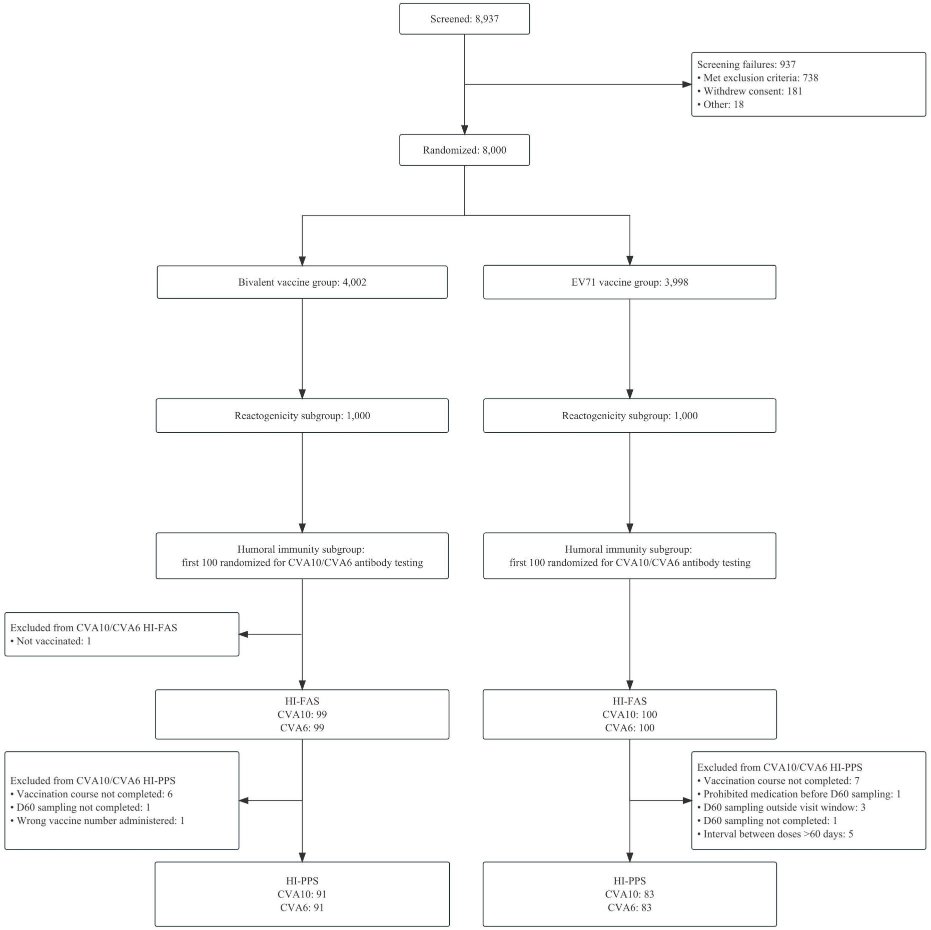

**Figure S7. Participant flow for the cross-reactive immunogenicity against CVA10 and CVA6 (HI-PPS)**

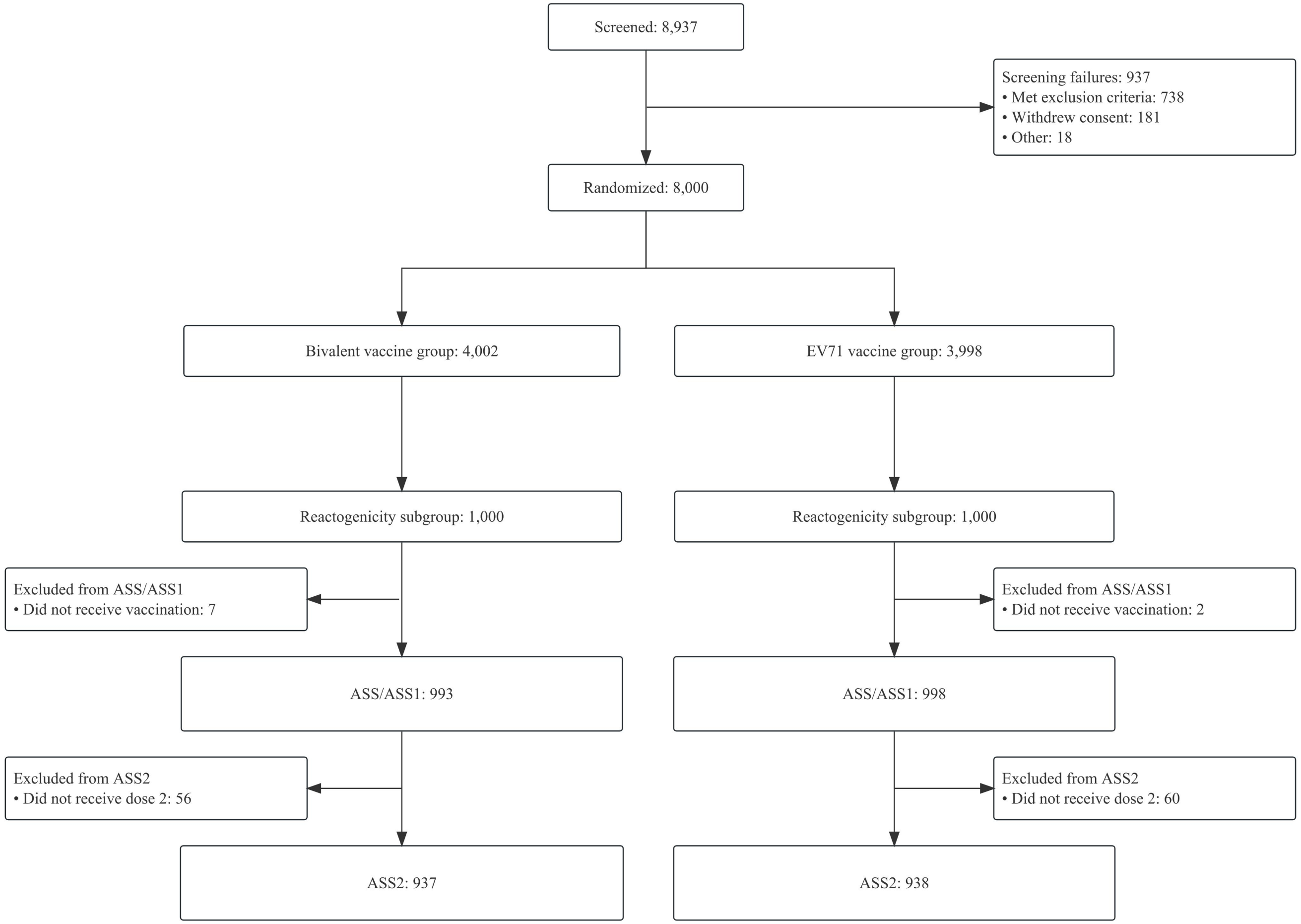

**Figure S8. Participant flow for the safety analysis sets in the reactogenicity subgroup**

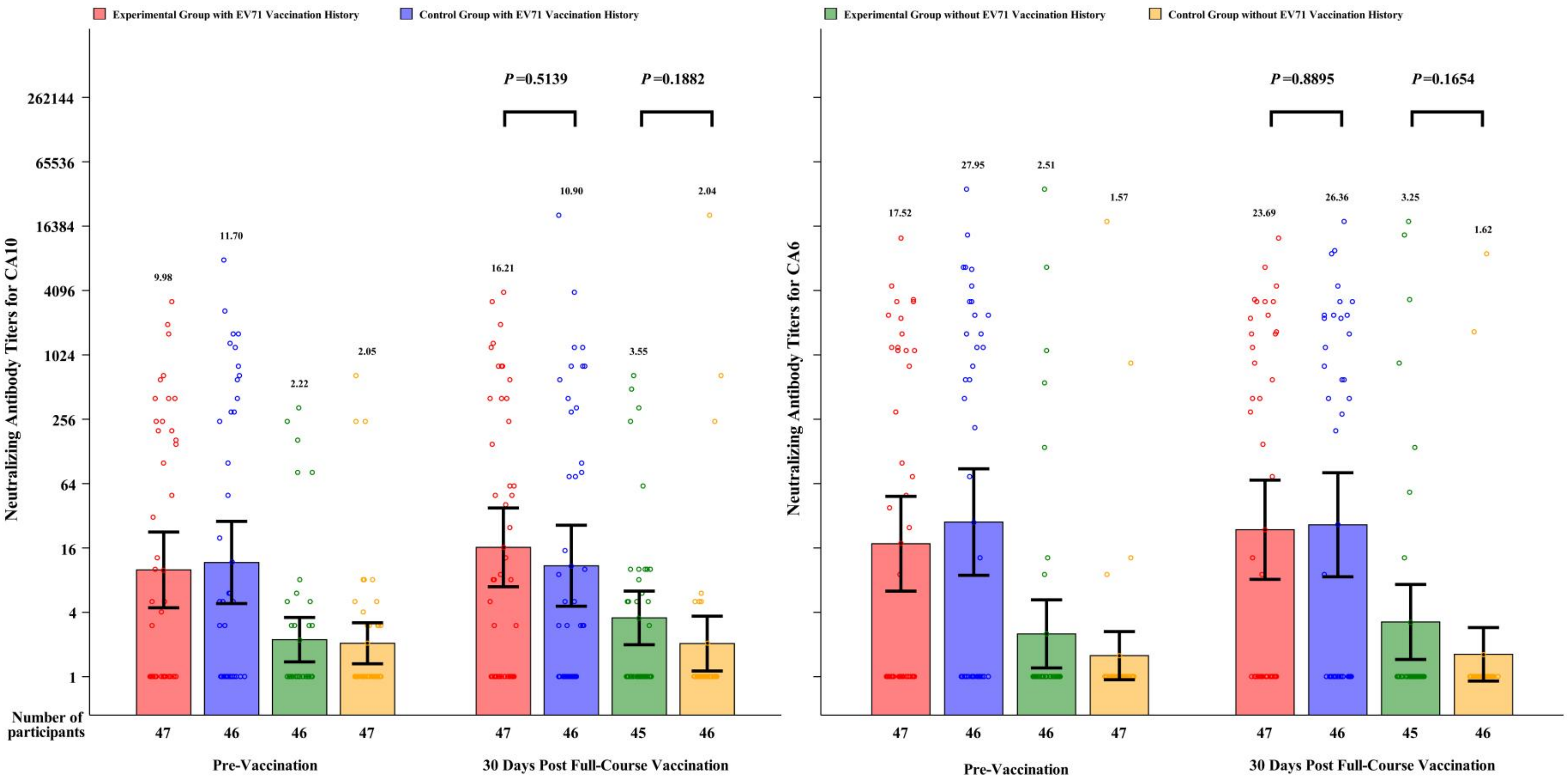

**Figure S9. Histogram of Neutralizing Antibody Level, Aged 6-71 Months (CVA10-HI-mFAS, CVA6-HI-mFAS)**

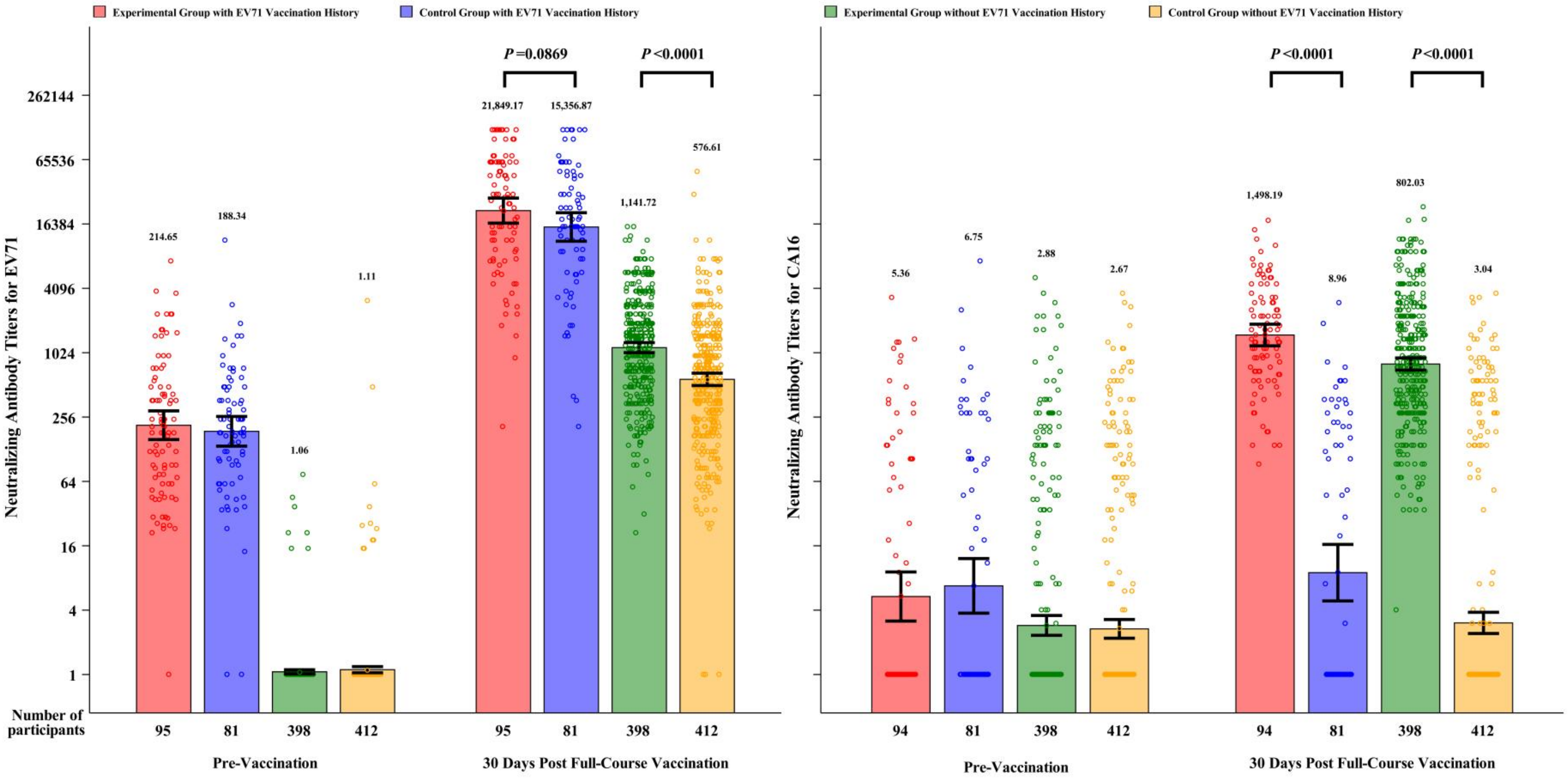

**Figure S10. Histogram of Neutralizing Antibody Level, Aged 6-71 Months (EV-A71-PPS, CVA16-PPS)**

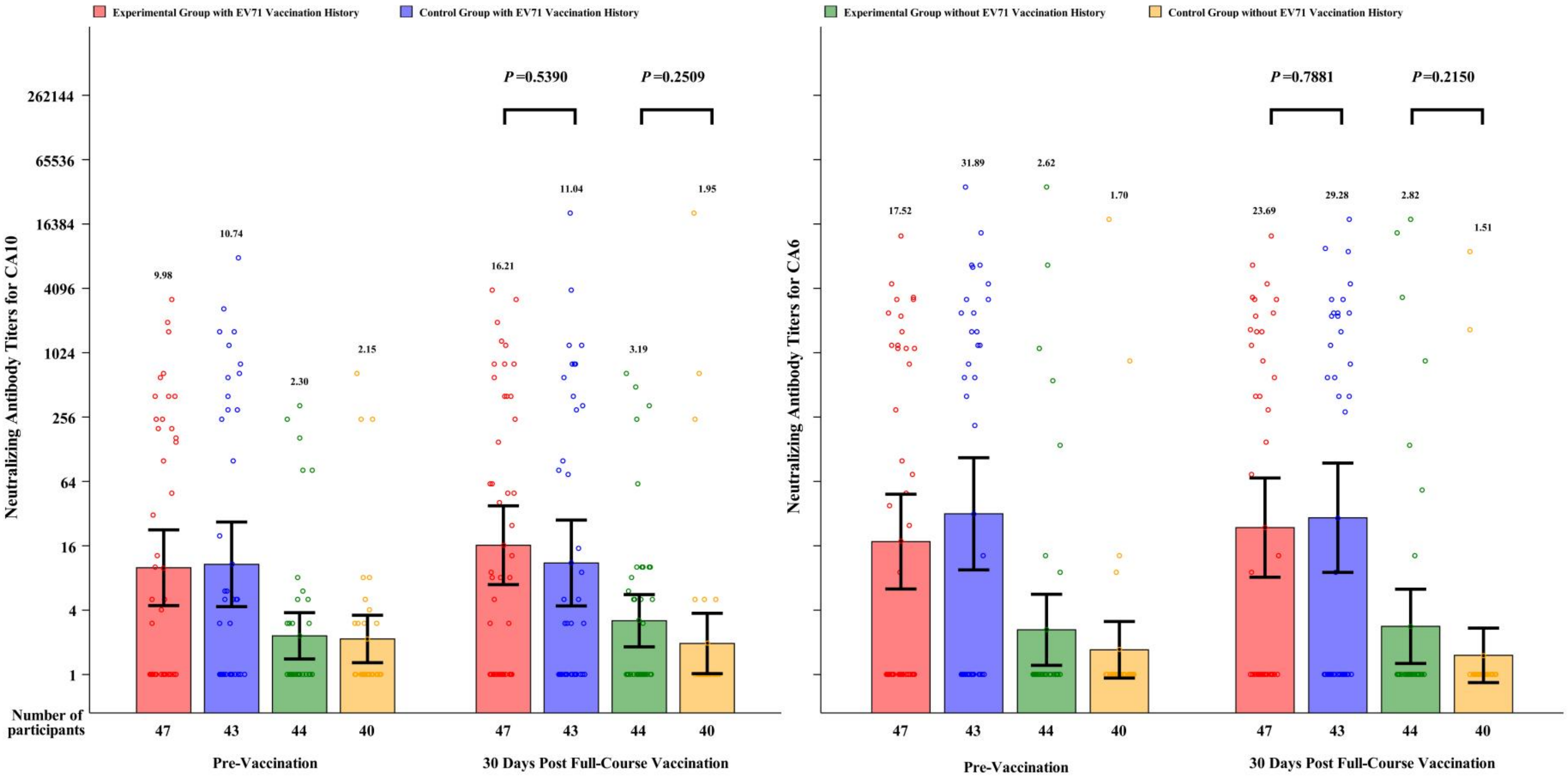

**Figure S11. Histogram of Neutralizing Antibody Level, Aged 6-71 Months (CVA10-PPS,CVA6-PPS)**
